# Comparing mental health treatment outcomes for autistic and non-autistic people: a systematic review

**DOI:** 10.64898/2026.10.08.26364831

**Authors:** Jessica L. Griffiths, Freya Elise, Jordan Thompson, Siying Li, Rachel Rowan Olive, Anam Bhutta, Winnie Liu, Mark Holden, Marshall Mandiangu, Danielle Baptiste, Rebecca Appleton, Phoebe Barnett, Maev Conneely, Lucy Goldsmith, Kristoffer Halvorsrud, Hannah Kate Lewis, Patrick Nyikavaranda, Julian Edbrooke-Childs, William Mandy, Juliana Onwumere, Vaso Totsika, Kylee Trevillion, Brynmor Lloyd-Evans, Alan Simpson, Sonia Johnson

## Abstract

**Background:** Autistic people experience substantially higher rates of mental health conditions and are overrepresented in mental health services. However, most interventions and service models have been developed for non-autistic people, and evidence is needed as to whether there are disparities in outcomes. This systematic review aimed to compare outcomes between autistic and non-autistic people receiving the same mental health interventions or services.

**Methods:** We conducted a systematic review following PRISMA guidelines (PROSPERO: CRD420251021068). We searched academic databases (MEDLINE, Embase, PsycINFO, PubMed, CINAHL), a grey literature database (HMIC), a trial registry (ISRCTN), preprint servers (medRxiv and PsyArXiv), Google, and Google Scholar, to identify relevant papers published 01/01/1992-30/04/2026. We conducted backwards and forwards citation searching and contacted experts. Study quality was assessed using a modified Critical Appraisal Skills Programme Cohort Study Checklist and the Autism-Inclusive Research Assessment (AIRA). Data were synthesised narratively, with random effects meta-analyses where appropriate.

**Results:** Thirty-nine studies were included, comprising 4787730 participants, assessing outcomes of psychological therapies (n = 17) and multi-component or service-level approaches (n = 22). Nearly half (46.2%) were UK-based, and under a quarter (23.1%) included people with intellectual disabilities. Ten studies (25.6%) were rated low quality, twenty-two (56.4%) medium quality, and seven (17.9%) high quality. AIRA ratings highlighted concerns about research inclusiveness for autistic people, including limited involvement of autistic people and lack of validated or adapted outcome measures. Findings varied by intervention: for some, multiple large, higher-quality studies showed less favourable outcomes for autistic people (e.g., less improvement in symptoms or functioning, or higher rates of restrictive practices or acute service use). For other interventions, comparable or mixed outcomes were found across studies, with interpretation often limited by small samples.

**Conclusions:** Mental health treatment outcomes for autistic versus non-autistic people varied across interventions and outcome types. Where significant differences were observed, these usually showed poorer outcomes for autistic people, although they generally reflected smaller improvements rather than a lack of benefit. Action is needed to improve the accessibility and appropriateness of mental health care for autistic people, supported by more high-quality, co-produced research across a broader range of countries and service contexts.

## 1. Introduction

Autistic individuals have a higher risk than the general population of developing a wide range of mental health conditions, including anxiety disorders, depressive disorders, bipolar and related disorders, schizophrenia and psychotic disorders, and obsessive compulsive and related disorders (1). They also experience elevated rates of self-harm (2) and suicidal thoughts and behaviours (3), including deaths by suicide (4).

These co-occurring mental health difficulties contribute to poorer quality of life and wellbeing for autistic people (5,6), increased risk of crisis presentations (7), and greater use of mental health services across the lifespan (8–10). Autistic people are also overrepresented in certain specialist settings, such as eating disorder inpatient units, where one study found that around 30% of patients had high levels of autistic traits (11). National statistical data in England indicate that numbers of autistic people accessing mental health services are increasing (9). This could in part reflect increased rates of autism identification and diagnosis (12,13). However, autism still remains under-recognised in these settings, meaning that the number of autistic people in mental health services is likely to be underestimated (9,14). Furthermore, autistic people admitted to psychiatric hospitals can face longer stays with higher rates of restrictive practices, reflecting gaps in community provision and challenges in delivering appropriate care (15–18).

Additional to the high levels of need, autistic people face multiple barriers to accessing mental health care and receiving care that meets their needs. These include diagnostic overshadowing – where mental health difficulties may be misattributed to autism rather than being recognised and treated – as well as services and interventions that are not adapted to autistic people’s communication styles or sensory and cognitive needs (19,20). In addition, higher rates of psychotropic medication being prescribed to autistic people raises concerns that treatment may focus on the management of observable symptoms (21–23), rather than addressing underlying causes of distress or supporting recovery. Potential sources of disparities in outcomes and experiences include misconceptions and lack of understanding about autism amongst clinicians, and healthcare services and interventions developed primarily for non-autistic populations (19,20,24,25).

Evidence suggests that marginalised and underserved groups, such as autistic people with co-occurring intellectual disabilities, high support needs, or minimal verbal communication (26), ethnic minority groups (27), those assigned female at birth and gender diverse individuals (25,28–30), and older adults (31,32), face additional barriers to accessing appropriate and effective mental health care.

When interventions or services are poorly aligned with autistic people’s needs, this may reduce their accessibility, effectiveness, and even cause iatrogenic harm (unintended harm caused by care or treatment) (19). For instance, inappropriate support may increase distress, hopelessness, and isolation amongst autistic people and their caregivers, potentially exacerbating mental health difficulties and increasing disengagement from services (19,25). Indeed, many autistic people describe negative experiences of mental health services (19,20,31,33). As a result, inadequate mental health support can reinforce a cycle of unmet mental health need amongst autistic people.

Improving mental health support for autistic people has been identified as a top priority for autism research (34,35), and it has been a longstanding national policy ambition both in England (9,35–38) and internationally (39). Identifying where mental health care outcomes differ between autistic and non-autistic people is a crucial step in determining which treatments and service contexts most require adaptation for autistic people.

To our knowledge, this review is the first to systematically compare mental health treatment outcomes for autistic and non-autistic people of all ages, in diverse settings, and across a range of mental health difficulties, interventions, and outcomes. The findings could support researchers, clinicians, and policymakers in targeting areas for improvement, including tailoring interventions for autistic people, and to identify evidence gaps to guide future research, ultimately helping to ensure that autistic people receive appropriate and effective mental health support.

In this review, quantitative evidence is synthesised to answer the following research question: do mental health treatment outcomes differ between autistic and non-autistic people receiving the same services or interventions?

## 2. Methods

We conducted a systematic review following the Preferred Reporting Items for Systematic Review and Meta-Analyses (PRISMA) statement (40). The PRISMA checklist can be found in Appendix A [see Additional File 1]. The protocol for our review was prospectively registered with PROSPERO (CRD420251021068). The rationale for a deviation from the registered protocol regarding the planned use of GRADE is provided in Appendix B [see Additional File 1].

The review was conducted by the National Institute for Health and Care Research (NIHR) Policy Research Unit in Mental Health (MHPRU) based at King’s College London, University College London, and University of York. The NIHR MHPRU conducts research in response to policymaker need (e.g., in the Department of Health and Social Care or NHS England).

### 2.1. Research team positionality

This review was conducted by a multidisciplinary working group comprising academic and lived experience researchers, clinicians, and a policymaker, bringing diverse perspectives shaped by clinical, policy, research and personal or caregiver experience of mental health difficulties and services. Team members’ experiences with autism research varied, from those new to the field to those with extensive experience. Clinical expertise included mental health nursing, clinical psychology, psychiatry, and social work. The team comprised autistic and non-autistic members, including individuals with lived experience of mental health difficulties, accessing services, and/or supporting someone who has. Autistic team members had varied experiences and perspectives on diagnosis and how it intersected with mental health.

The working group was diverse in professional background, gender, and age (all adults, though some members had lived experience of mental health difficulties in childhood). Whilst some ethnic diversity was present, particularly among lived experience researchers, the group was predominantly White ethnicities. All members were UK-based. No members reporting having an intellectual disability or were minimally verbal, though some had cared for autistic individuals with substantial support needs.

We approached the review from a neurodiversity-affirming perspective, viewing autism as a form of neurodevelopmental variation (41). Autism and mental health difficulties were viewed as distinct; autism was not viewed as something to be ‘treated’, but as something to be taken into account when supporting individuals’ mental health. We recognise that perspectives on neurodiversity-affirming language and practice vary across cultural contexts and over time, and that our views are shaped by contemporary UK-based research, policy, and community discourse. Working group members’ lived experiences, demographics, professional identities, and cultural context likely influenced the research process, including framing of the review question, scope, development and application of the quality appraisal tools, and interpretation of findings.

Seventeen working group meetings were held, approximately monthly, throughout the review, with input sought on all stages, including refining the research question, developing the protocol, deciding scope, making decisions about study eligibility, specifying data extraction, developing and applying quality appraisal criteria, and interpreting and presenting findings. Additional meetings with lived experience researchers enabled more in-depth discussion and input on key aspects of the review, including, for example, refining the quality appraisal criteria, discussing quality appraisal ratings, and interpreting the findings.

### 2.2. Lived experience involvement

An academic researcher (FE) is autistic and has lived experience of mental health difficulties and services, meaning lived experience informed both the leadership and conduct of the review. The working group also included eight lived experience researchers, who contributed their expertise from their own experiences to all stages of the review, from developing the protocol and adapting the quality appraisal tools to providing input on screening, data extraction, quality appraisal, analysis and write-up. All lived experience researchers had lived experience of mental health difficulties. Several lived experience researchers also had lived experience of autism, including those who had an autism diagnosis, suspected they may be autistic, and some who had experience caring for autistic people, including one carer who was not autistic themselves. Two autistic researchers with experience of mental health difficulties wrote the lived experience commentary (RRO, DB).

### 2.3. Search strategy

We searched five electronic databases (MEDLINE via Ovid, Embase via Ovid, PsycInfo via Ovid, PubMed, and CINAHL) for peer-reviewed literature relevant to our research question. We searched for relevant grey literature in an electronic grey literature database (HMIC), two pre-print servers (medRxiv and PsyArXiv, via Europe PMC), a clinical trial repository (ISRCTN), and using Google and Google Scholar (limiting screening to the first 10 pages of results). Searches were initially conducted between 7^th^ April 2025 and 11^th^ April 2025, with no language or country restrictions, and updated on 30^th^ April 2026. Searches were limited to articles published from 1^st^ January 1992 to the date of the searches (to cover the Diagnostic and Statistical Manual of Mental Disorders, Fourth Edition [DSM-IV], Diagnostic and Statistical Manual of Mental Disorders, Fifth Edition [DSM-5], International Classification of Diseases, Tenth Revision [ICD-10], and International Classification of Diseases, Eleventh Edition [ICD-11] periods). Screening of non-English language papers was conducted using Google Translate, with plans to check interpretations with someone with knowledge of the language if papers appeared relevant.

We contacted 22 experts suggested by working group members, including a range of academic, clinical, and lived experience experts and relevant organisations/groups, to request additional grey literature we may not have identified. We also conducted forwards and backwards citation searching of our included papers, using Citation Chaser (42), to identify relevant literature. Our search strategy included key terms relating to autism, mental health, and interventions, as detailed in Appendix C [see Additional File 1].

### 2.4. Screening

Title and abstract and full text screening were conducted in Covidence, a systematic review management software (43). Title and abstract screening were conducted by four researchers (JG, FE, JT, KT). All titles and abstracts were independently double screened. Full text screening was conducted by three researchers (JG, FE, JT). All full texts were independently double screened. Any disagreements were resolved by discussion between JG, FE, JT, or the wider working group, where necessary.

### 2.5. Eligibility criteria

#### 2.5.1. Participants

Samples in eligible studies included individuals of all ages formally diagnosed as autistic, who met or exceeded cut-off scores on screening tools for autism, or were suspected to be autistic based on self-identification or clinician assessment. Participants must have received, or be receiving, mental health assessment or care for a mental health condition. Autistic participants must have been a distinct subsample for whom data were reported separately.

We excluded studies where autistic participants were mixed within a broader group and their data could not be separately extracted, and research about organic conditions (e.g., dementia) due to differences in clinical course and care pathways, which would have introduced substantial clinical heterogeneity.

#### 2.5.2. Intervention

We included any form of assessment, intervention, or service that aimed to target mental health problem(s), delivered by mental health staff, including: pharmacological treatments (e.g., antidepressants, antipsychotics), non-pharmacological interventions (e.g., psychotherapy, behavioural interventions, peer support), multi-component interventions (e.g., mental health care delivered by more than one person or by a team, or interventions with multiple elements), and mental health services or teams.

#### 2.5.3. Comparators

We included studies that compared mental health treatment outcomes for autistic individuals to those for non-autistic individuals, who are receiving a comparable mental health service or intervention.

#### 2.5.4. Setting

We included studies conducted in any setting providing mental health services or support, including community-based services (including voluntary and third sector services), outpatient services, emergency departments, inpatient services, primary care-based services, digital or telehealth-delivered services, educational settings, and forensic settings.

#### 2.5.5. Outcomes

We included quantitative outcomes (self-report, clinician-rated, or parent/carer-rated), if they were measured pre-and post-intervention, or at post-intervention. To be included, studies were required to report at least one of the following outcomes: mental health symptoms, safety (physical and psychological safety), wellbeing, social functioning and participation, loneliness or perceived social support/isolation, personal recovery, quality of life, housing security, financial security, employment outcomes, educational outcomes, use of restrictive practices and other containment measures (e.g., seclusion, physical restraint, rapid tranquillisations), or acute service use (e.g., presentations to acute services [e.g. emergency departments, crisis services], hospital admissions and detentions).

We excluded studies that did not report one of those listed mental health treatment outcomes – for example, studies which only reported a measure of autistic traits/behaviour.

#### 2.5.6. Study design

We included quantitative studies (e.g., cohort studies, case control studies, mixed-methods studies with a quantitative component, and randomised controlled trials (RCTs) where secondary analyses compared outcomes for autistic and non-autistic participants). Grey literature, including pre-prints, service evaluations, clinical audits, and PhD theses were included. To be eligible for inclusion, grey literature needed to include a methodology description.

We excluded studies using only qualitative methods, conference proceedings, abstracts without an associated full text, books, MSc/BSc theses, case studies and case series, opinion pieces, reviews, blog posts, social media content, and grey literature without a clear methodology description.

### 2.6. Data extraction

A data extraction sheet was designed in Microsoft Excel and revised based on feedback from the working group and piloting on a small sample of eligible papers by JG, JT, and FE. The final data extraction sheet template can be seen in Additional File 2. Data extraction was conducted by six researchers (JG, FE, JT, KH, MC, SL). Data were independently double extracted for all included papers, and study authors were contacted for missing information where relevant. Any discrepancies were resolved through discussion between JG, JT, FE, SL.

### 2.7. Quality appraisal

#### 2.7.1. Critical Appraisal Skills Programme checklist for cohort studies

All papers were appraised using a modified version of the Critical Appraisal Skills Programme (CASP) checklist for cohort studies (44). Adaptation involved expanding the original yes/no/can’t tell scoring to a four-point scale (yes/partially/no/can’t tell) to allow for more sensitive appraisal. For each item, our team agreed on specific criteria for assigning each rating, including considerations relevant to autism-related research, to improve consistency across raters. The modified CASP checklists are available in Additional File 3.

Two members of the research team (JG, FE, JT, KH, MC, SL) rated each study independently. Any disagreements were resolved through discussion with a third researcher or the wider working group. Scores for the individual question items (‘no’ or ‘can’t tell’ = 0; ‘partially’ = 1; ‘yes’ = 2) were combined to give a total overall score for each study. Studies scoring under 50% of the total possible score were classified as low quality; 50-74% as medium quality; and 75%+ as high quality. Whilst we did not exclude papers based on their quality, this was taken into consideration during interpretation of the findings.

#### 2.7.2. Autism-Inclusive Research Assessment (AIRA) Ratings

We assessed autism-inclusive research practices in the included studies using a modified version of the Autism-Inclusive Research Assessment (AIRA). This tool was developed by the MHPRU team, led by RRO and other lived experience researcher colleagues who are or suspect they are autistic, for use in two previous systematic reviews to assess the autism-inclusiveness of designs and methods in studies (45,46). The AIRA was developed in response to limitations in existing quality appraisal tools when applied to autism research in mental health, where there is a history of contested assumptions, including the use of neurotypical norms in outcome measures and deficit-focused models of autism. It was designed to support the explicit and systematic incorporation of these autism-specific considerations within systematic review quality assessment.

For this review, we further developed the AIRA to expand the domains included in the original version of the AIRA. The adaptation was led by lived experience researchers within the working group, with feedback from the wider working group. The revised AIRA considers whether autistic people were involved in designing the research and in shaping outcome selection and measurement; whether outcome measures were adapted for, or validated with, autistic populations; and whether any outcome measures risk conflating autistic traits and mental health symptoms. It also examines whether adjustments were made to data collection tools or procedures to support wide participation; whether the intervention encouraged masking of autistic traits/behaviours; whether the authors use language that fits with a neurodiversity-affirming approach throughout (informed by Botha & Cage (2022) (47); and the extent to which the research reflected priorities and values identified by autistic communities, drawing on UK-based work (34,48). For each included study, each AIRA domain was rated ‘yes’, ‘partially’, ‘no’, ‘can’t tell’, or ‘not applicable’ (N/A). The revised AIRA tool is available in Additional File 3. Double independent AIRA ratings were made for each study by two members of the research team, including at least one autistic researcher. Any disagreements were resolved through discussion between the researchers or with the wider working group.

### 2.8. Evidence synthesis

Evidence synthesis was led by JG and FE, supported by the working group. Characteristics of included studies were tabulated and described narratively. Quantitative outcome data were tabulated and summarised narratively (49). This included reporting original measures of effect (e.g., mean differences or standardised mean differences [SMD] for continuous outcomes, and odds ratios, risk ratios, or risk differences for dichotomous outcomes) and p-values, where available. Results were grouped according to service/intervention type. There were no minimum number of studies or required level of agreement across studies for data synthesis.

Meta-analysis was undertaken only when there were at least two studies judged to be sufficiently comparable in terms of population, intervention, setting, and outcomes. The ‘metafor’ package (50) in R (version 4.5.3) was used to calculate the SMD between groups at end-of-treatment and pooled using random-effects models, correcting for small sample sizes (Hedges’ *g*), with 95% confidence intervals. Effect sizes were considered significant if *p* < 0.05, and were tentatively interpreted as small (0.2), medium (0.5), and large (0.8) (51). Heterogeneity was assessed using Higgins’ *I^2^* (25% = low, 50% = moderate, 75% = high) (52).

## 3. Results

### 3.1. Characteristics of included studies

Figure 1 presents the PRISMA flow diagram of the screening and selection process. We identified 39 studies for inclusion. Excluded full texts and their reasons for exclusion can be seen in Appendix D [see Additional File 1]. Studies were most commonly conducted in the UK (n = 18; 46.2%). Outside the UK, single-country studies were conducted in the USA (n = 2), Japan (n = 5), Australia (n = 3), Italy (n = 2), the Netherlands (n = 2), Sweden (n = 2), Denmark (n = 1), Spain (n = 1), and Korea (n = 1). One study involved multiple countries (Sweden, Denmark, and Norway) (53) and one study did not report its location.

**Figure 1.**
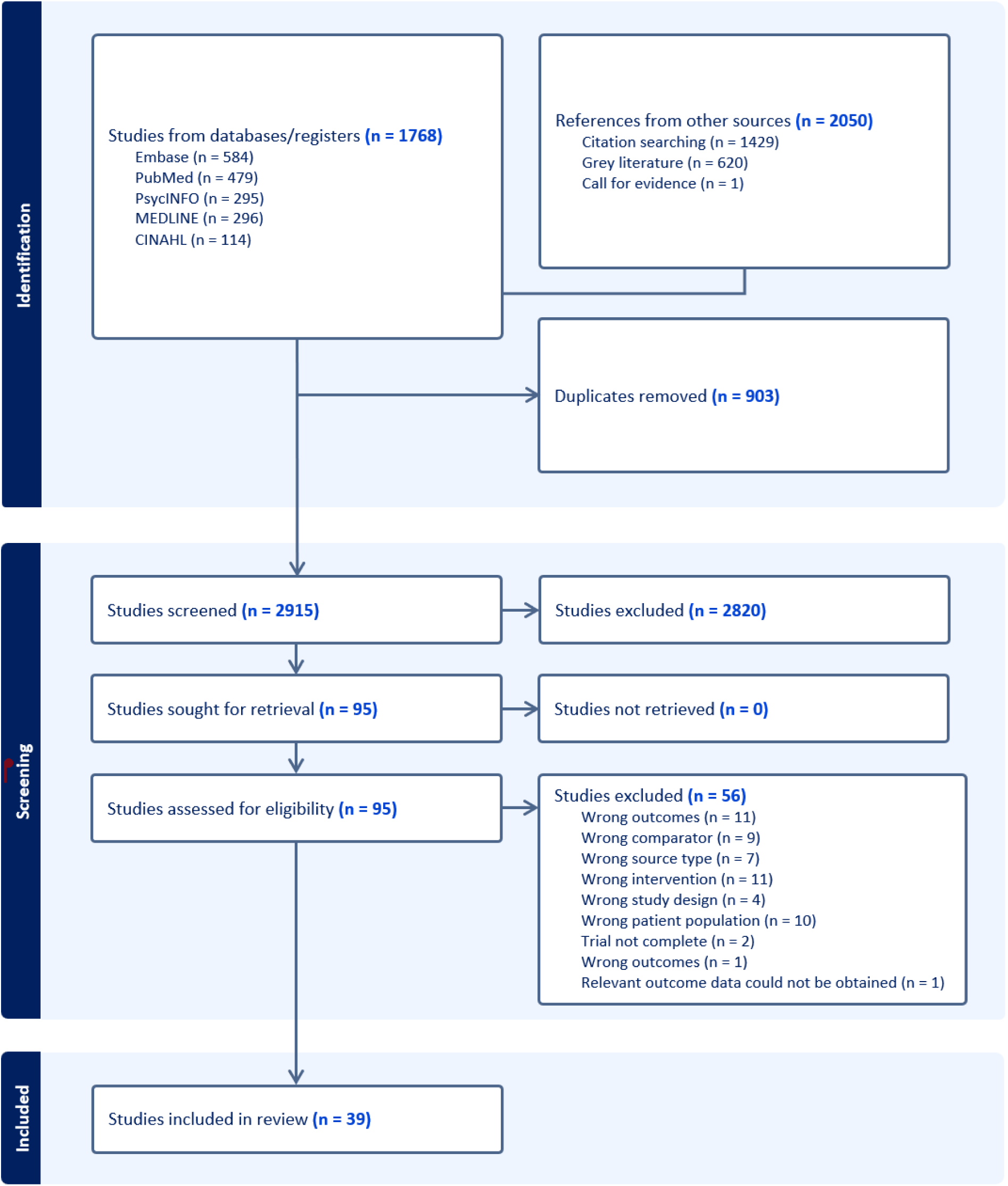
PRISMA flow diagram

All studies had cohort designs and were published in English. Fifteen studies (38.5%) included adults only (aged 18+), fifteen (38.5%) children and young people (CYP) only, and nine (23.1%) both adults and CYP. Only nine studies (23.1%) had samples including people with intellectual disabilities. The remainder either reported that their sample did not include people with intellectual disabilities (n = 18; 46.2%) or did not report this information (n = 12; 30.8%).

Only two studies (5.1%) reported conducting power calculations (54,55). Power calculations estimate the number of participants needed to reliably detect differences between groups. One of the studies indicated that some analyses were powered to detect medium-to-large effects (with others only powered for large effects) (54), and the other reported that the study was underpowered (55).

Seventeen studies examined psychological interventions and twenty-two focused on multi-component or service-level approaches. Early psychosis care was the most frequently studied intervention (n = 10), followed by Cognitive Behavioural Therapy (CBT) for obsessive-compulsive disorder (OCD) (n = 7 studies), while most other interventions were assessed in only one or two studies each. The included studies evaluated interventions for OCD, anxiety disorders, mild to moderate anxiety and/or depression, eating disorders, psychotic disorders, and “emerging borderline personality disorder” (BPD). Some studies evaluated services for people with a range of mental health symptoms or conditions. Study characteristics are summarised in Table 1. A full list of outcome measures reported in this review, along with their corresponding references, is provided in Appendix E [see Additional File 1].

**Table 1:**
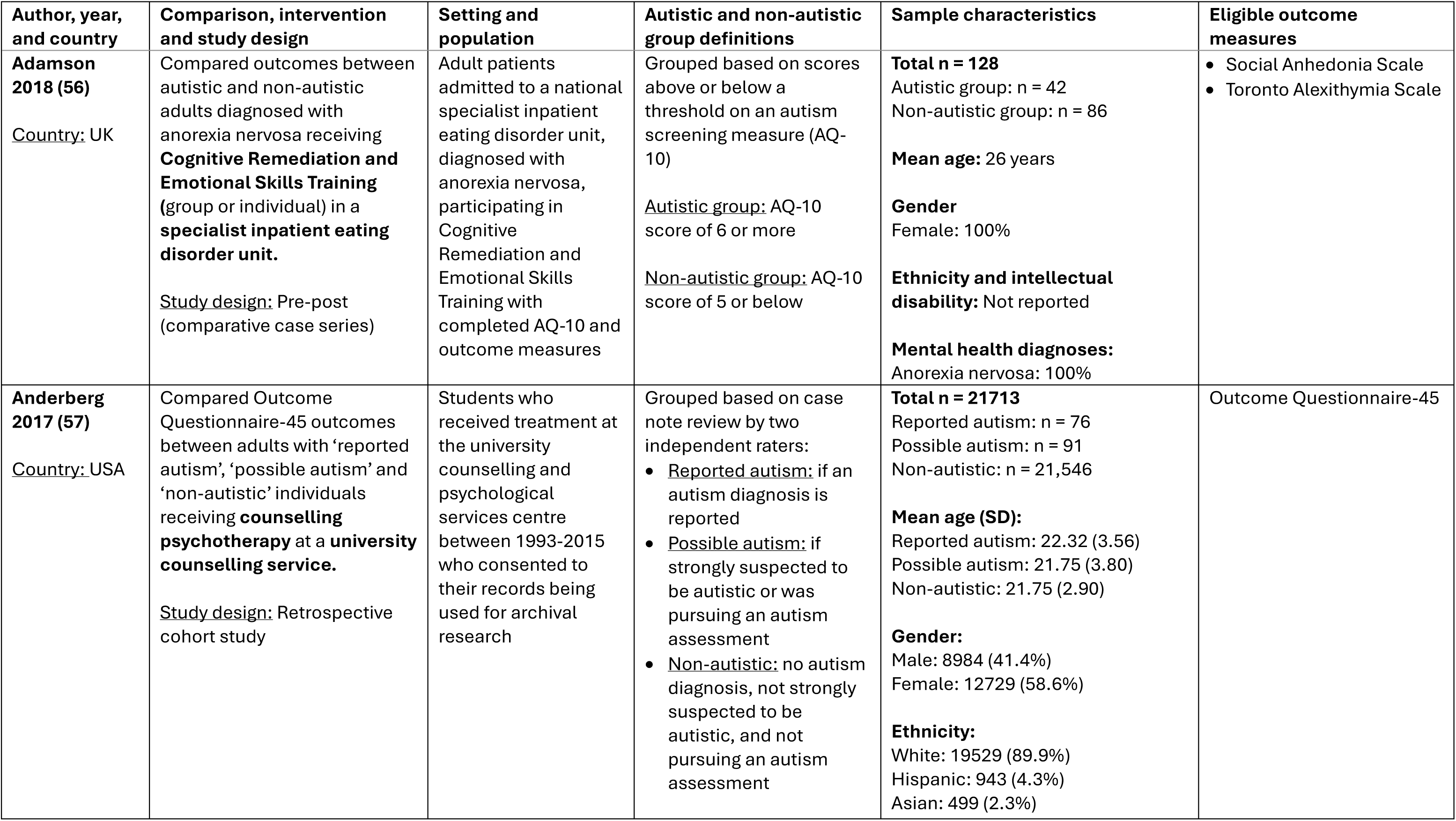
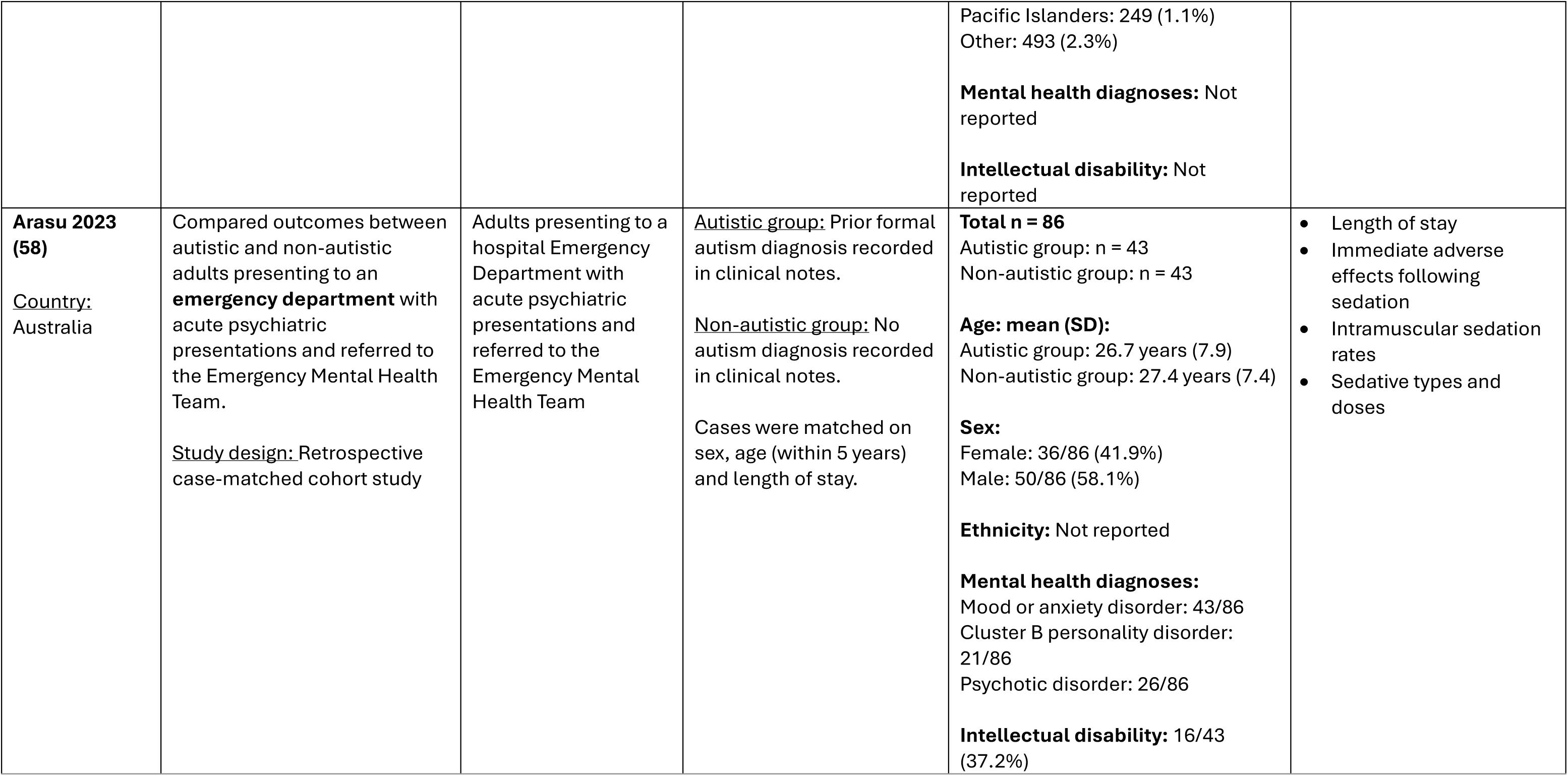
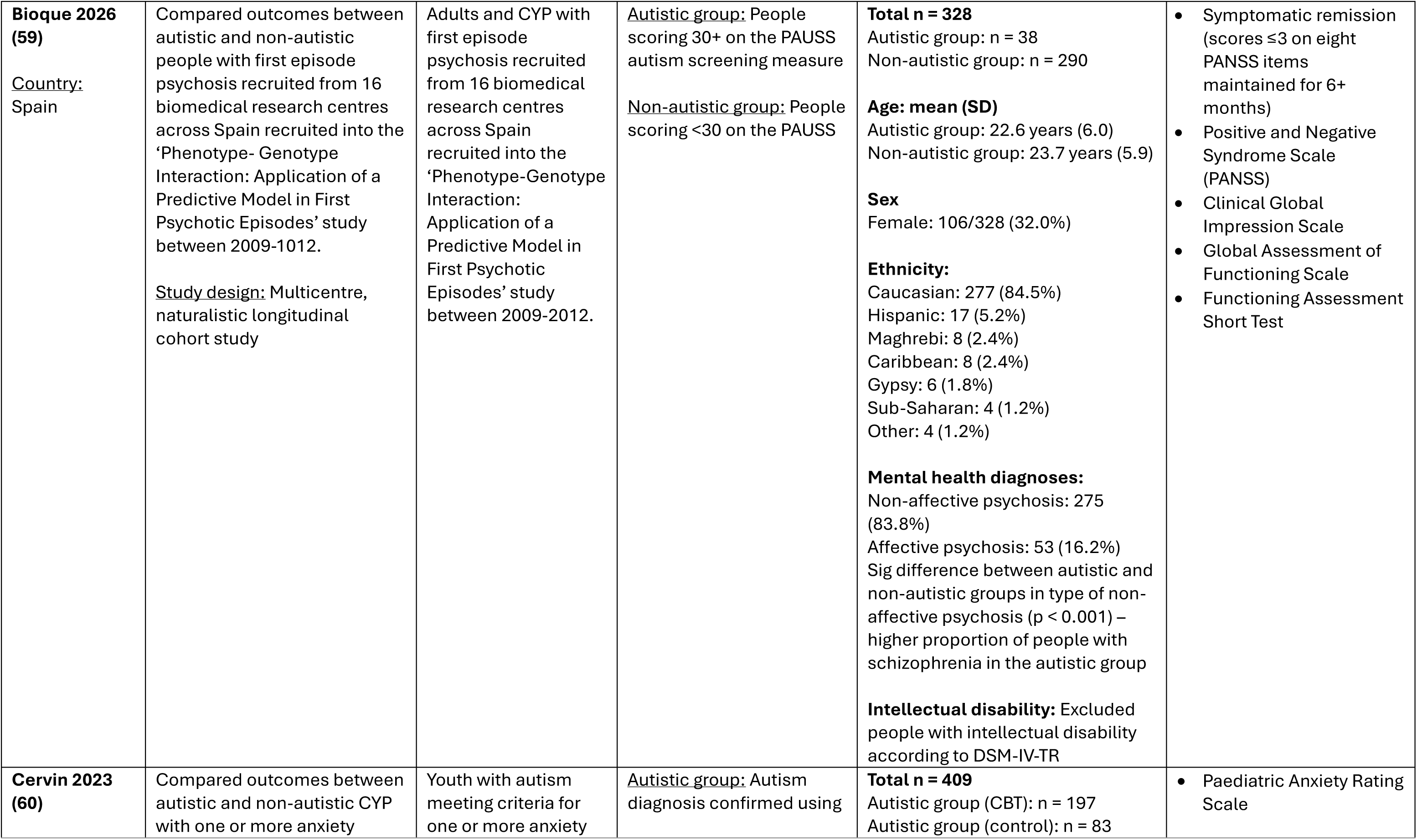
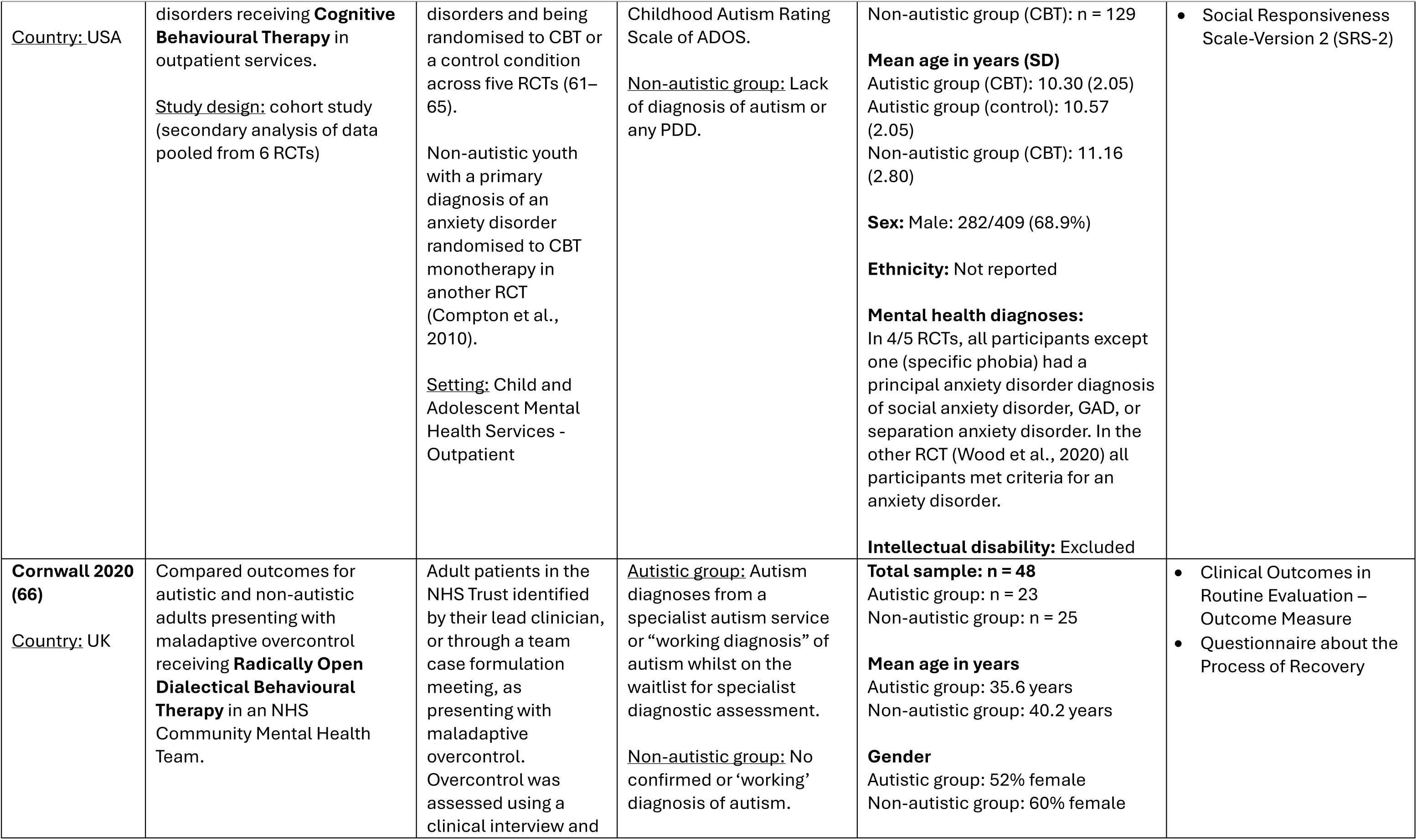
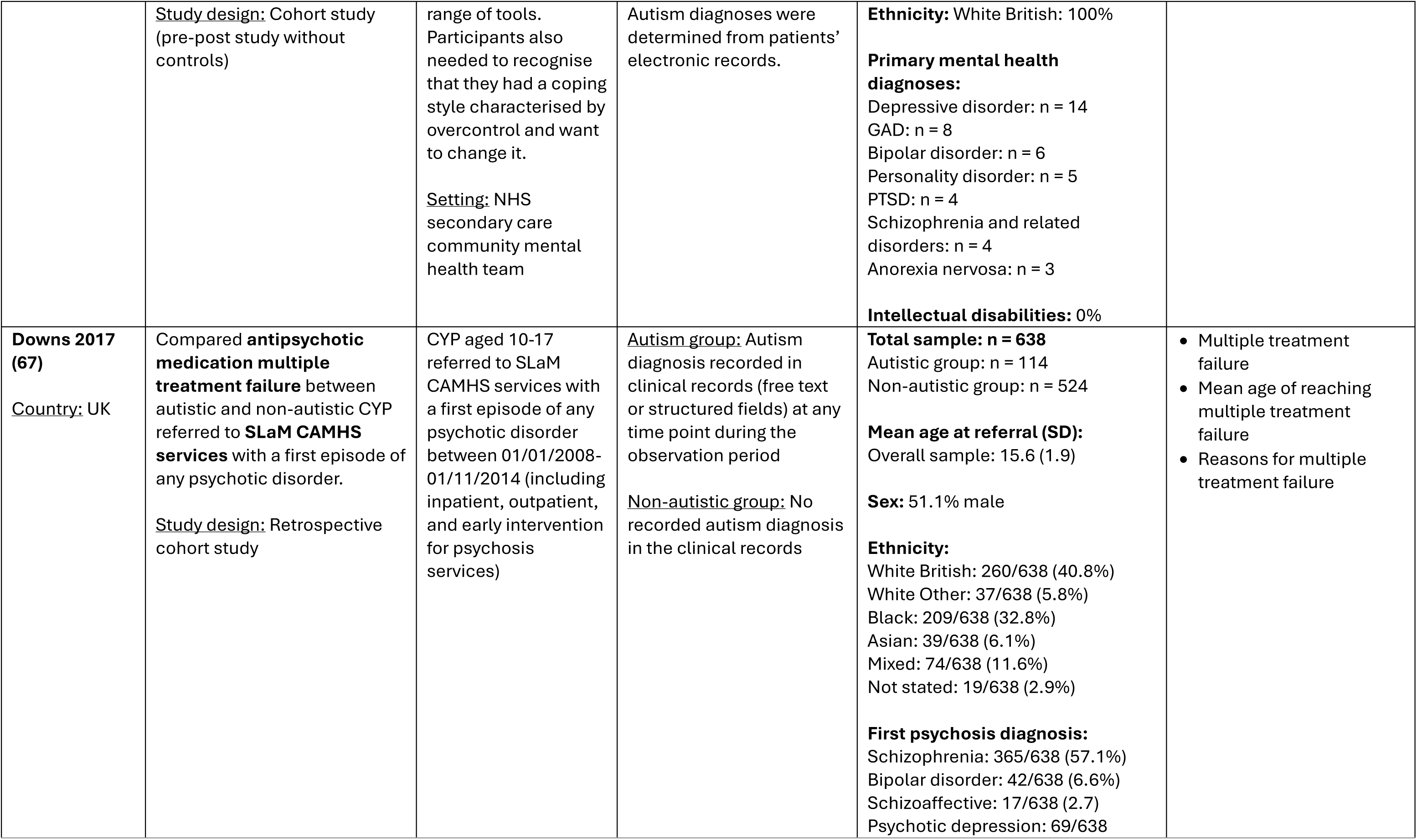
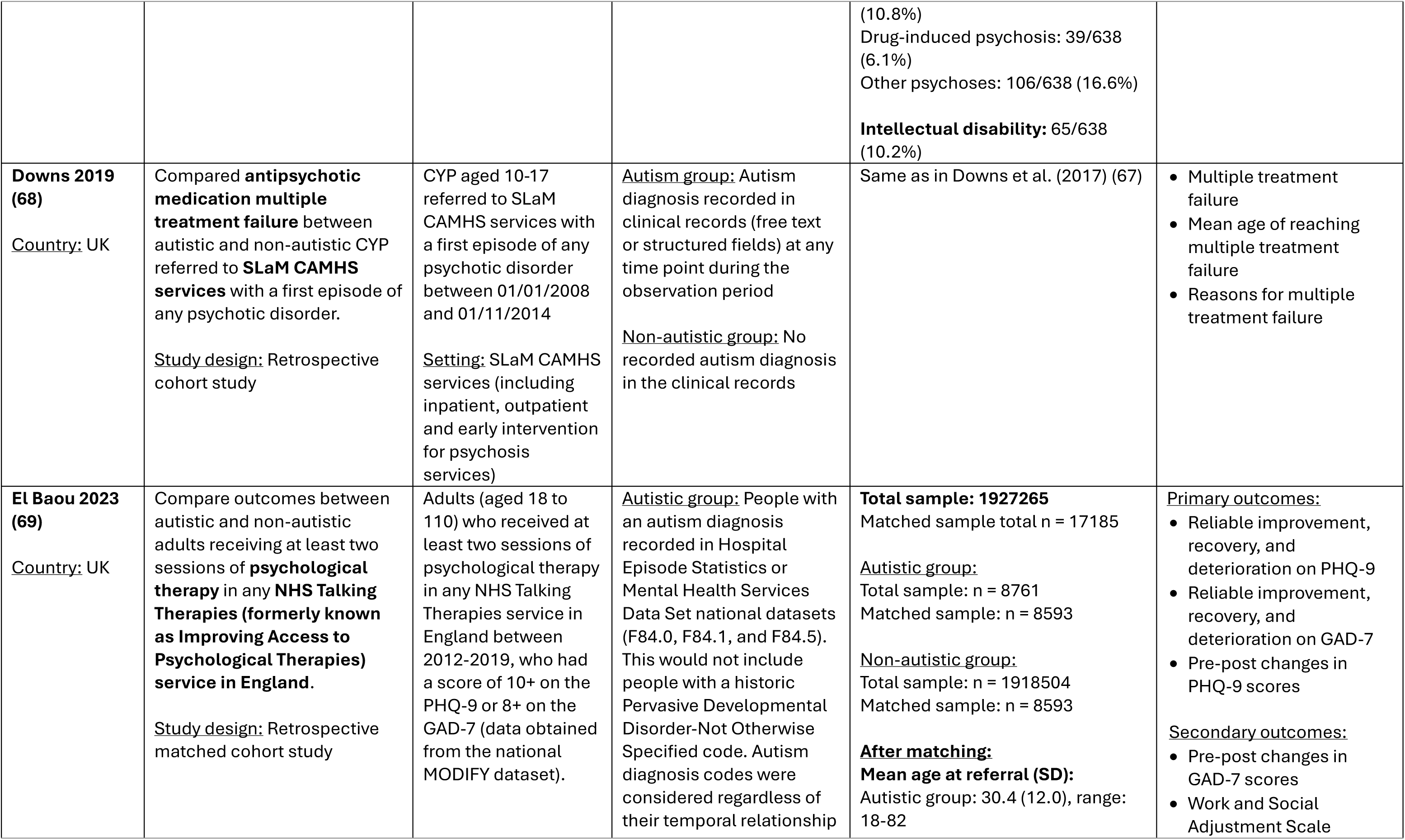
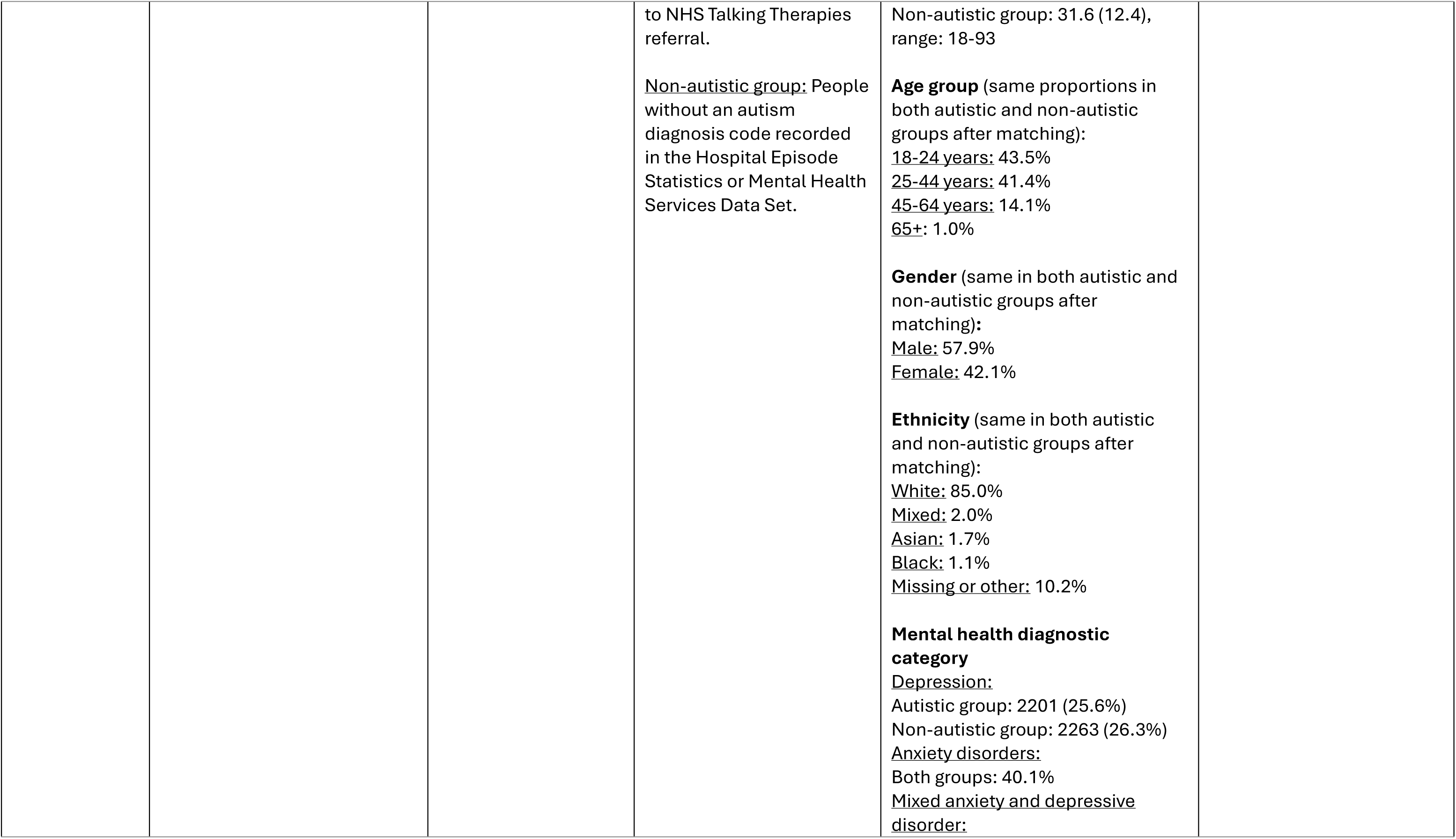
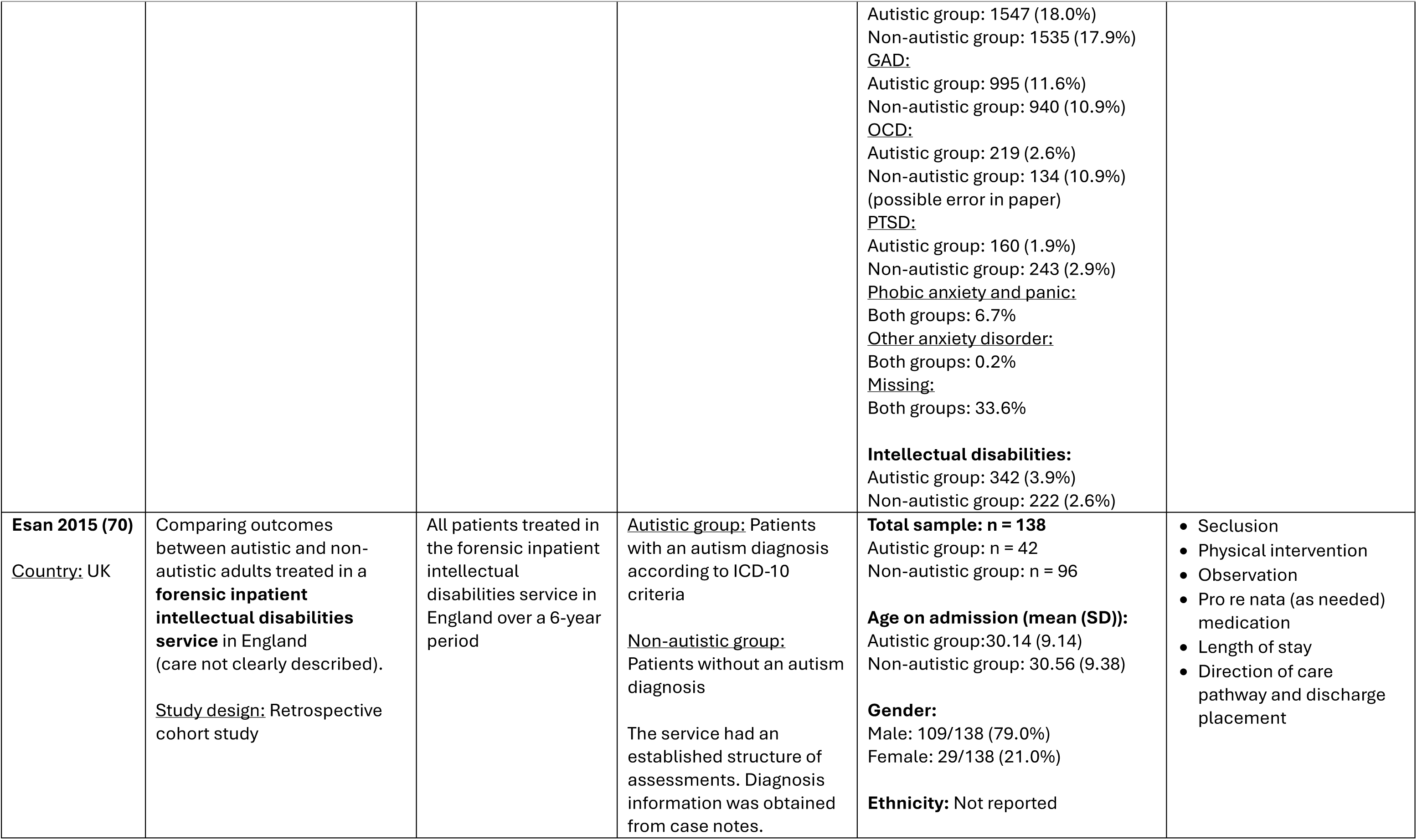
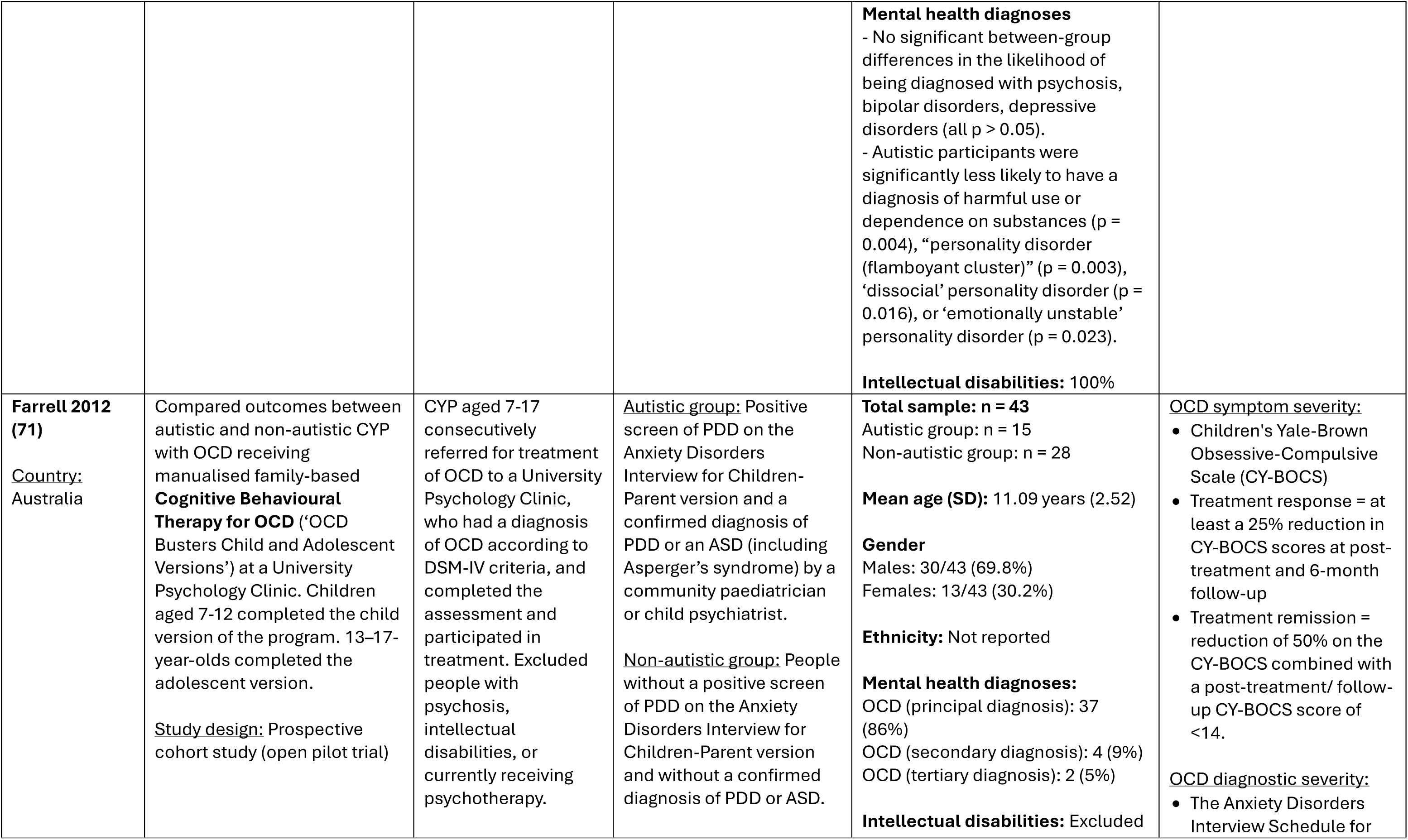
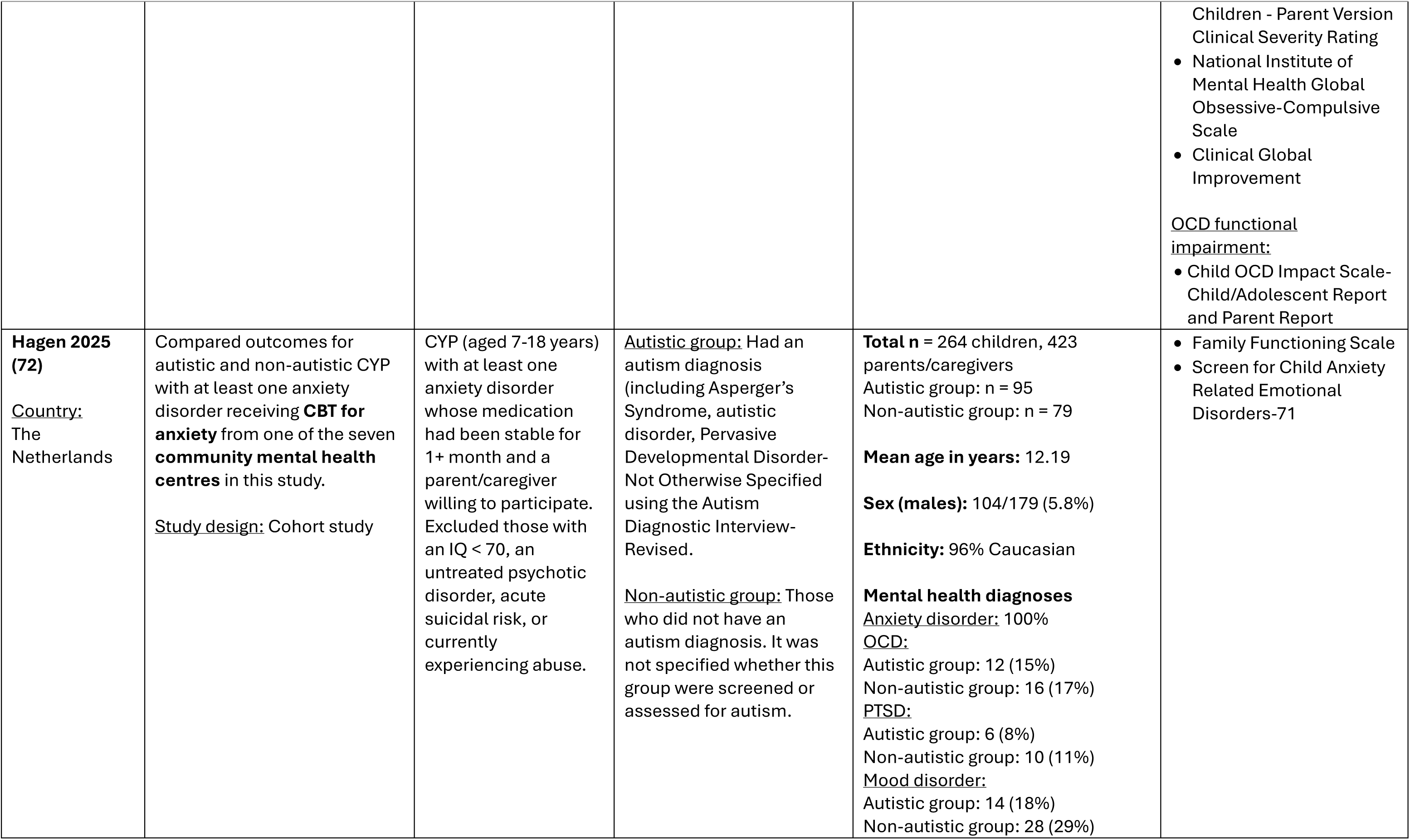
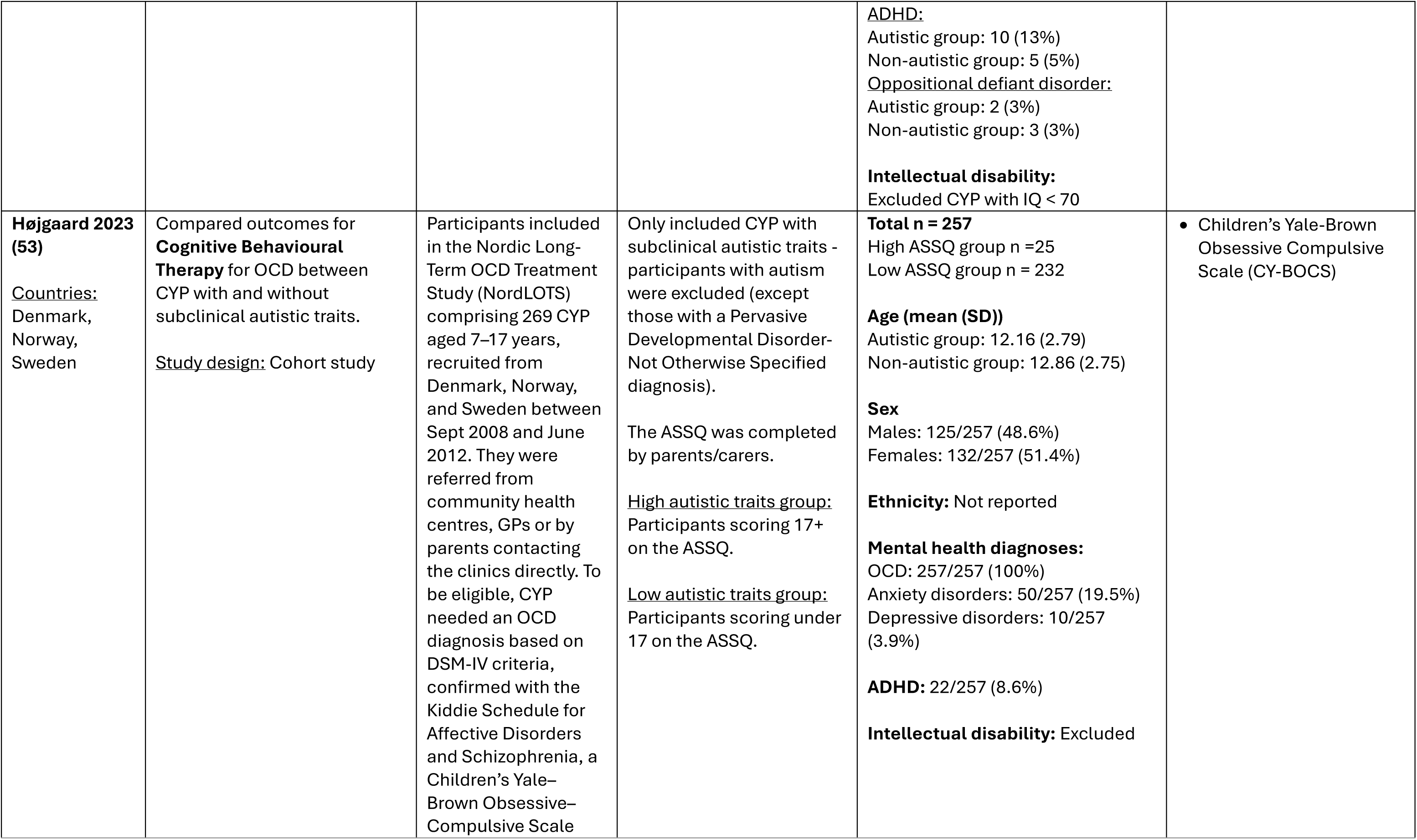
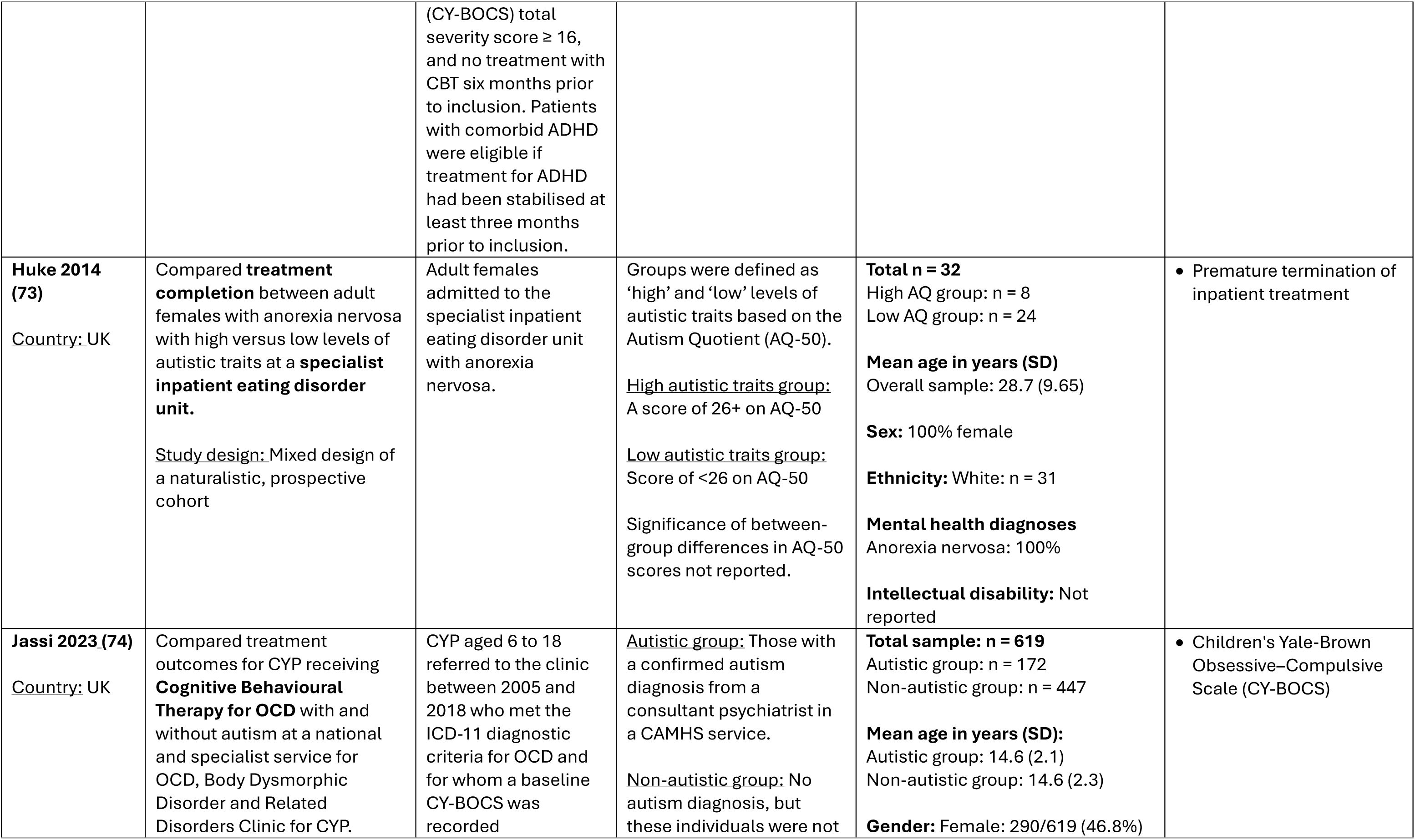
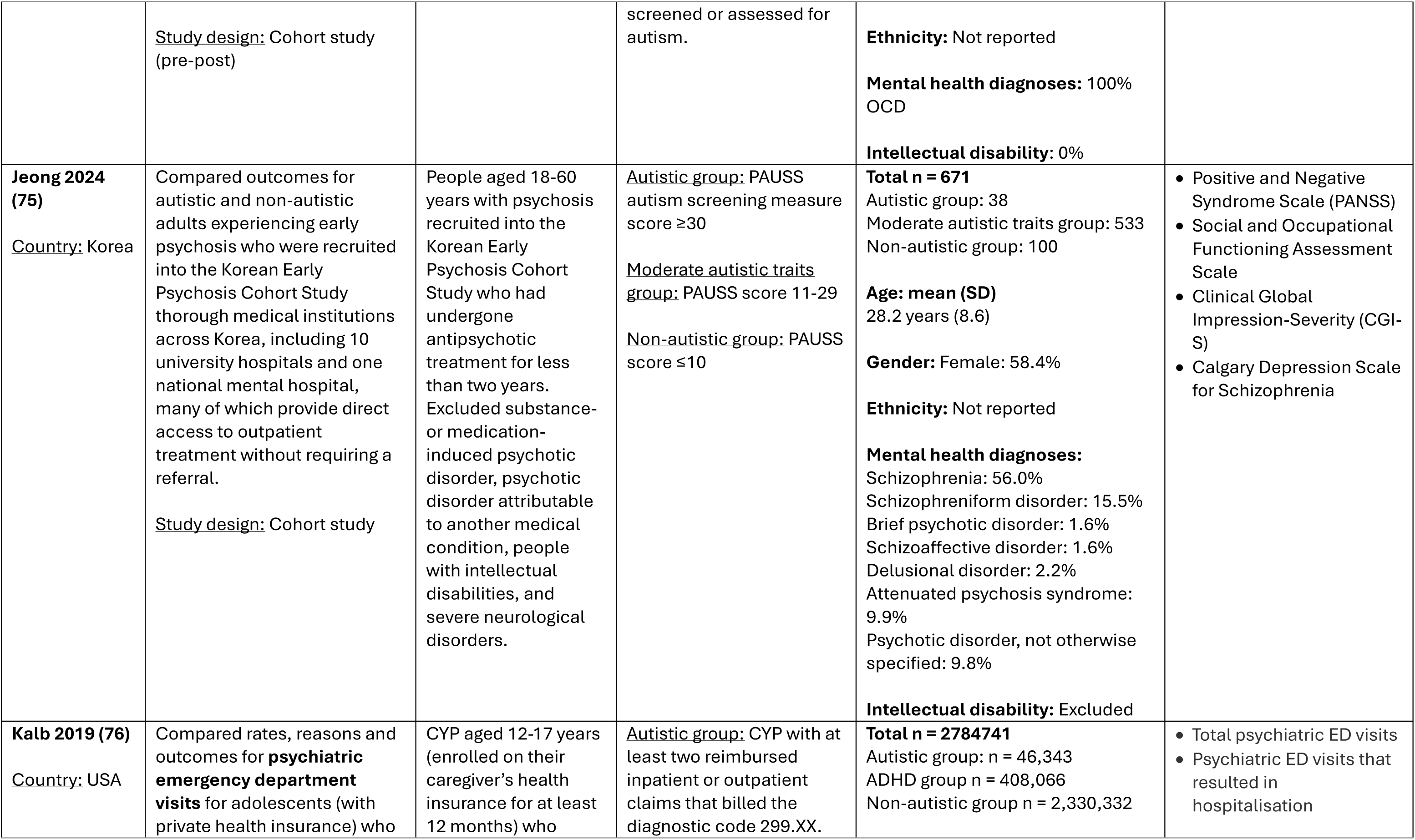
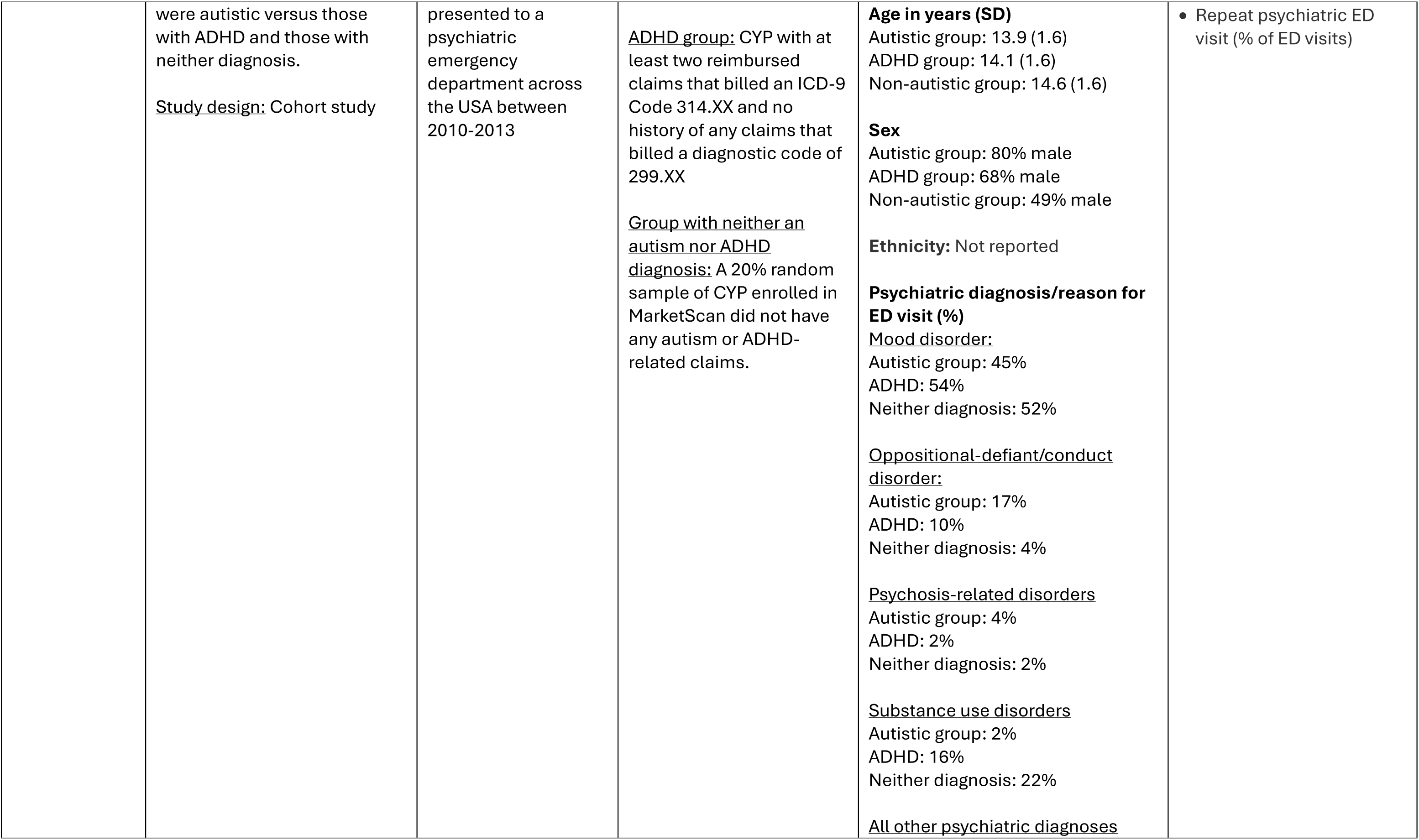
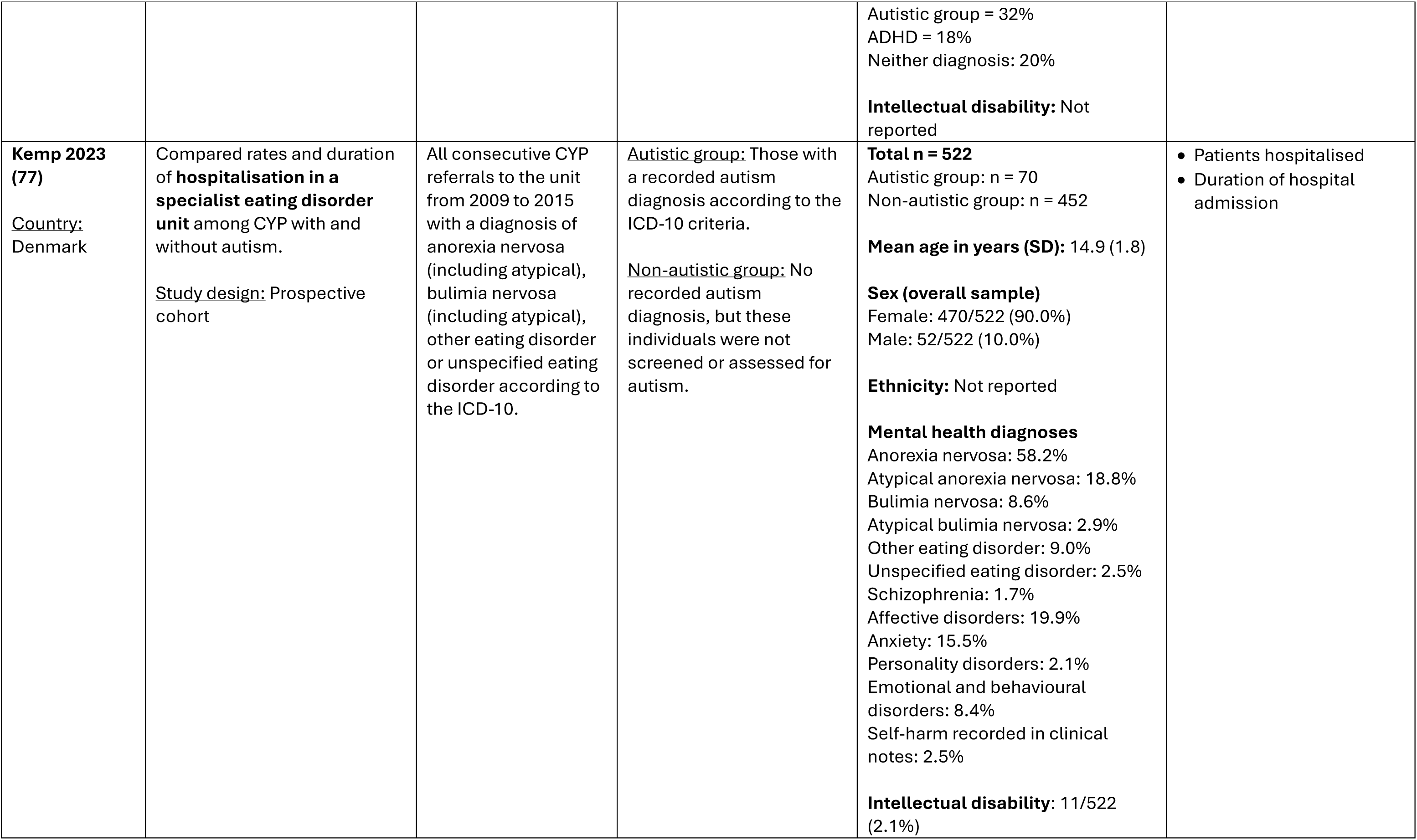
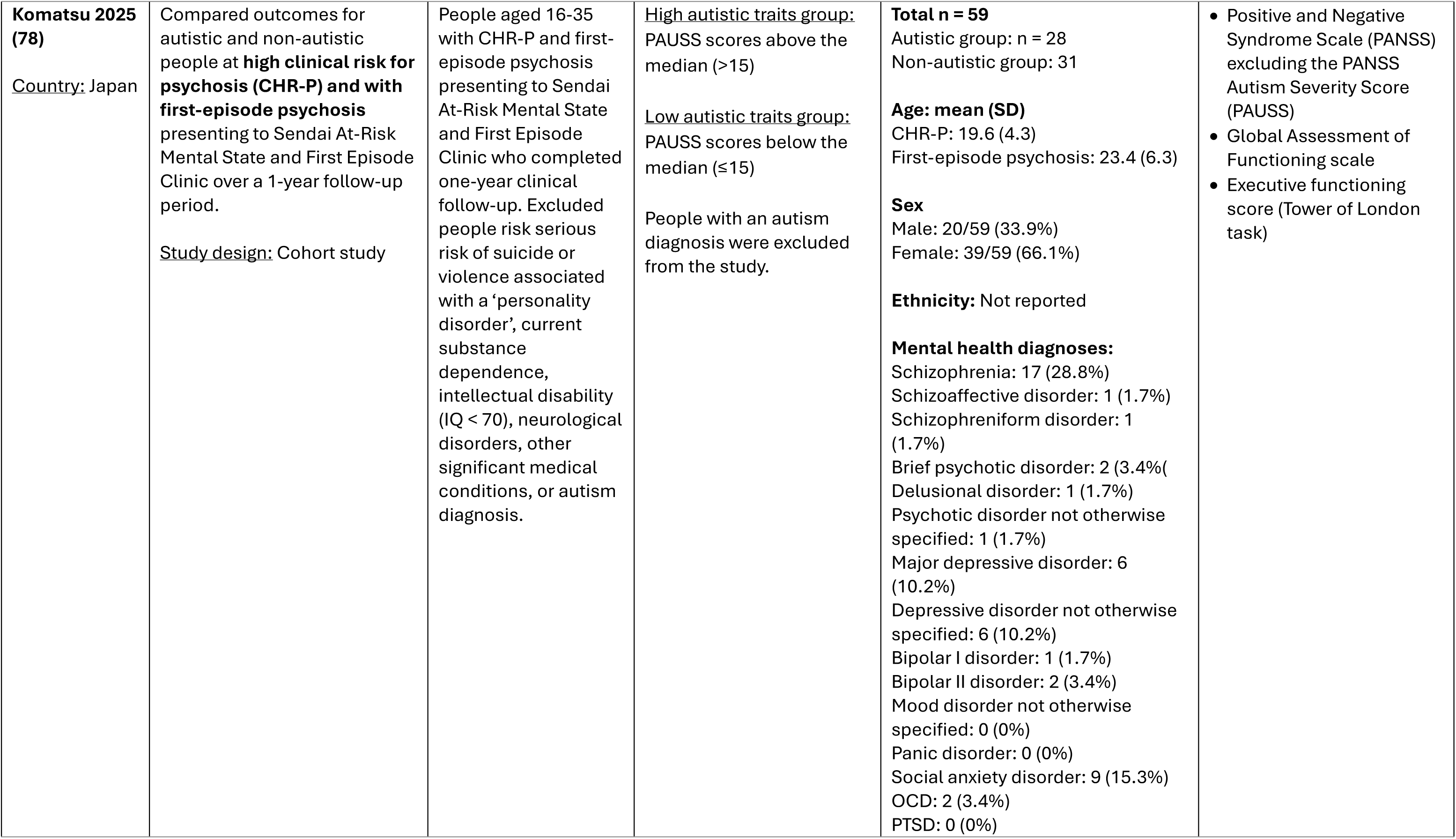
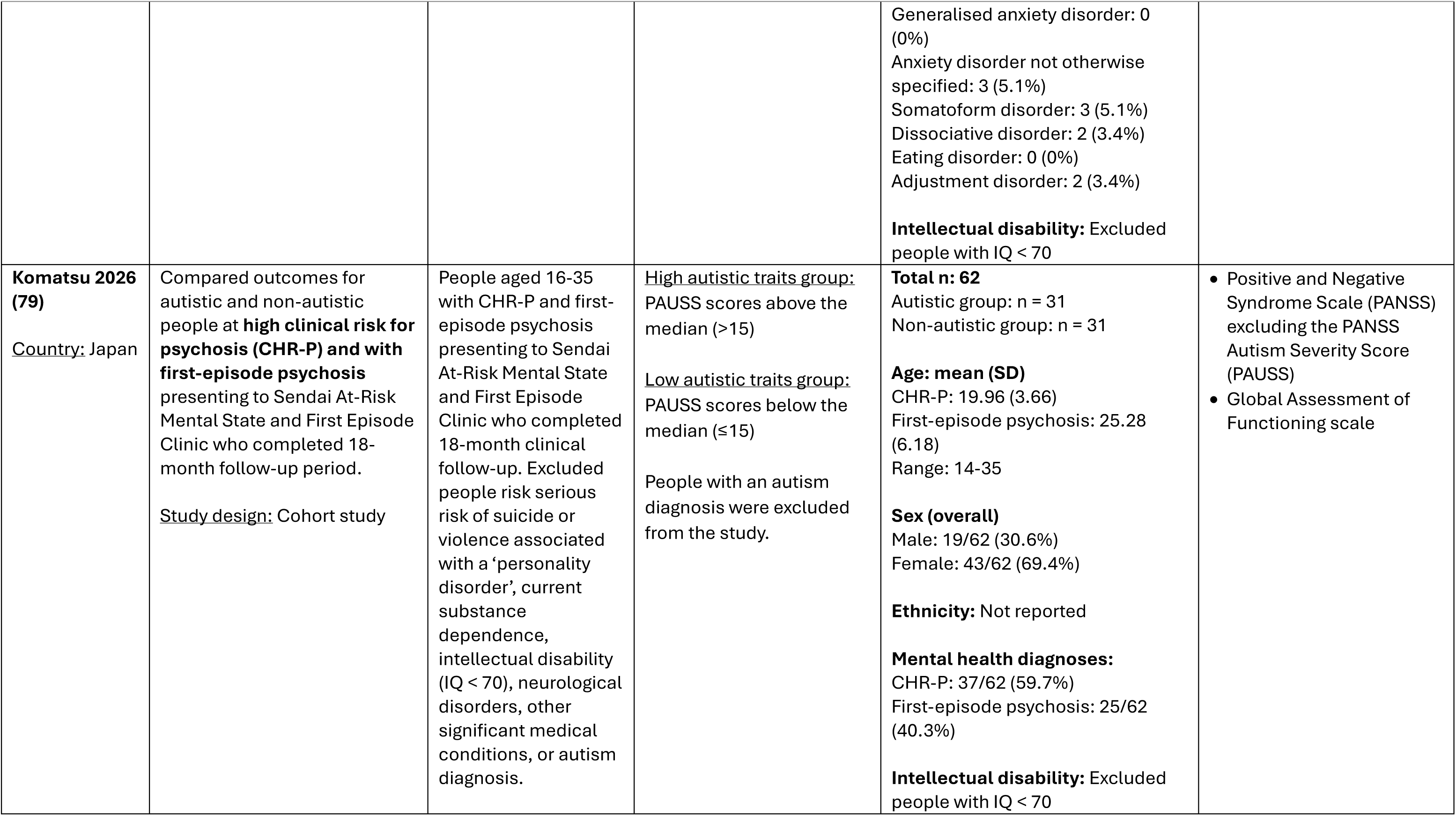
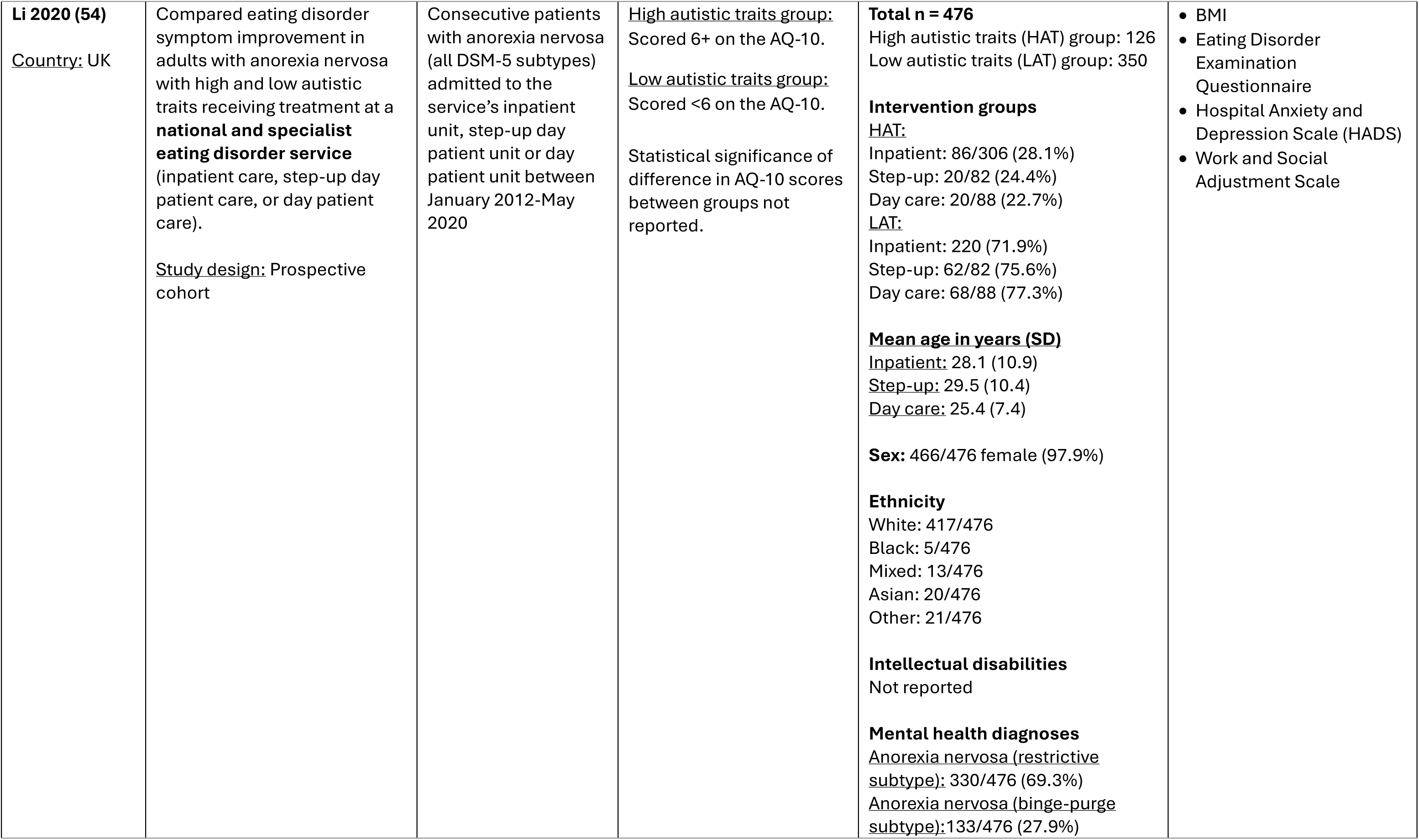
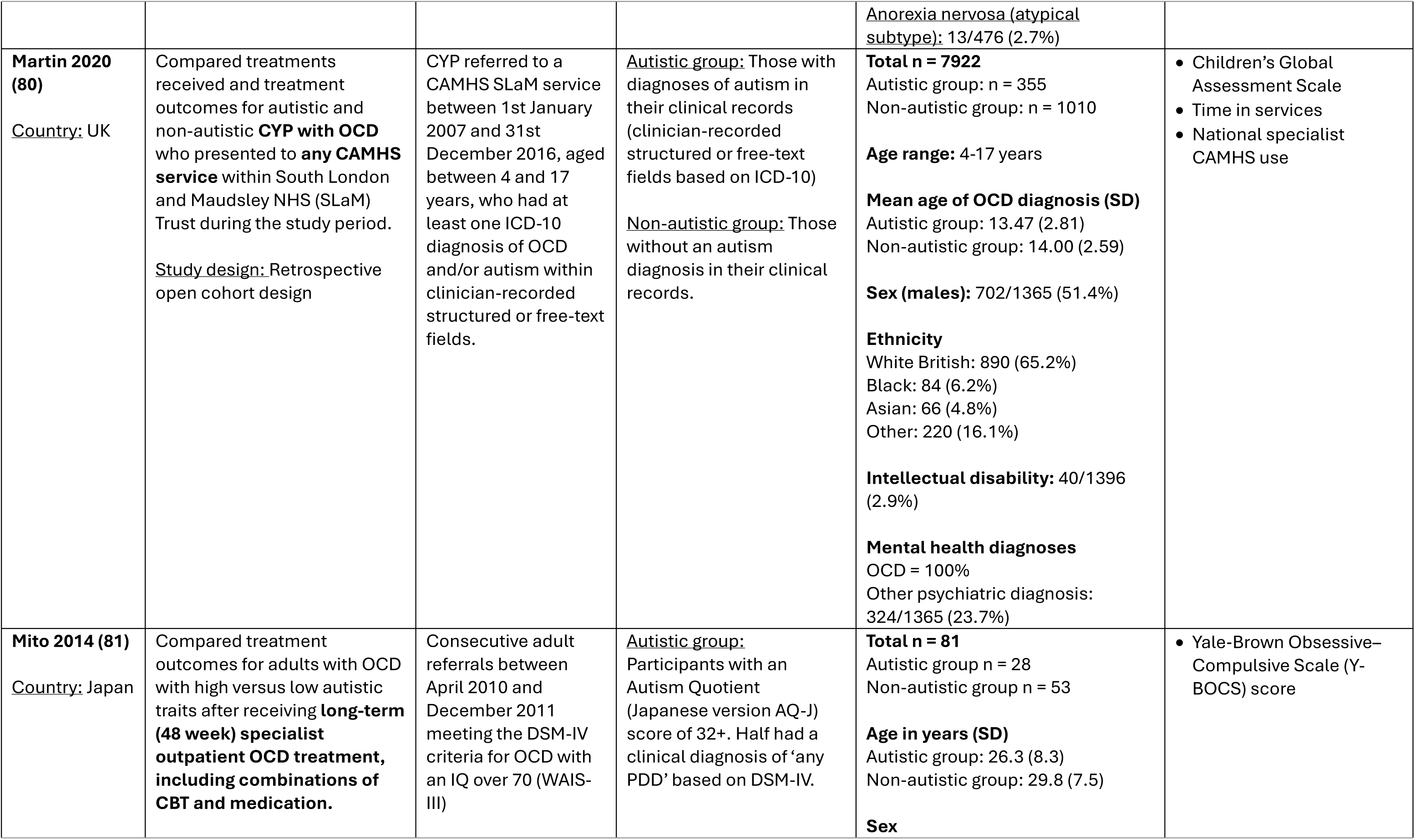
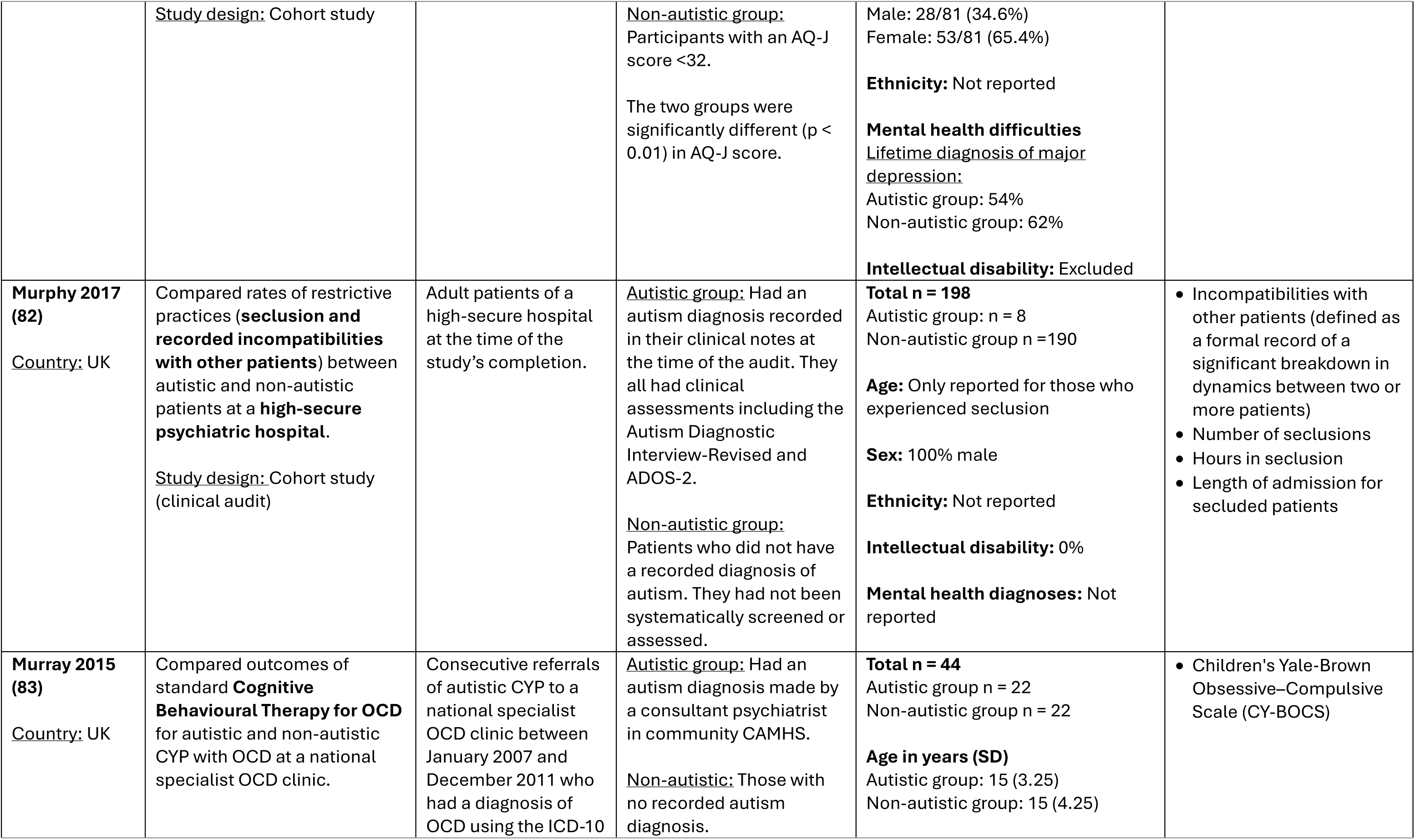
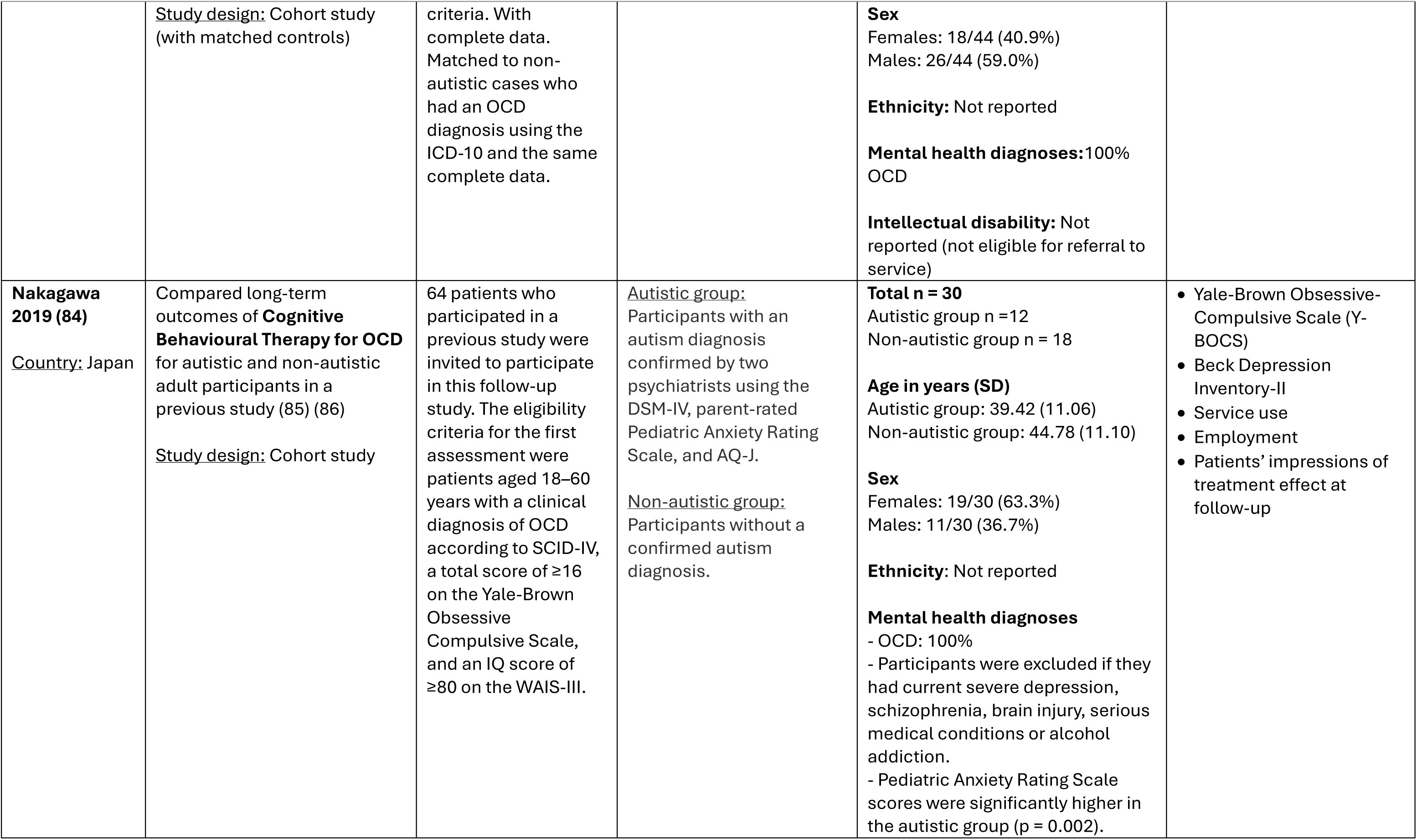
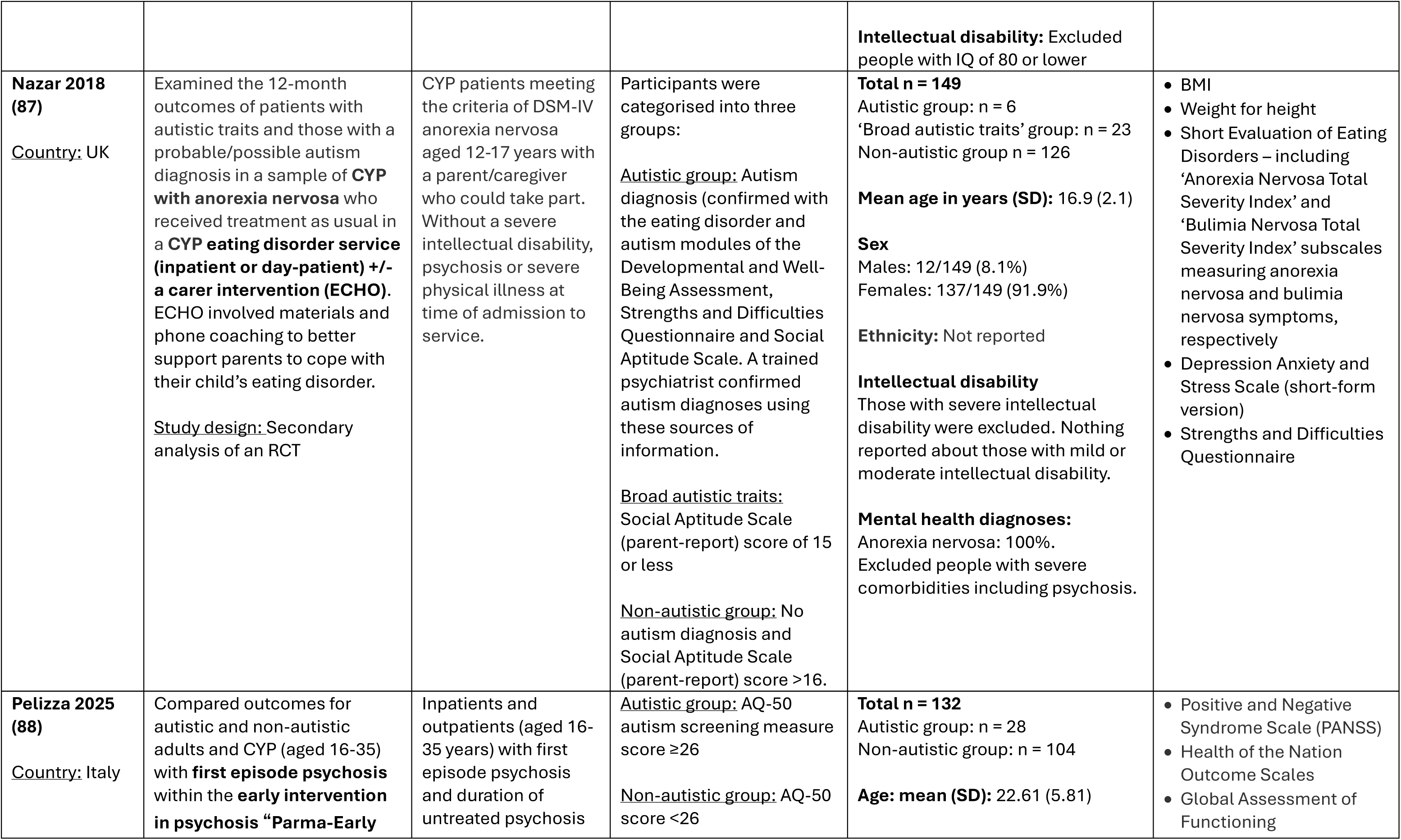
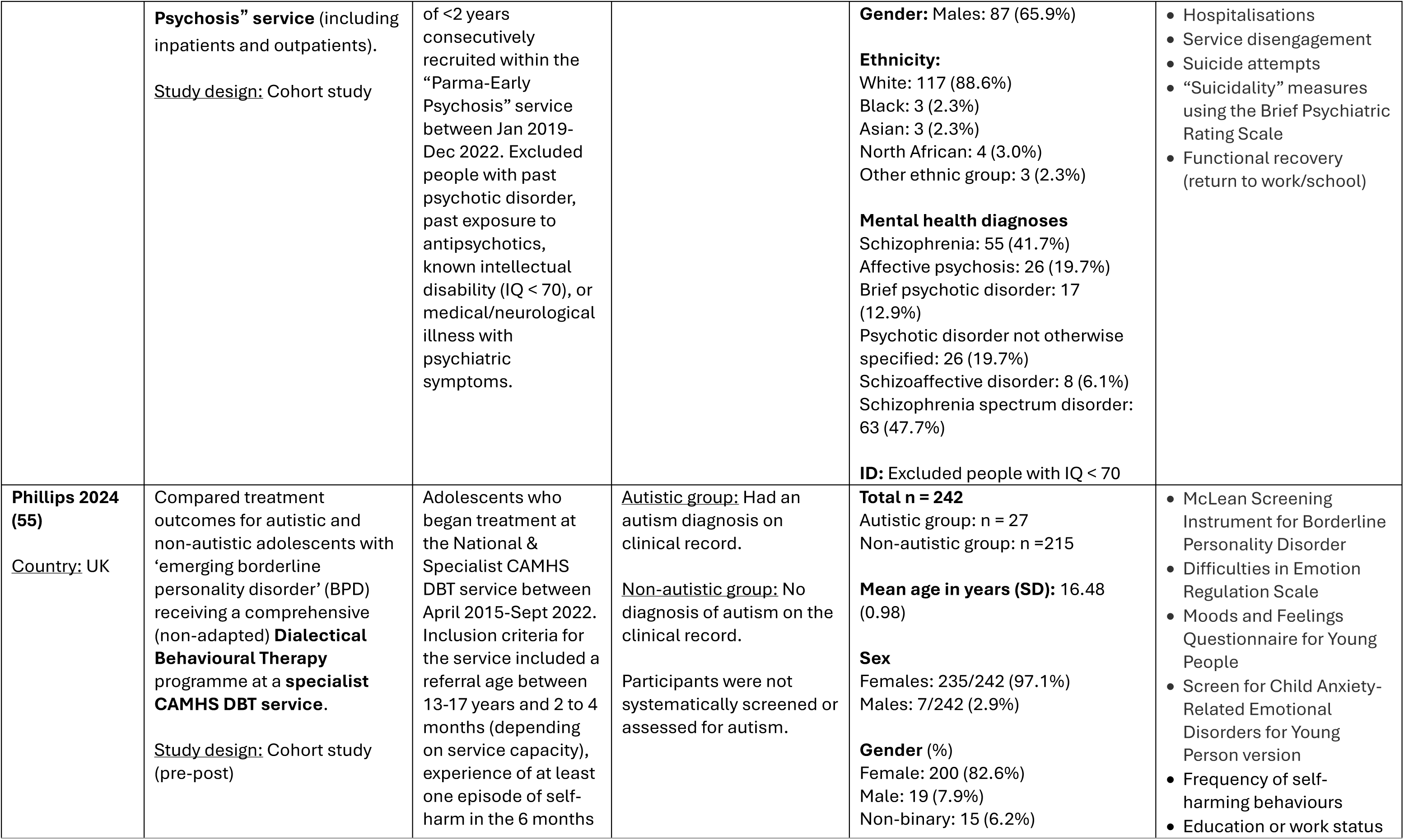
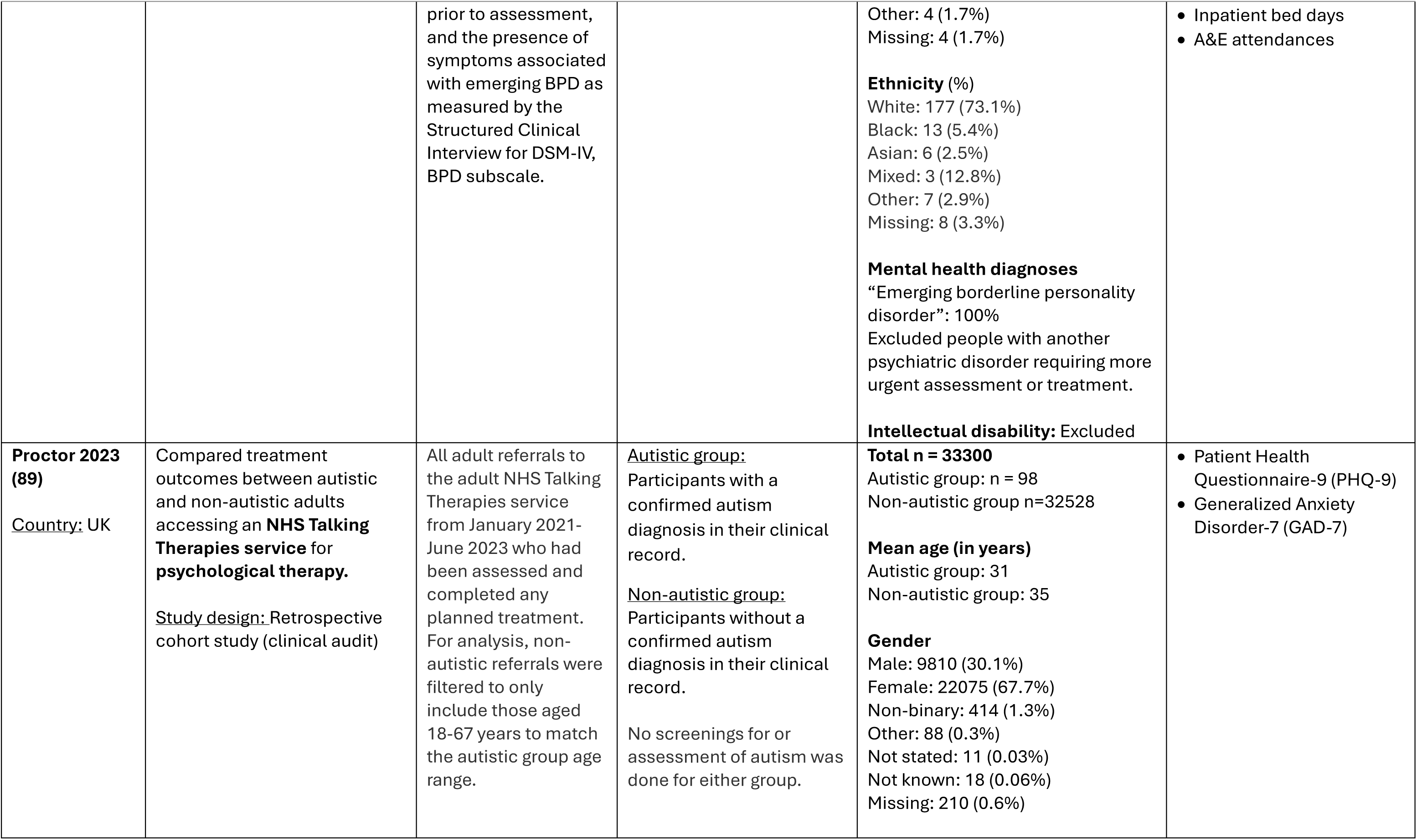
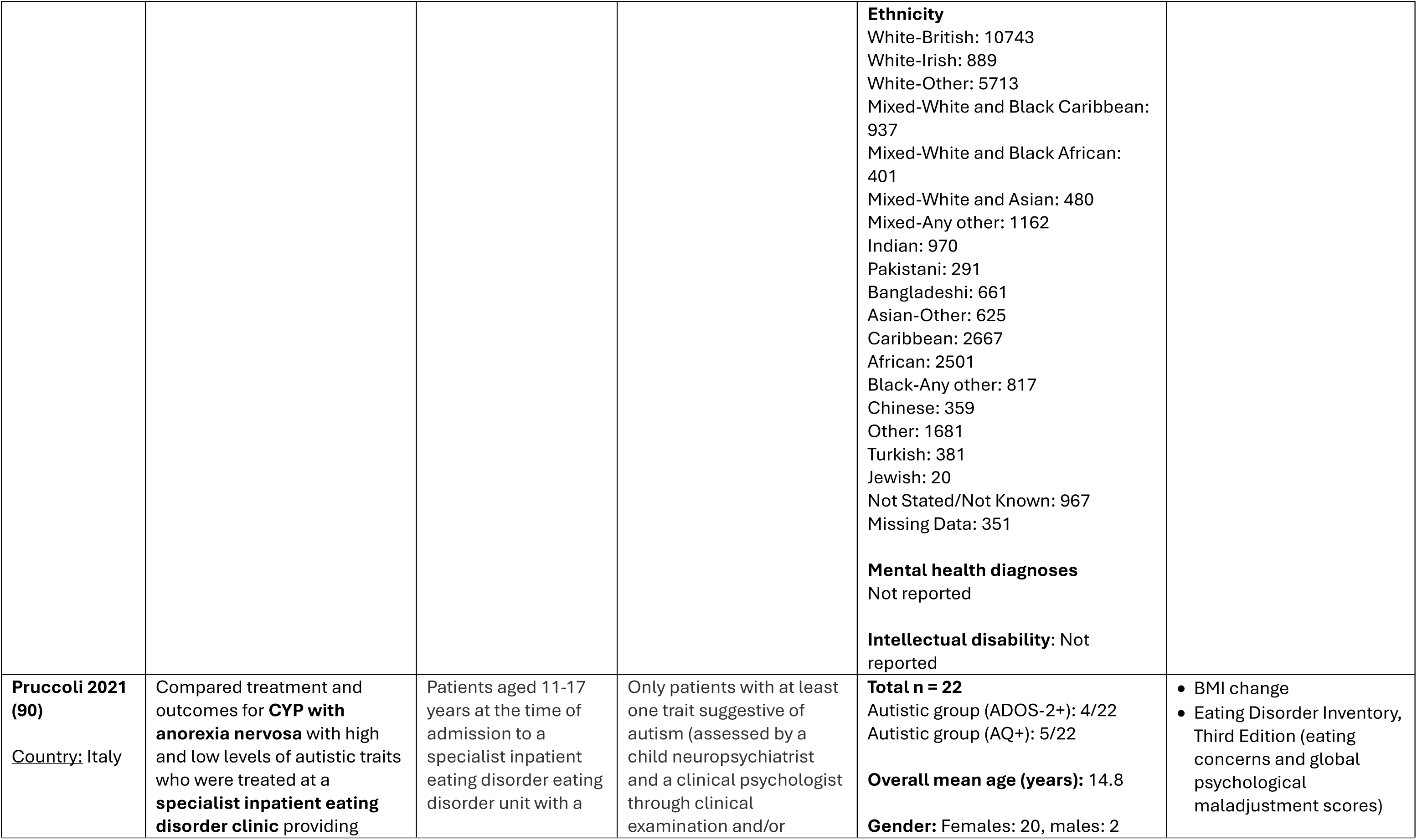
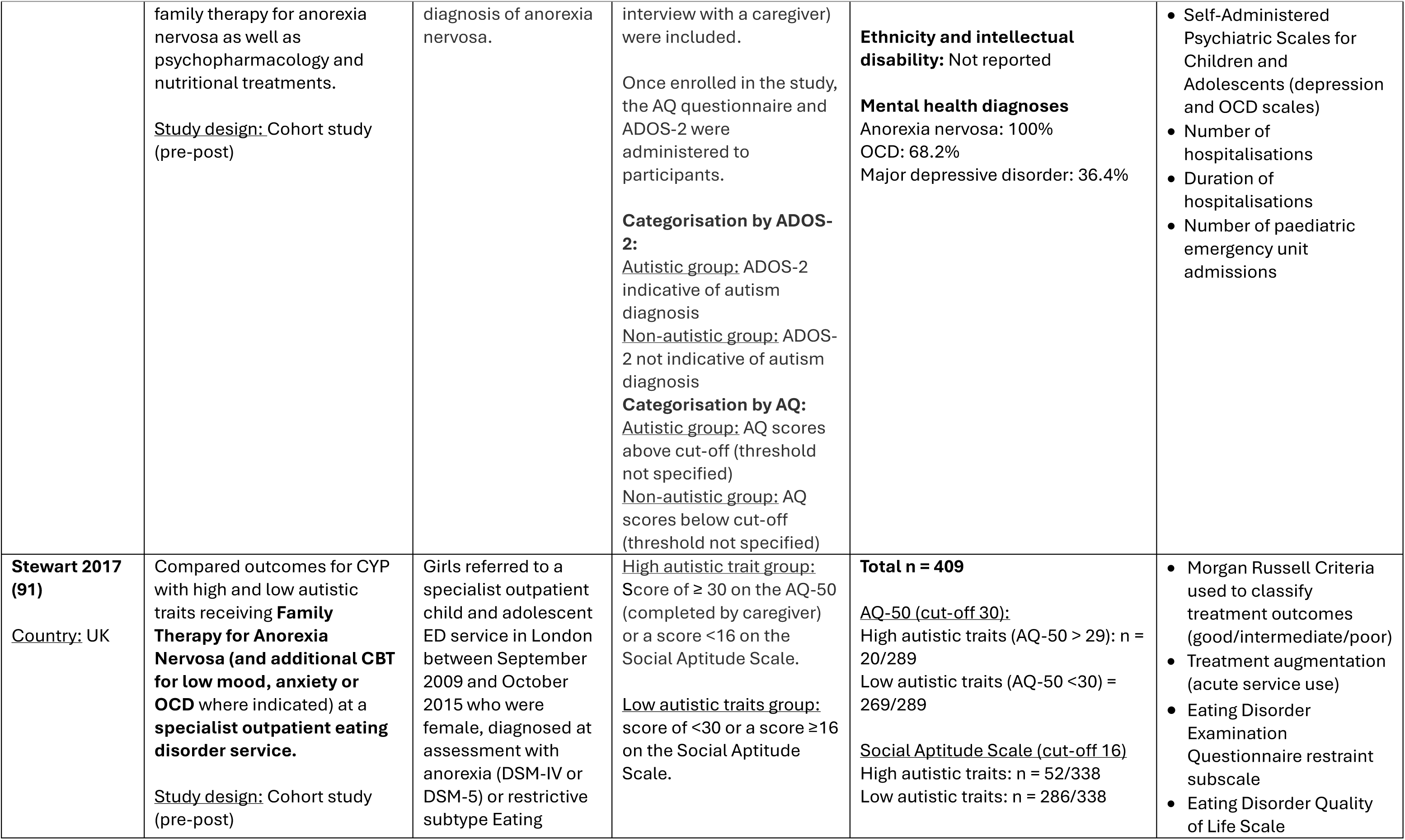
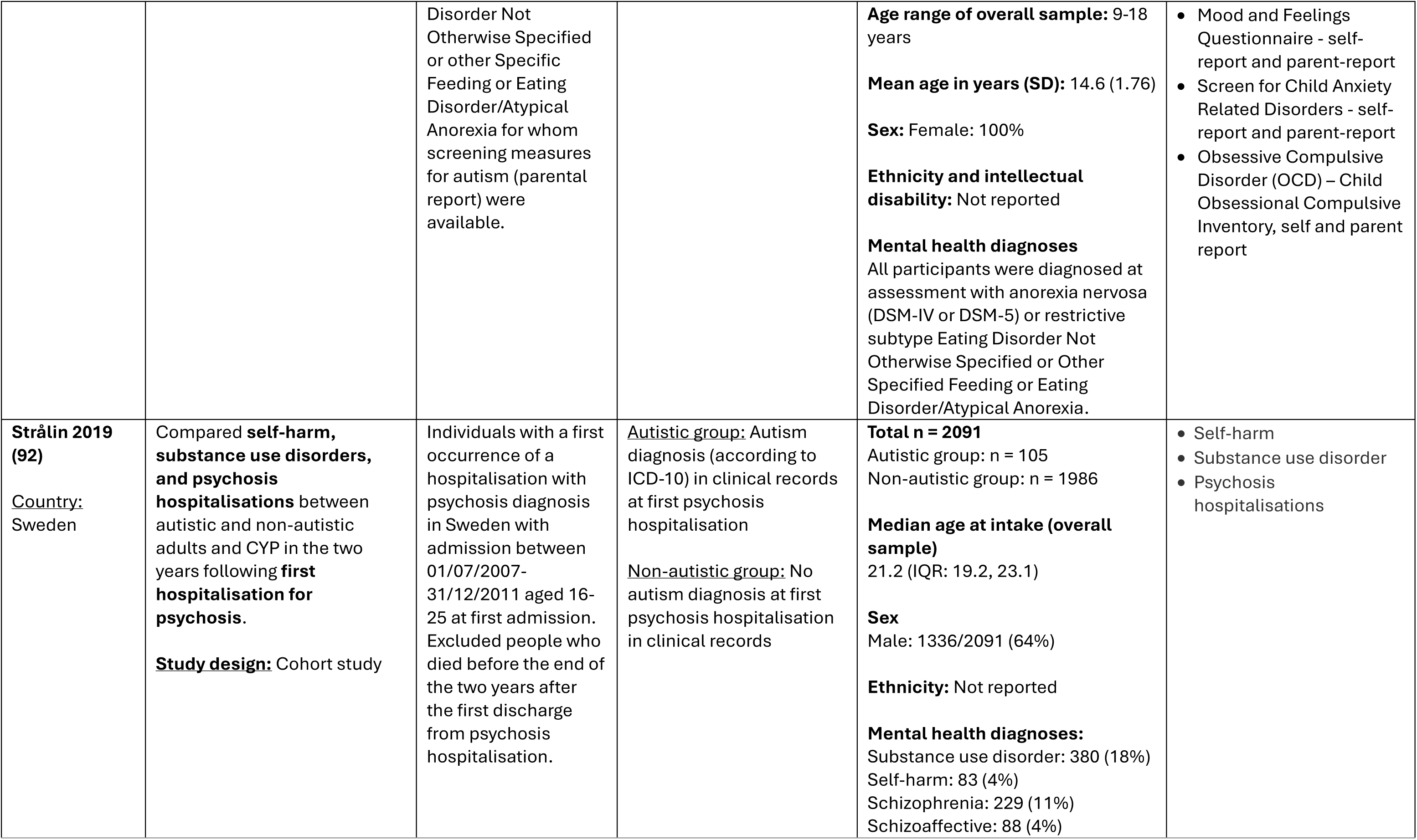
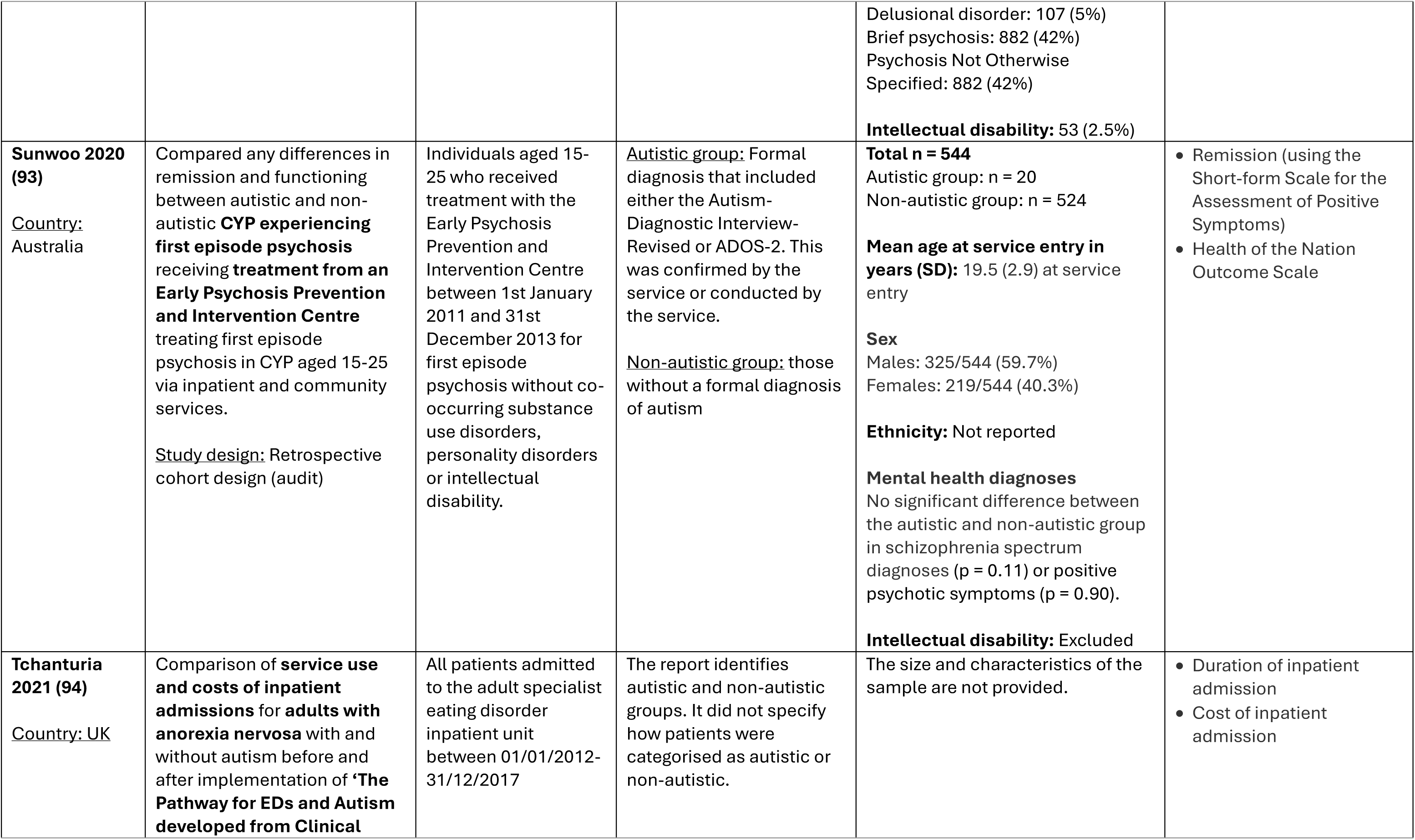
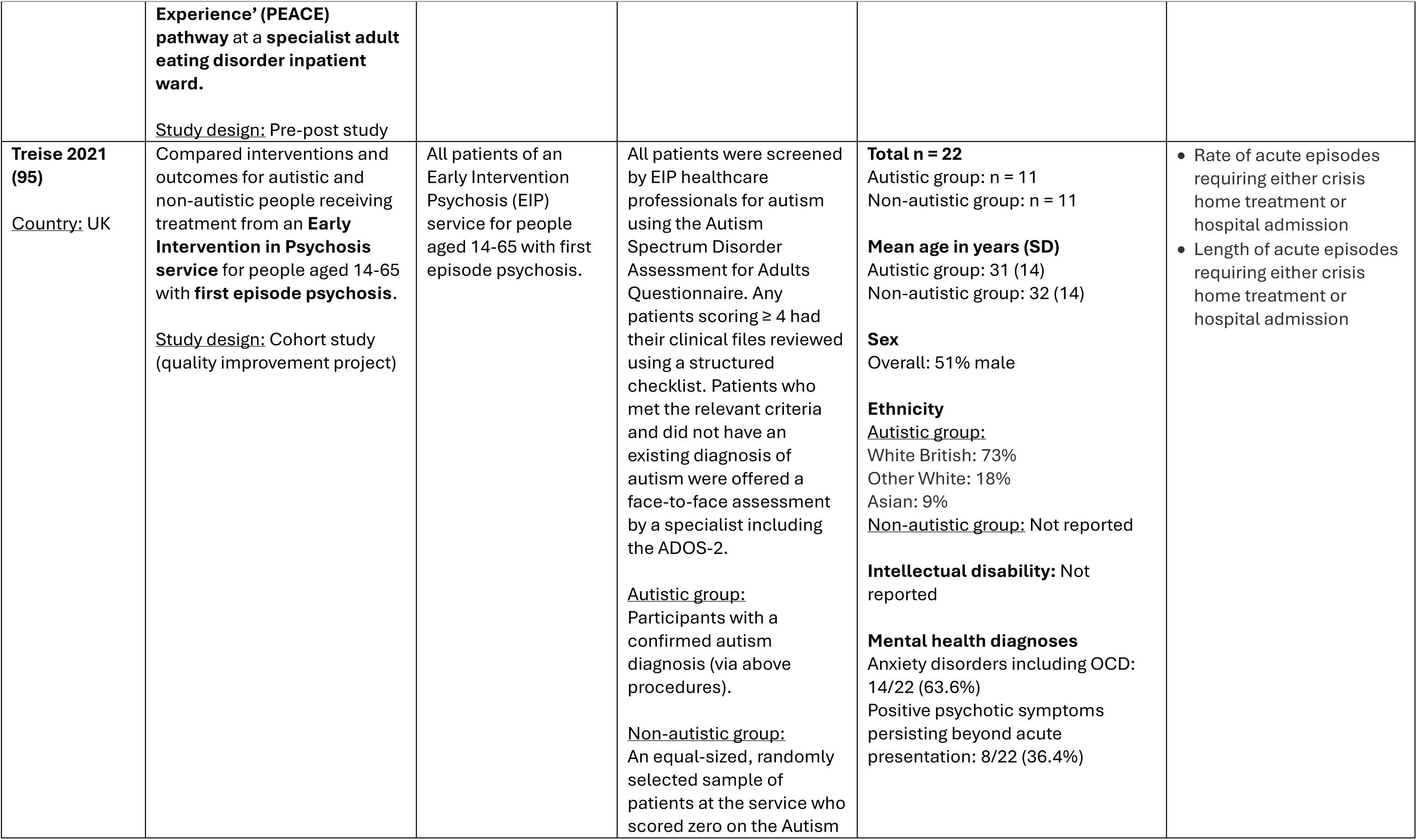
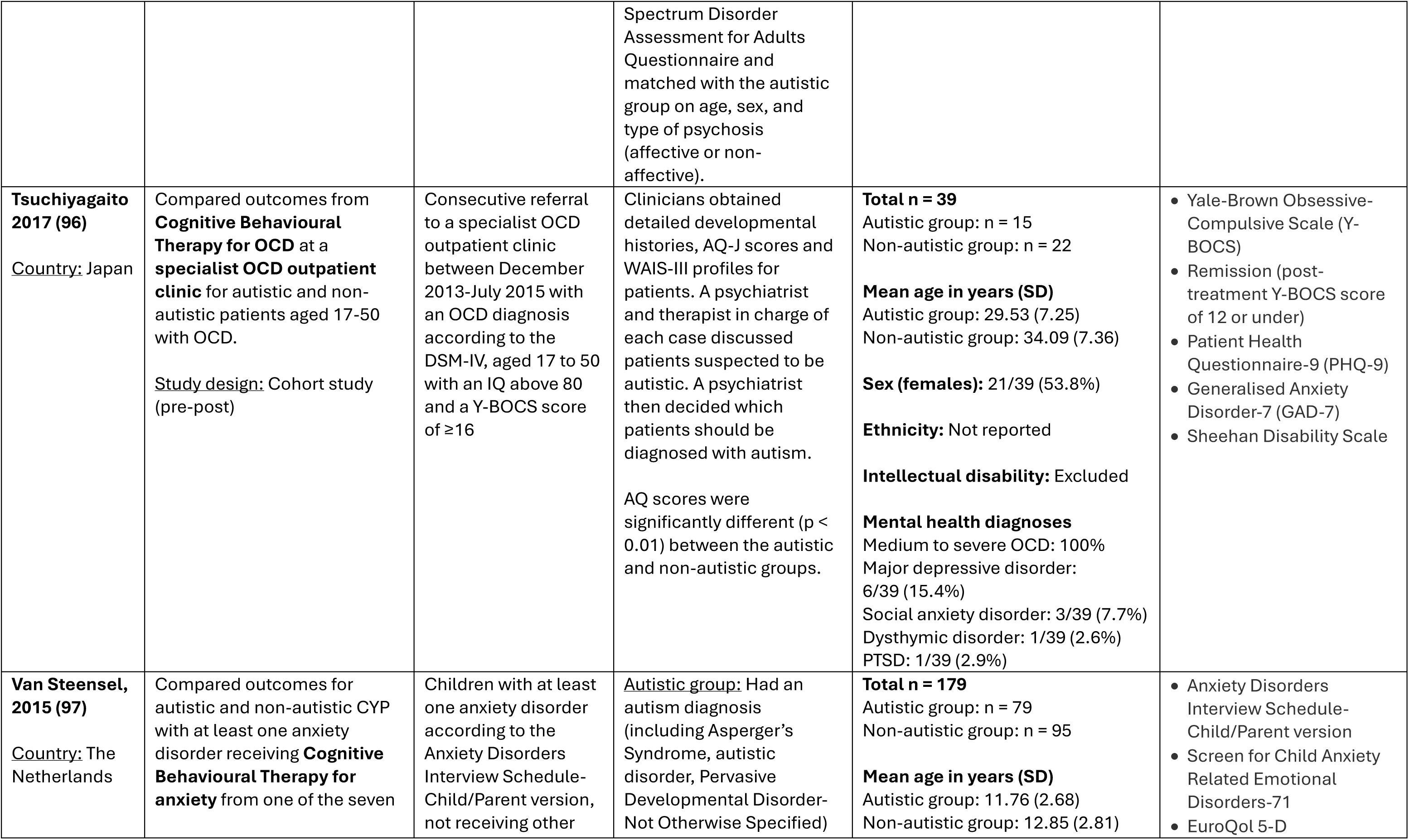
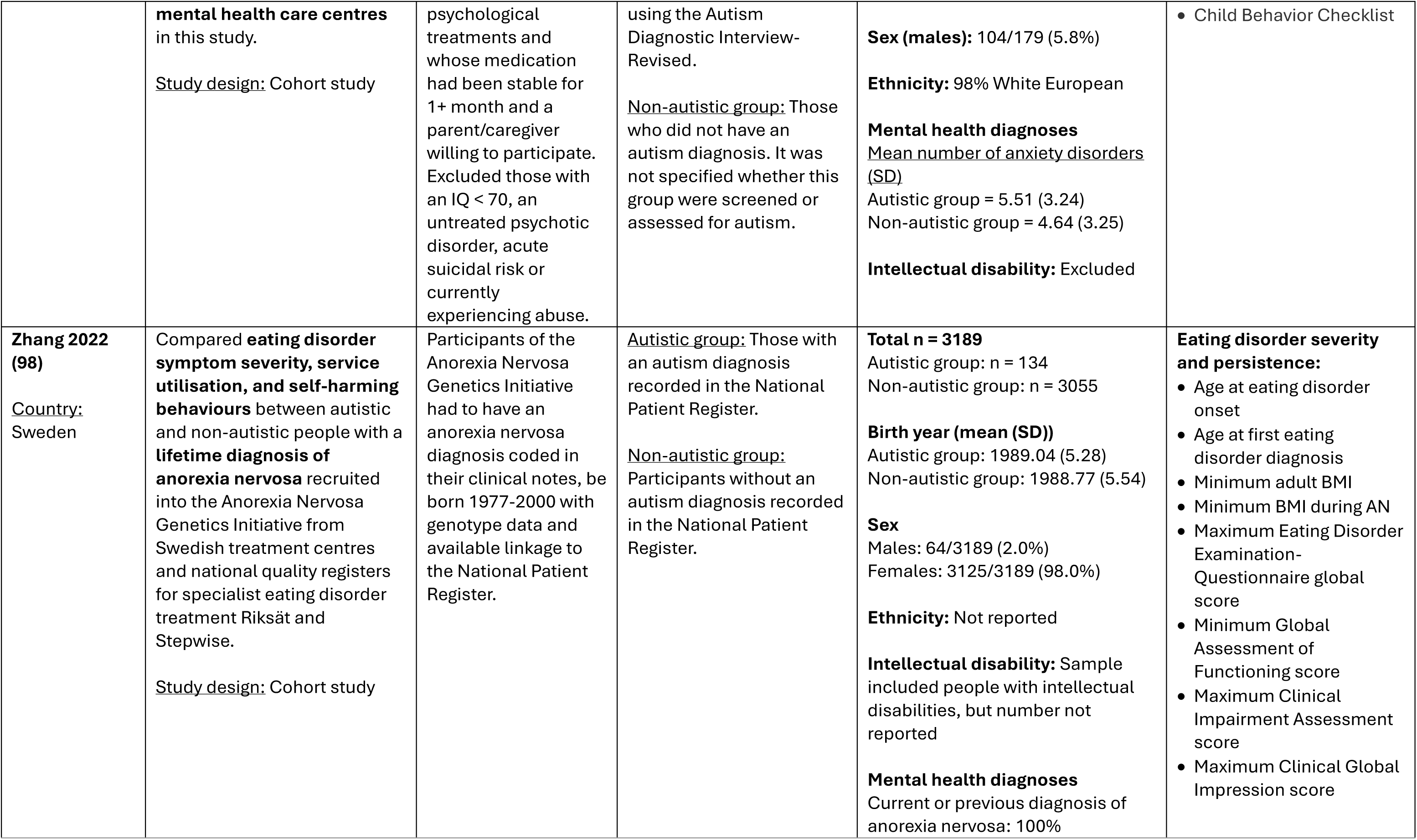
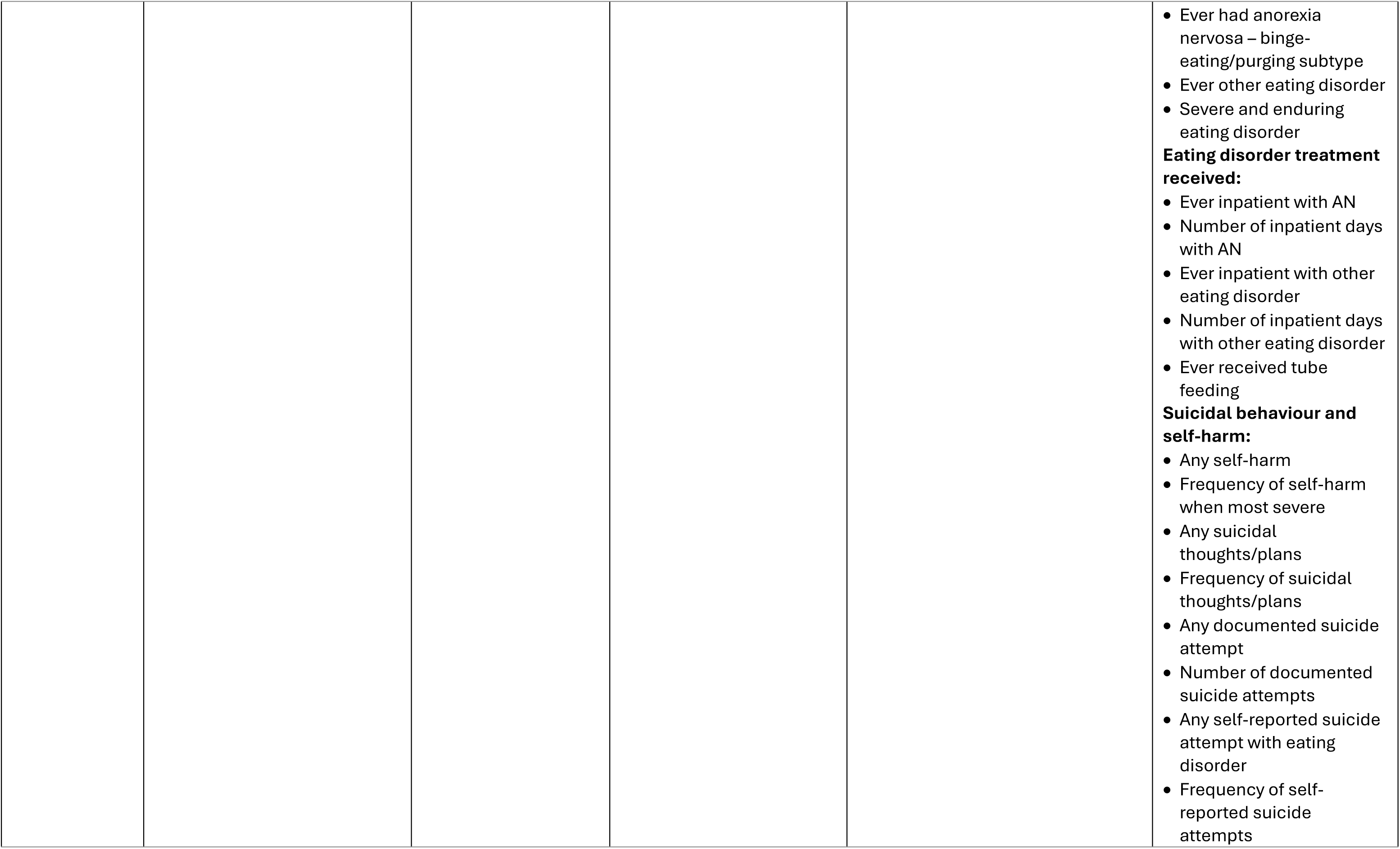
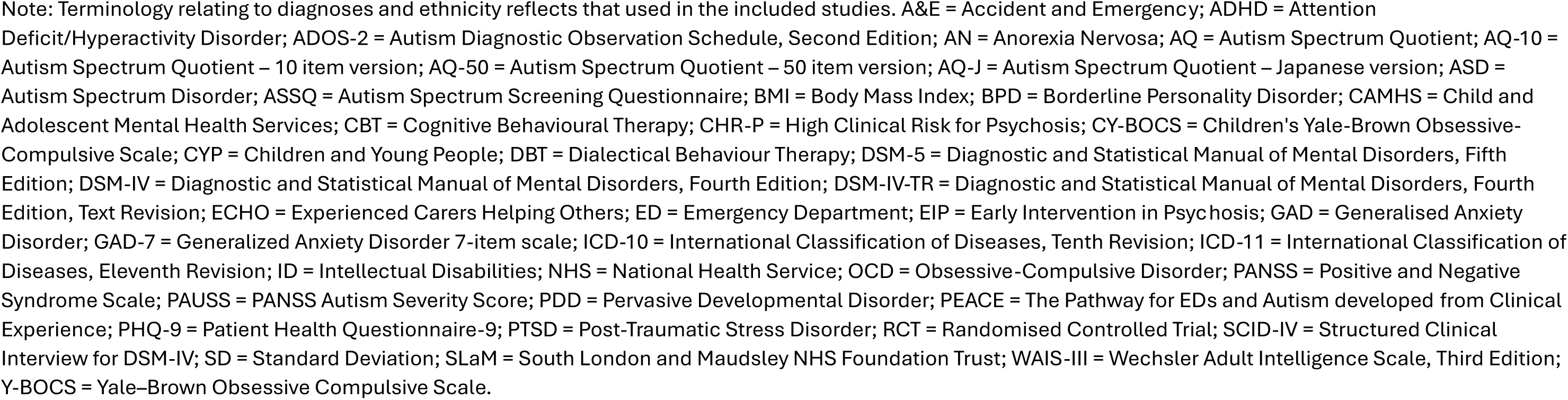
Table of study characteristics.

**Table 2:**
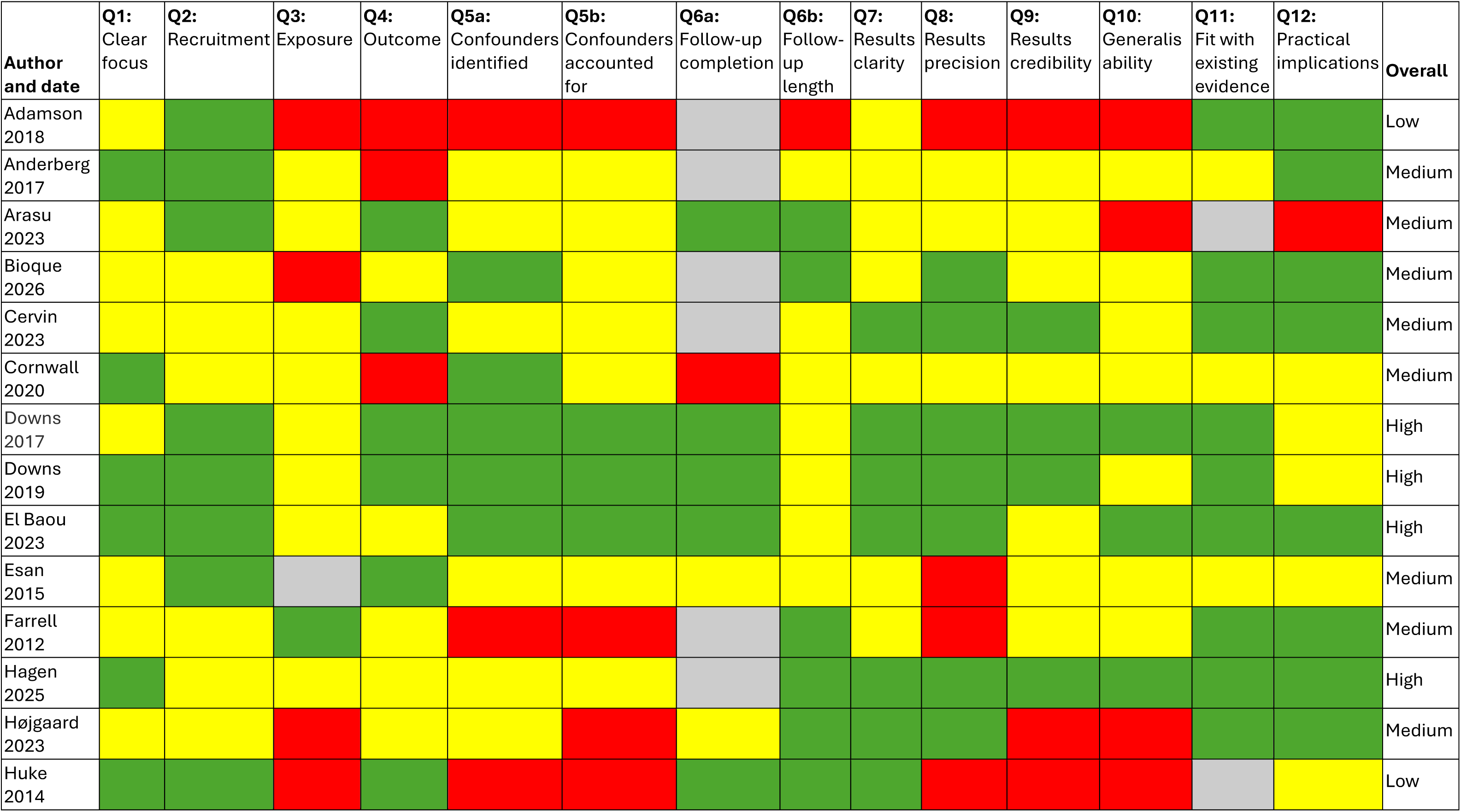
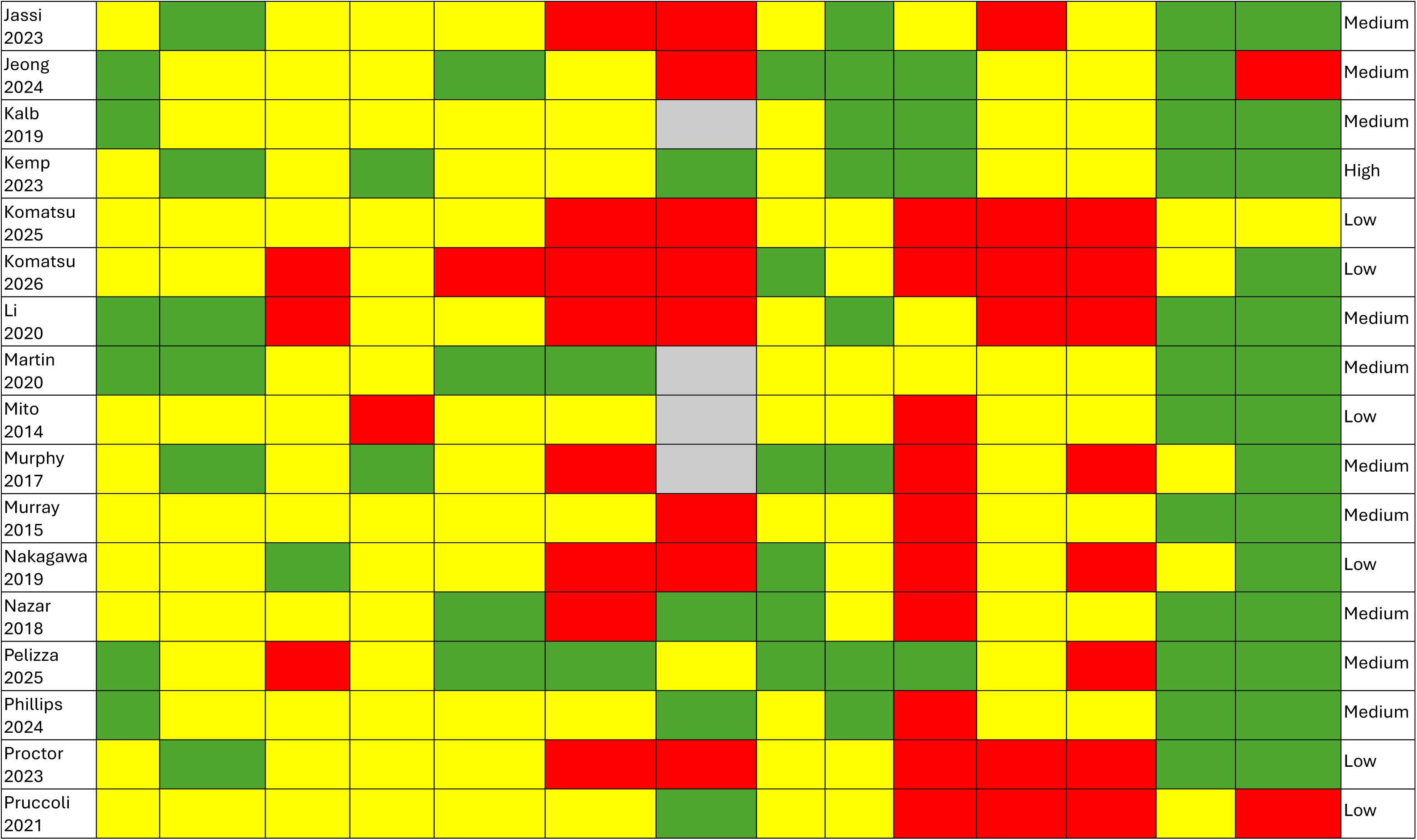
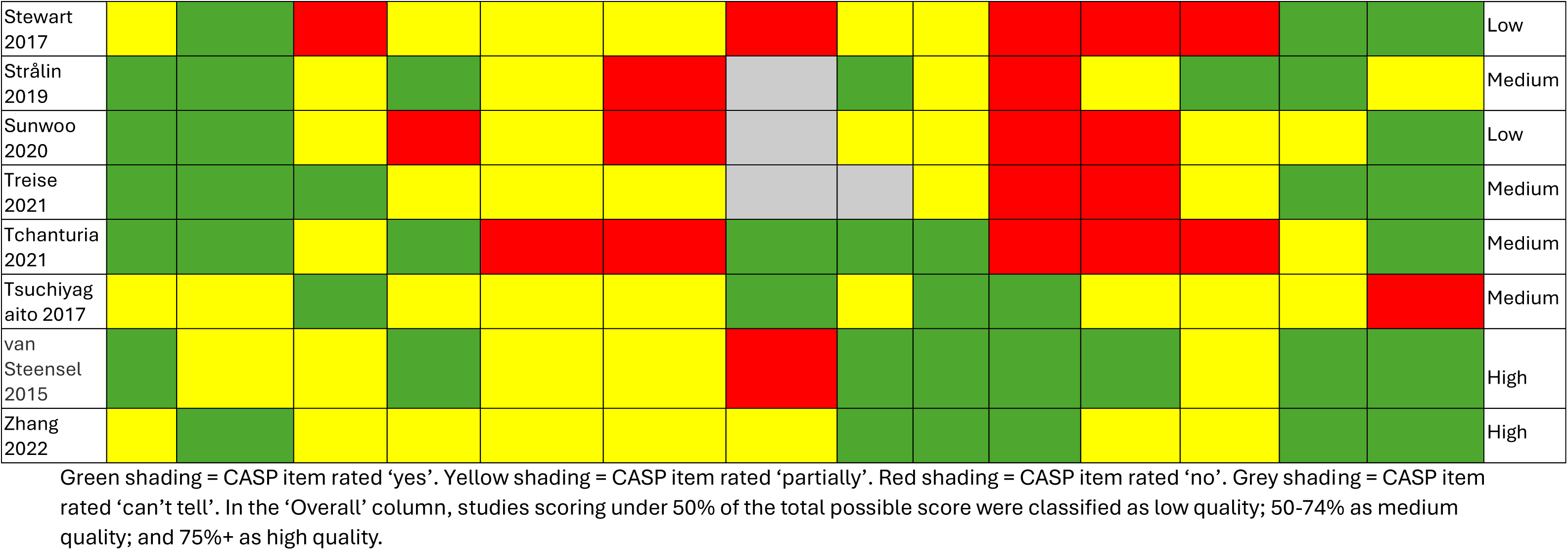
Quality appraisal ratings of included studies using the Critical Appraisal Skills Programme (CASP) Checklist for Cohort Studi. es

### 3.2. Critical Appraisal Skills Programme (CASP) checklist ratings

Ten studies (25.6%) were rated low quality, twenty-two (56.4%) medium quality, and seven (17.9%) high quality according to CASP ratings. CASP ratings for each study are displayed in Table 2. Additional File 3 presents each CASP item, the scoring criteria used, the ratings assigned to each study, and the overall quality score for each included study.

Across the included studies, the highest-scoring CASP domains related to acceptable recruitment methods, coherence with existing evidence, and discussion of implications for practice. However, several recurring limitations were identified. Only 5/39 studies adequately accounted for key confounders in their design or analyses (other factors that may influence outcomes), and only 14/39 consistently reported measures of precision (e.g. confidence intervals or standard errors) for results relevant to our review, where appropriate. Robust classification of participants as autistic or non-autistic was also uncommon (4/39 studies scored ‘yes’ on CASP item 3). Many studies relied on autism screening tools rather than more clinically-robust comprehensive diagnostic assessments recommended by clinical guidelines (99,100) and national guidance (101), did not test for statistically significant between-group differences in autism screening measure scores, or relied on historic autism diagnoses without screening participants who did not already have a diagnosis, increasing the risk of false negatives (102). Most cohort studies scored ‘no’ (11/39) or ‘can’t tell’ (13/39) on completeness of follow-up data, reflecting substantial drop-out or missing data.

CASP item 9 (“Do you believe the results?”), which captures overall methodological rigour, was scored ‘yes’ in only 5/39 studies, and ‘partially’ in 21/39 studies. ‘Partially’ or ‘no’ ratings reflected concerns about internal validity, commonly relating to classification of the autistic and non-autistic groups, handling of confounders, completeness of follow-up data, or validity of outcome measures. The extent to which findings could be generalised to wider autistic populations and settings was similarly limited (4/39 studies scoring ‘yes’, 21/39 scoring ‘partially’ on CASP item 10), often due to small sample sizes, single-site recruitment, limited reporting of sample characteristics (e.g., ethnicity), exclusion of key groups such as people with intellectual disabilities, and lack of detail about interventions provided.

### 3.3. Autism-Inclusive Research Assessment (AIRA) ratings

AIRA ratings for each study are presented in Table 3. Domain scores and rationales for ratings are provided in Additional File 3. Autistic people were not involved in research design in any of the studies. No studies involved autistic people in prioritising outcomes or deciding how they were measured. Only a minority used patient-reported outcome measures adapted or validated for autistic participants (5/39 ‘yes’, 10/39 ‘partially’), while 11/39 (28.2%) relied solely on service-use outcomes and were scored ‘N/A’. Over half of studies (29/39; 74.4%) used outcome measures that risked misinterpreting autistic traits and mental health symptoms (e.g. rigid and repetitive behaviours in autism being misinterpreted as compulsions in OCD), often relying on brief, non-adapted self-report questionnaires with limited consideration of context or function of behaviours.

**Table 3:**
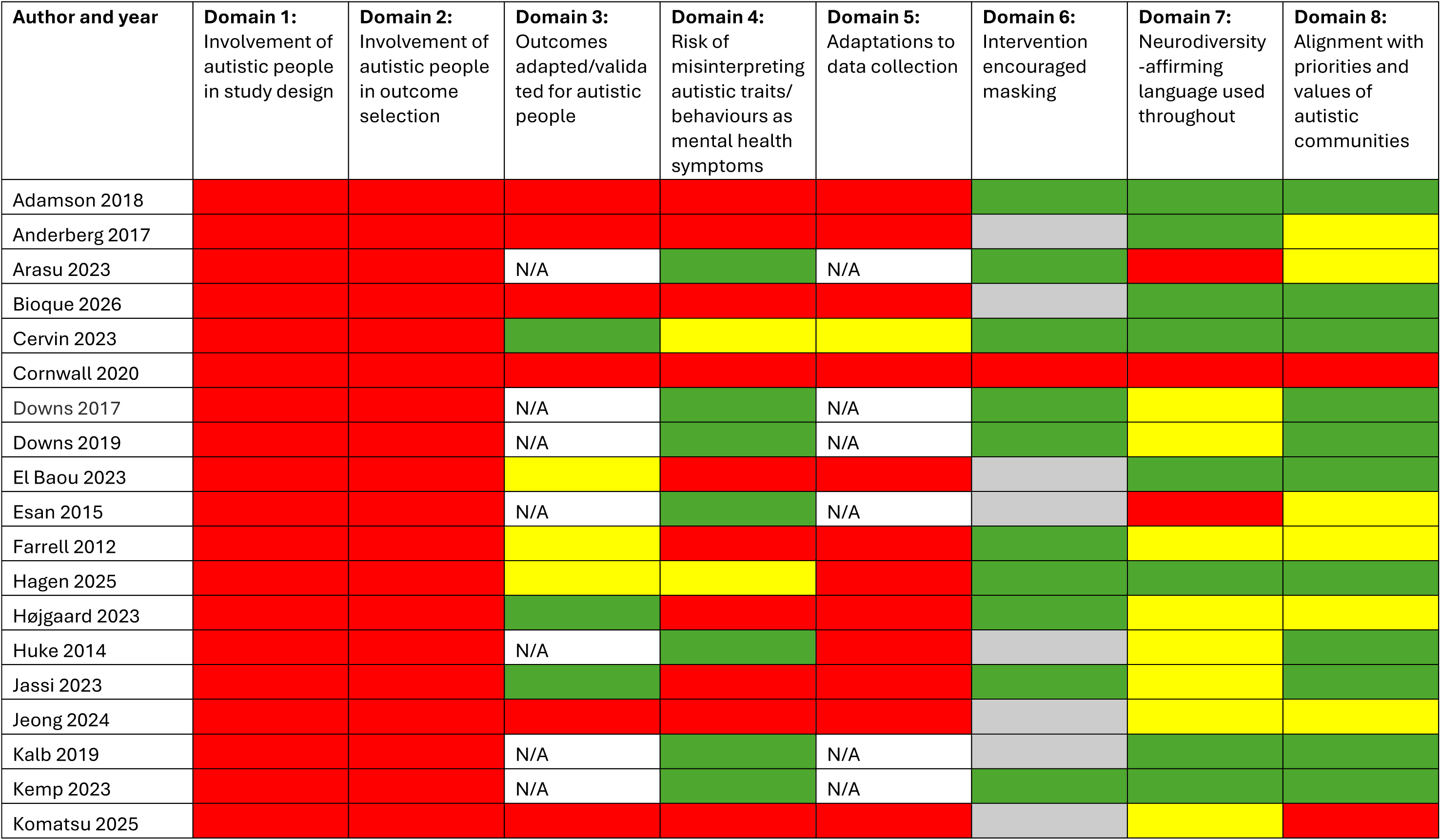
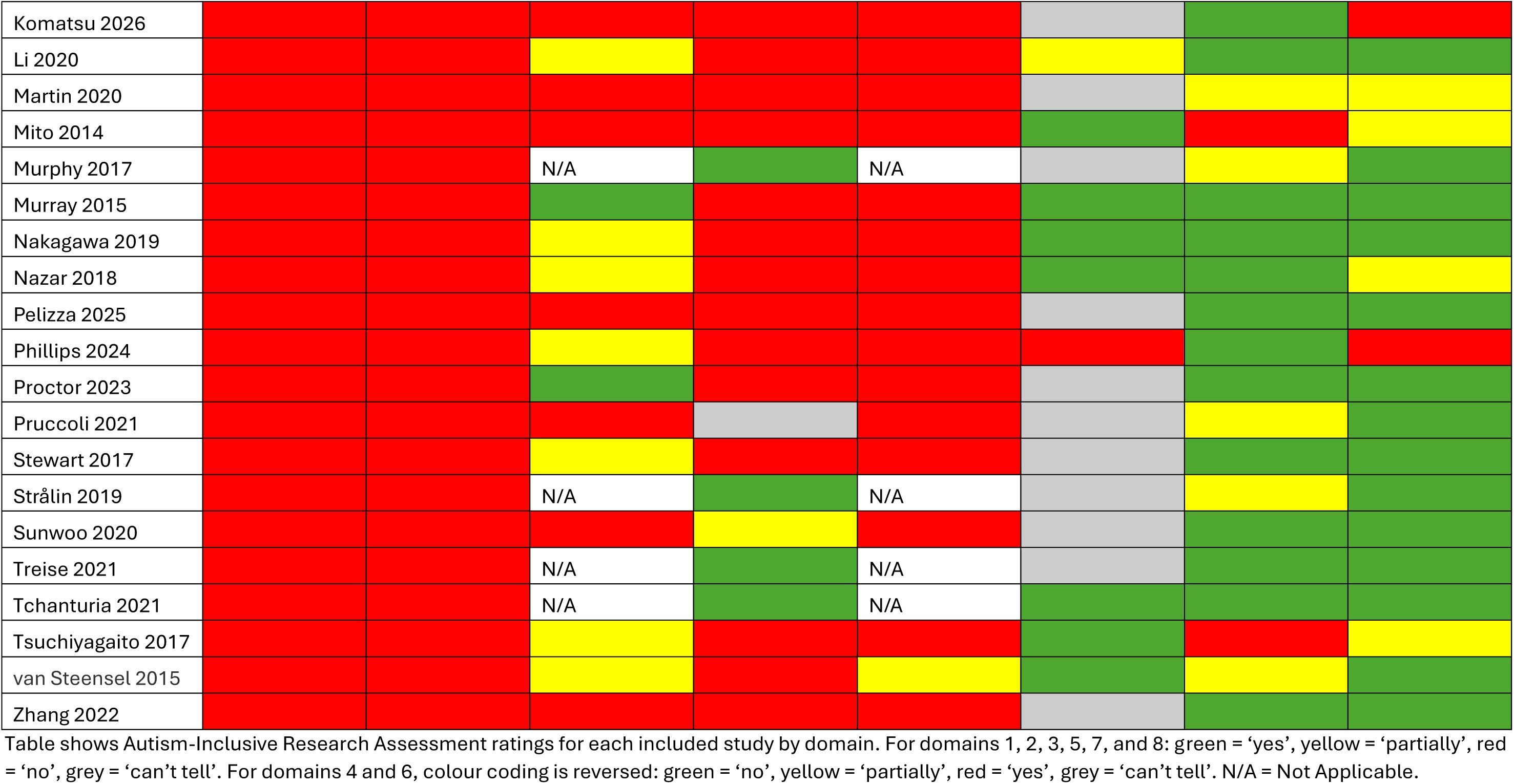
Autism-Inclusive Research Assessment ratings for each included study.

| Author and year | Domain 1:<br>Involvement of autistic people in study design | Domain 2:<br>Involvement of autistic people in outcome selection | Domain 3:<br>Outcomes adapted/validated for autistic people | Domain 4:<br>Risk of misinterpreting autistic traits/behaviours as mental health symptoms | Domain 5:<br>Adaptations to data collection | Domain 6:<br>Intervention encouraged masking | Domain 7:<br>Neurodiversity-affirming language used throughout | Domain 8:<br>Alignment with priorities and values of autistic communities |
| --- | --- | --- | --- | --- | --- | --- | --- | --- |
| Adamson 2018 |  |  |  |  |  |  |  |  |
| Anderberg 2017 |  |  |  |  |  |  |  |  |
| Arasu 2023 |  |  | N/A |  | N/A |  |  |  |
| Bioque 2026 |  |  |  |  |  |  |  |  |
| Cervin 2023 |  |  |  |  |  |  |  |  |
| Cornwall 2020 |  |  |  |  |  |  |  |  |
| Downs 2017 |  |  | N/A |  | N/A |  |  |  |
| Downs 2019 |  |  | N/A |  | N/A |  |  |  |
| El Baou 2023 |  |  |  |  |  |  |  |  |
| Esan 2015 |  |  | N/A |  | N/A |  |  |  |
| Farrell 2012 |  |  |  |  |  |  |  |  |
| Hagen 2025 |  |  |  |  |  |  |  |  |
| Højgaard 2023 |  |  |  |  |  |  |  |  |
| Huke 2014 |  |  | N/A |  |  |  |  |  |
| Jassi 2023 |  |  |  |  |  |  |  |  |
| Jeong 2024 |  |  |  |  |  |  |  |  |
| Kalb 2019 |  |  | N/A |  | N/A |  |  |  |
| Kemp 2023 |  |  | N/A |  | N/A |  |  |  |
| Komatsu 2025 |  |  |  |  |  |  |  |  |
| Komatsu 2026 |  |  |  |  |  |  |  |  |
| Li 2020 |  |  |  |  |  |  |  |  |
| Martin 2020 |  |  |  |  |  |  |  |  |
| Mito 2014 |  |  |  |  |  |  |  |  |
| Murphy 2017 |  |  | N/A |  | N/A |  |  |  |
| Murray 2015 |  |  |  |  |  |  |  |  |
| Nakagawa 2019 |  |  |  |  |  |  |  |  |
| Nazar 2018 |  |  |  |  |  |  |  |  |
| Pelizza 2025 |  |  |  |  |  |  |  |  |
| Phillips 2024 |  |  |  |  |  |  |  |  |
| Proctor 2023 |  |  |  |  |  |  |  |  |
| Pruccoli 2021 |  |  |  |  |  |  |  |  |
| Stewart 2017 |  |  |  |  |  |  |  |  |
| Strålin 2019 |  |  | N/A |  | N/A |  |  |  |
| Sunwoo 2020 |  |  |  |  |  |  |  |  |
| Treise 2021 |  |  | N/A |  | N/A |  |  |  |
| Tchanturia 2021 |  |  | N/A |  | N/A |  |  |  |
| Tsuchiyagaito 2017 |  |  |  |  |  |  |  |  |
| van Steensel 2015 |  |  |  |  |  |  |  |  |
| Zhang 2022 |  |  |  |  |  |  |  |  |

Table shows Autism-Inclusive Research Assessment ratings for each included study by domain. For domains 1, 2, 3, 5, 7, and 8: green = ‘yes’, yellow = ‘partially’, red = ‘no’, grey = ‘can’t tell’. For domains 4 and 6, colour coding is reversed: green = ‘no’, yellow = ‘partially’, red = ‘yes’, grey = ‘can’t tell’. N/A = Not Applicable.

Only 2/39 (5.1%) studies scored ‘partially’ for adapting data collection tools or procedures -Cervin et al. 2023 used a supplement to an anxiety measure to capture autism-specific symptoms of anxiety (60), and van Steensel et al. (2015) reported making minor accessibility modifications (e.g., simplifying wording or providing additional examples when needed), but no planned autism-specific adaptations (97). A minority of studies (2/39; 5.1%) included interventions with a risk of encouraging masking of autistic traits (e.g., promoting conformity to more neurotypical communication styles) (dialectical behaviour therapy [DBT] (55); radically open dialectical behaviour therapy [RO-DBT] (66)), whilst 19/39 (48.7%) could not be rated on this domain due to insufficient intervention detail.

Just over half of studies (21/39; 53.8%) used neurodiversity-affirming language throughout, such as acknowledging autistic strengths, recognising diversity within autistic populations, or focusing on adapting supports or environments rather than changing people’s autistic traits. 13/39 (33.3%) did so partially, and 5/39 (12.8%) scored ‘no’, most commonly due to deficit-based framings of autism, use of interventions that risked encouraging masking of autistic traits/behaviours, or use of stigmatising or over-generalising language. Nearly two-thirds of studies (25/39; 64.1%) were judged to align with priorities and values of autistic communities; although all included interventions targeting mental health, some were rated lower on this domain if they risked encouraging masking of autistic traits, used deficit-based framing, or lacked a neurodiversity-affirming approach.

### 3.4. Mental health treatment outcomes for autistic compared to non-autistic people

We grouped the interventions and services reported in the included studies into two categories: psychological interventions (n = 8) or multi-component or service-level approaches (n = 7) (see Table 4 for an overview).

**Table 4:** Categorisation of psychological and multi-component or service level interventions.

| Intervention/service type | Studies (n) | Citations |
| --- | --- | --- |
| ***Psychological interventions*** |  |  |
| Cognitive Behavioural Therapy (CBT) for obsessive-compulsive disorder (OCD) | 7 (17.9%) | (53,71,74,81,83,84,96) |
| CBT for anxiety | 3 (7.7%) | (60,72,97) |
| Dialectical Behaviour Therapy (DBT) | 1 (2.6%) | (55) |
| Radically Open Dialectical Behaviour Therapy (RO-DBT) | 1 (2.6%) | (66) |
| Cognitive Remediation and Emotional Skills Training (CREST) – individual and group formats | 1 (2.6%) | (56) |
| Family Therapy for Anorexia Nervosa + CBT for low mood, anxiety, or OCD when indicated | 1 (2.6%) | (91) |
| NHS Talking Therapies | 2 (5.1%) | (69,89) |
| Counselling psychotherapy at a university counselling service | 1 (2.6%) | (57) |
| ***Multi-component or service-level approaches*** |  |  |
| Emergency departments | 2 (5.1%) | (58,76) |
| Forensic intellectual disabilities inpatient service | 1 (2.6%) | (70) |
| High-secure psychiatric hospital | 1 (2.6%) | (82) |
| Specialist eating disorder inpatient units | 4 (10.3%) | (54,73,90,94) |
| Mixed specialist eating disorder settings (e.g. inpatient, step-up, day patient, outpatient) | 4 (10.3%) | (54,77,87,98) |
| Early psychosis care | 10 (25.7%) | (59,67,68,75,78,79,88, 92,93,95) |
| Child and Adolescent Mental Health Services (CAMHS) at an NHS Trust | 1 (2.5%) | (80) |

CAMHS = Child and Adolescent Mental Health Services; CBT = Cognitive Behavioural Therapy; CREST = Cognitive Remediation and Emotional Skills Training; DBT = Dialectical Behaviour Therapy; NHS = National Health Service; OCD = Obsessive-Compulsive Disorder.

‘Psychological interventions’ included structured therapies such as CBT and DBT, and services such as NHS Talking Therapies services (formerly known as Improving Access to Psychological Therapies [IAPT]) or a university counselling service, which primarily provided psychological treatments. ‘Multi-component or service-level approaches’ includes services delivering complex or multi-component interventions that could not be clearly classified as purely psychological or pharmacological, such as emergency departments or specialist inpatient eating disorder services.

#### 3.4.1. Psychological interventions

Seventeen studies compared outcomes of psychological interventions for autistic and non-autistic people. The findings below are reported by type of intervention and tabulated in Table 5.

**Table 5:** Quantitative evidence comparing mental health treatment outcomes for autistic and non-autistic people from psychological interventions.

| Author, year & study design | Sample, setting, comparison and analysis methods | Results | Quality appraisal (CASP and AIRA ratings) |
| --- | --- | --- | --- |
| **Cognitive behavioural therapy (CBT) for obsessive-compulsive disorder (OCD)** |  |  |  |
| *Adults* |  |  |  |
| **Nakagawa 2019 (84)**<br><u>Study design:</u><br>Cohort study | **Sample and setting:** Long-term follow-up of adults with OCD who received CBT for OCD.<br>**Comparison:**<br><u>Autistic group:</u> The autism diagnosis was confirmed by two experienced psychiatrists using the DSM-IV, the parent-rated Pervasive Developmental Disorders Autism Society Japan Rating Scale, and the Autism Spectrum Quotient Japanese version.<br><u>Non-autistic group:</u> Not diagnosed with autism.<br>**Analysis methods:** ANOVAs were used to compare symptom reduction at follow-up. Neither statistical controls nor stratification for confounders were used. | **Mental health symptoms:**<br>There were no significant differences between the autistic and non-autistic groups in pre-post changes in OCD or depression symptoms:<br><u>OCD symptoms</u><br>• Y-BOCS total scores (group x time effect: $F = 0.95$, $p = 0.34$)<br>• Y-BOCS Obsessive subscale scores (group x time effect: $F = 0.86$, $p = 0.36$)<br>• Y-BOCS Compulsive subscale scores (group x time effect: $F = 0.85$, $p = 0.37$)<br><u>Depression symptoms</u><br>• Beck Depression Inventory-II (group x time effect: $F = 0.68$, $p = 0.42$) | <u>Overall CASP rating:</u> Low quality<br><u>AIRA ratings:</u><br>1. Involvement of autistic people in study design: No<br>2. Involvement of autistic people in outcome selection: No<br>3. Outcomes adapted/validated for autistic people: Partially<br>4. Risk of misinterpreting autistic traits/behaviours and mental health symptoms: Yes<br>5. Adaptations to data collection: No<br>6. Intervention encouraged masking: No<br>7. Neurodiversity-affirming language used throughout: Yes<br>8. Alignment with priorities and values of autistic communities: Partially |
| *Adults and CYP* |  |  |  |
| **Tsuchiyagaito 2017 (96)**<br><u>Study design:</u><br>Cohort study (pre-post) | **Sample and setting:** People (aged 17-50) referred to an OCD outpatient clinic with medium to severe OCD who received CBT for OCD.<br>**Comparison:**<br><u>Autistic group:</u> A psychiatrist and therapist considered suspected autism cases and decided according to DSM-IV criteria, considering developmental and life histories, AQ scores and WASI-III scores.<br><u>Non-autistic group:</u> Those without a diagnosis of autism.<br>**Analysis methods:** Multilevel linear mixed model for three time points (pre-, mid-, and post-treatment) of Y-BOCS total score and Sheehan Disability Scale scores. Gender, age, age at onset, baseline values of the outcome measures, and PHQ-9 and GAD-7 were included as covariates. | There was a significant difference in change in OCD symptoms and sense of disability over time between the autistic and non-autistic groups, with the autistic group reporting significantly higher OCD symptoms and sense of disability, and lower OCD remission rates.<br>**OCD symptoms (Y-BOCS):**<br>There was a significant interaction between time and group in the total Y-BOCS scores ($p < 0.001$) – indicating significant differences in changes in Y-BOCS scores across pre-, mid-, and post-treatment timepoint between the autistic and non-autistic groups. Mean Y-BOCS scores were significantly higher at mid-point and post-intervention in the autistic group, compared to the non-autistic group. For the non-autistic group, there was already a significant reduction in Y-BOCS scores at mid-treatment, but not for the autistic group.<br>**Sheehan Disability Scale:**<br>There was a significant interaction between time and group in the Sheehan Disability Scale scores ($p < 0.001$) – indicating significant differences in changes in Sheehan Disability Scale scores across pre-, mid-, and post-treatment timepoints between the autistic and non-autistic groups. Over the course of the CBT sessions, the autistic group reported a greater sense of disability.<br>**Remission rates (post-treatment Y-BOCS score <13)**<br>Autistic participants had significantly lower remission rates (13.33%) compared to non-autistic participants (54.54%) ($p$ value not reported). | <u>Overall CASP rating:</u> Medium quality<br><u>AIRA ratings:</u><br>1. Involvement of autistic people in study design: No<br>2. Involvement of autistic people in outcome selection: No<br>3. Outcomes adapted/validated for autistic people: Partially<br>4. Risk of misinterpreting autistic traits/behaviours and mental health symptoms: Yes<br>5. Adaptations to data collection: No<br>6. Intervention encouraged masking: No<br>7. Neurodiversity-affirming language used throughout: No<br>8. Alignment with priorities and values of autistic communities: Partially |
| CYP |  |  |  |
| **Farrell 2012 (71)**<br><u>Study design:</u><br>Prospective cohort | **Sample and setting:** CYP aged 7-17 consecutively referred for treatment of OCD to the University Psychology Clinic, who had a diagnosis of OCD, completed the assessment and participated in treatment. | There were <u>no significant differences</u> (all $p > 0.05$) between the autistic and non-autistic groups in change over time (pre-intervention to post-intervention and 6-month follow-up) in OCD symptoms, diagnostic severity, or functional impairment: | <u>Overall CASP rating:</u> Medium quality<br><u>AIRA ratings:</u> |
| study (open pilot trial) | **Comparison:**<br><u>Autistic group:</u> Positive screen of PDD on the ADIS-P and a confirmed diagnosis of PDD or an ASD (including Asperger's syndrome) by a community paediatrician or child psychiatrist.<br><u>Non-autistic group:</u> People with OCD but no positive screen of autism on the ADIS-P and no confirmed diagnosis of autism.<br>**Analysis methods:**<br>• Repeated factorial ANOVAs with a Time (pre-, post-, 6-month follow-up) x Group x group (comorbid PDD, ADHD or depression versus no comorbidity group) design.<br>• Post-hoc analyses (t tests) of significant multivariate time main effects explored changes in outcomes across the entire sample.<br>• Proportions of treatment responders and remitters between groups were compared using Chi-squared tests. | • **OCD diagnostic severity** (measured with NIMH Global OC Scale and ADIS-P Clinician Severity Ratings)<br>• **OCD symptoms severity** (measured using the Child Yale Brown Obsessive-Compulsive Scale (CY-BOCS))<br>• **OCD functional impairment** (Child OCD Impact Scale- Parent report (COIS-P)) and Child ((COIS-C))<br>There was also <u>no significant difference</u> between the autistic and non-autistic groups at post-intervention or 6-month follow-up in treatment response or remission rates:<br>• **Treatment response rates** (>25% reduction in CY-BOCS) (both $p > 0.05$)<br>• **Treatment remission rates** (score of <14 on CY-BOCS and at least 50% reduction on CY-BOCS) at post-intervention or 6-month follow-up (both $p > 0.05$). | 1. Involvement of autistic people in study design: No<br>2. Involvement of autistic people in outcome selection: No<br>3. Outcomes adapted/validated for autistic people: Partially<br>4. Risk of misinterpreting autistic traits/behaviours and mental health symptoms: Yes<br>5. Adaptations to data collection: No<br>6. Intervention encouraged masking: No<br>7. Neurodiversity-affirming language used throughout: Partially<br>8. Alignment with priorities and values of autistic communities: Partially |
| **Højgaard 2023 (53)**<br><u>Study design:</u><br>Cohort study | **Sample and setting:** CYP aged 7-17 years with subclinical autistic traits and OCD in the Nordic Long-Term OCD Treatment Study (NordLOTS) receiving CBT for OCD. CYP with autism were excluded (except those with a PDD-NOS diagnosis).<br>**Comparison:**<br><u>High autistic traits (HAT) group:</u> ASSQ scores of 17+ | There were mostly no significant differences in OCD symptoms between the HAT and LAT groups at post-intervention or follow-up timepoints, except for when the groups were defined using a cut-off on the ASSQ motor/OCD/tics subscale.<br>**OCD symptom severity (CY-BOCS)**<br>• <u>ASSQ total score and social difficulties scale:</u> There were no significant differences at post-treatment or any follow-up time point (all $p > 0.05$) between the autistic and non-autistic groups (defined by 90<sup>th</sup> percentile cut-offs on the ASSQ total | <u>Overall CASP rating:</u> Medium quality<br><u>AIRA ratings:</u><br>1. Involvement of autistic people in study design: No<br>2. Involvement of autistic people in outcome selection: No |
|  | <u>Low autistic traits (LAT) group:</u> ASSQ scores <17<br>**Analysis methods:** Used a series of piecewise regression and linear mixed effects models to compare changes in CY-BOCS symptoms at baseline, post-intervention, and 1-year, 2-year and 3-year follow-up between autistic and non-autistic groups (categorised based on 90<sup>th</sup> percentile cut-offs on the ASSQ including:<br>- ASSQ total score: >16<br>- ASSQ autistic style subscale: >4<br>- ASSQ social difficulties subscale: >7<br>- ASSQ motor/tics/OCD subscale: >6) | score or social difficulties subscale) in OCD symptom severity.<br>• <u>ASSQ motor/OCD/tics subscale:</u> The ≥90<sup>th</sup> percentile group had a significantly higher OCD severity score at post-treatment ($t = 2.75$, $p = 0.006$), though it was significantly lower at 3-year follow-up ($t = -2.06$, $p = 0.040$). There were no significant differences at any other time point (all $p > 0.05$).<br>• <u>ASSQ autistic style subscale:</u> There were no significant differences in estimated CY-BOCS scores between the groups at any time point (all $p > 0.05$). | 3. Outcomes adapted/validated for autistic people: Yes<br>4. Risk of misinterpreting autistic traits/behaviours and mental health symptoms: Yes<br>5. Adaptations to data collection: No<br>6. Intervention encouraged masking: No<br>7. Neurodiversity-affirming language used throughout: Partially<br>8. Alignment with priorities and values of autistic communities: Partially |
| **Jassi 2023 (74)**<br><u>Study design:</u><br>Cohort study (pre-post) | **Sample and setting:** CYP (aged 6-18) with OCD referred to a national and specialist outpatient OCD, Body Dysmorphic Disorder and Related Disorders Clinic.<br>**Comparison:**<br><u>Autistic group:</u> Confirmed autism diagnosis from a consultant psychiatrist in a CAMHS service.<br><u>Non-autistic group:</u> Those with no autism diagnosis (this group was not screened or assessed for autism).<br>**Analysis methods:**<br>• Repeated-measures ANOVA compared treatment response (on CY-BOCS) between autistic and non-autistic participants with baseline and post- | The autistic group showed significantly smaller improvements in OCD symptoms, and significantly lower rates of treatment response and remission, compared to the non-autistic group:<br>**Change in OCD symptoms (CY-BOCS scores):**<br>There was a significant group by time interaction ($F(1,642) = 8.72$, $p = 0.003$). The <u>autistic group showed significantly less improvement than the non-autistic group</u> in CY-BOCS scores at post-treatment ($F(1,642) = 22.34$, $p < 0.001$).<br>**Treatment responders (35%+ reduction on CY-BOCS):** A significantly lower proportion of participants in the autistic group responded to treatment (53%) compared to the non-autistic group (76.7%) ($OR = 0.34$, 95% CI: 0.21 to 0.57, $p < 0.001$).<br>**Remission (CY-BOCS score <13 at end of treatment):** A significantly lower proportion of participants in the autistic | <u>Overall CASP rating:</u> Medium quality<br><u>AIRA ratings:</u><br>1. Involvement of autistic people in study design: No<br>2. Involvement of autistic people in outcome selection: No<br>3. Outcomes adapted/validated for autistic people: Yes<br>4. Risk of misinterpreting autistic traits/behaviours and mental health symptoms: Yes<br>5. Adaptations to data collection: No |
|  | treatment outcome data available (n = 323, 52%).<br>Differences in rates of treatment responders (CY-BOCS decrease of 35%+) and remitters (post-treatment CY-BOCS score of <13) examined using logistic regression. | group remitted (31%) compared to the non-autistic group (49.8%) (OR = 0.45, 95% CI: 0.28 to 0.75, p = 0.002). | 6. Intervention encouraged masking: No<br>7. Neurodiversity-affirming language used throughout: Partially<br>8. Alignment with priorities and values of autistic communities: Yes |
| **Murray 2015 (83)**<br><u>Study design:</u><br>Cohort study (with matched controls) | **Sample and setting:** Consecutive CYP who received CBT for OCD at a national specialist OCD clinic (referred 2007-2011) diagnosed with OCD and with complete data. Matched to non-autistic cases who had an OCD diagnosis and the same complete data.<br>**Comparison:**<br><u>Autistic group:</u> Had an autism diagnosis made by a consultant psychiatrist in community CAMHS.<br><u>Non-autistic:</u> No recorded autism diagnosis.<br>**Analysis methods:**<br>Independent t-tests compared pre-post mean percentage decrease in CY-BOCS scores between autistic and non-autistic groups.<br>Response and remission rates were compared between the groups using chi-squared tests. | The autistic group had significantly smaller improvements in OCD symptoms, and significantly lower remission rates post-CBT, compared to the non-autistic group. However, there were no significant between-group differences in response rates post-CBT:<br>**Change in OCD symptoms over the course of CBT:**<br>The percentage decrease in OCD symptoms (CY-BOCS scores) was significantly lower in the autistic group compared to the non-autistic group (t(42) = -2.50, p = 0.02, d = -0.75).<br>**Response rates following CBT (decrease in CY-BOCS of 35%+):** There was no significant difference in rates of treatment response between the autistic (46%) and non-autistic (83%) groups ($\chi^2(1, 44) = 3.36$, p = 0.07).<br>**Remission rates (post-treatment CY-BOCS of <13):**<br>Participants in the autistic group (9%) were significantly less likely to be in remission by the end of treatment compared to the non-autistic group (46%) ($\chi^2(1, 44) = 7.33$, p = 0.01). | <u>Overall CASP rating:</u> Medium quality<br><u>AIRA ratings:</u><br>Involvement of autistic people in study design: No<br>Involvement of autistic people in outcome selection: No<br>Outcomes adapted/validated for autistic people: Yes<br>Risk of misinterpreting autistic traits/behaviours and mental health symptoms: Yes<br>Adaptations to data collection: No<br>Intervention encouraged masking: No<br>Neurodiversity-affirming language used throughout: Yes<br>Alignment with priorities and values of autistic communities: Yes |
| **Cognitive behavioural therapy (CBT) for anxiety** |  |  |  |
| CYP |  |  |  |
| **Cervin 2023 (60)**<br><u>Study design:</u><br>Cohort study (secondary analysis of data pooled from 6 RCTs) | **Sample, setting and comparison groups:** CYP with at least one anxiety disorder randomised to CBT in a CAMHS outpatient setting or a control condition across five RCTs. And non-autistic youth with a primary diagnosis of an anxiety disorder randomised to CBT in another RCT.<br>**Analysis methods:** Data from the four RCTs were pooled. Ordinal logistic regression compared outcomes between autistic and non-autistic CYP receiving CBT. Age, sex, pre-CBT Paediatric Anxiety Rating Scale (PARS) domain severity, and trial were included as covariates. An alpha level of 0.01 was used as an indicator of statistical significance to account for familywise error across multiple models. | The autistic group reported significantly more physical symptoms of anxiety and impact of anxiety on family life post-CBT than the non-autistic group. However, there were no significant between-group differences in anxiety symptom frequency, intensity, avoidance, or impact on school/social life post-CBT.<br>**Pediatric Anxiety Rating Scale (PARS) scores post-CBT:**<br>• Autistic CYP had significantly more physical symptoms of anxiety (proportional OR = 2.44, 99%CI = 1.34–4.50, $p < 0.001$) and family interference (proportional OR = 2.22, 99%CI = 1.23–4.04, $p = 0.001$) post-CBT than non-autistic CYP.<br>• There were no significant differences between groups post-CBT for the other PARS domains (symptom frequency, intensity of anxiety, avoidance, or school/social interference) (all $p > 0.01$).<br>• Compared to non-autistic participants, autistic participants had a significantly higher risk of any family interference versus no interference (OR = 5.30, $p < 0.001$), and mild to severe interference versus minimal/no interference (OR = 3.22, $p < 0.001$) post-CBT.<br>• Across PARS domains, all significant effects were small to moderate. | <u>Overall CASP rating:</u> Medium quality<br><u>AIRA ratings</u><br>1. Involvement of autistic people in study design: No<br>2. Involvement of autistic people in outcome selection: No<br>3. Outcomes adapted/validated for autistic people: Yes<br>4. Risk of misinterpreting autistic traits/behaviours and mental health symptoms: Partially<br>5. Adaptations to data collection: Partially<br>6. Intervention encouraged masking: No<br>7. Neurodiversity-affirming language used throughout: Yes<br>8. Alignment with priorities and values of autistic communities: Yes |
| **Hagen 2025 (72)**<br><u>Study design:</u><br>Cohort study<br>(Same sample as in van Steensel et al., 2015) **(97)** | **Sample and setting:** CYP (aged 7-18) with at least one anxiety disorder receiving CBT for anxiety from one of the seven mental health centres in this study.<br>**Comparison:**<br><u>Autistic group:</u> Autism diagnosis (including Asperger's Syndrome, autistic disorder, PDD- | **Changes in family functioning**<br>In multilevel analyses, there were no significant differences between autistic and non-autistic participants in changes in family functioning from pre- to post-intervention or from post-intervention to the 3-month, 1-year, and 2-year follow-ups, when considering either Family Functioning Scale (FFS) 'relationship' or 'system maintenance' dimensions. Full statistics reported below: | <u>Overall CASP rating:</u> High quality<br><u>AIRA ratings:</u><br>1. Involvement of autistic people in study design: No |
|  | NOS) using Autism Diagnostic Interview-Revised.<br><u>Non-autistic group:</u> No autism diagnosis. It was not specified if this group were screened or assessed for autism.<br>**Analysis methods:**<br>• Multilevel analyses examined changes in family functioning (FFS scores) following treatment. All models used a first-order autoregression covariate structure. Scores on the family relational functioning dimension and system maintenance dimension were dependent variables. Time (pre-treatment, 3-month, 1-year, and 2-year follow-up, each contrasted against post-treatment), diagnosis (presence or absence of autism), and their interaction (time*diagnosis) were entered as predictors, with age and gender included as covariates. Separate analyses were conducted for child, mother, and father reports.<br>• Multilevel analyses explored whether family functioning at pre-treatment predicted a reduction in anxiety symptoms post-treatment. Time (pre-treatment, 3-month, 1-year, and 2-year follow-ups), diagnosis (presence or absence of autism), Family Functioning Scale score at pre-treatment were entered as predictors. Age and gender were included as covariates. Separate analyses were conducted for child, mother, and father reports. | **Changes in Family Functioning Scale (FFS) relationship dimension following treatment** (reference category = post-test)<br><u>Autism * pre-intervention FFS relationship dimension scores interaction:</u><br>Child-reported FFS: estimate = 0.04, SE = 0.123, p = 0.758<br>Mother-reported FFS: estimate = 0.11, SE = 0.112, p = 0.320<br>Father-reported FFS: estimate = 0.07, SE = 0.140, p = 0.614<br><u>Autism * 3-month follow-up FFS relationship dimension scores interaction:</u><br>Child-reported FFS: estimate = 0.08, SE = 0.130, p = 0.525<br>Mother-reported FFS: estimate = 0.02, SE = 0.117, p = 0.879<br>Father-reported FFS: estimate = 0.02, SE = 0.154, p = 0.899<br><u>Autism * 1-year follow-up FFS relationship dimension scores interaction:</u><br>Child-reported FFS: estimate = 0.17, SE = 0.174, p = 0.328<br>Mother-reported FFS: estimate = -0.10, SE = 0.163, p = 0.522<br>Father-reported FFS: estimate = -0.26, SE = 0.213, p = 0.231<br><u>Autism * 2-year follow-up FFS relationship dimension scores interaction:</u><br>Child-reported FFS: estimate = -0.02, SE = 0.197, p = 0.909<br>Mother-reported FFS: estimate = -0.06, SE = 0.185, p = 0.752<br>Father-reported FFS: estimate = 0.12, SE = 0.245, p = 0.614<br>**Changes in Family Functioning Scale system maintenance dimension following treatment** (reference category = post-test)<br><u>Autism * pre-intervention FFS system maintenance dimension scores interaction:</u> | 2. Involvement of autistic people in outcome selection: No<br>3. Outcomes adapted/validated for autistic people: Partially<br>4. Risk of misinterpreting autistic traits/behaviours and mental health symptoms: Partially<br>5. Adaptations to data collection: No<br>6. Intervention encouraged masking: No<br>7. Neurodiversity-affirming language used throughout: Yes<br>8. Alignment with priorities and values of autistic communities: Yes |
|  | • Bonferroni correction for six models was applied so $p < 0.0083$ was considered significant. | Child-reported FFS: estimate = 0.09, SE = 0.148, $p = 0.525$<br>Mother-reported FFS: estimate = 0.15, SE = 0.126, $p = 0.226$<br>Father-reported FFS: estimate = 0.08, SE = 0.160, $p = 0.600$<br><u>Autism * 3-month follow-up FFS system maintenance dimension scores interaction:</u><br>Child-reported: estimate = 0.06, SE = 0.157, $p = 0.724$<br>Mother-reported: estimate = 0.05, SE = 0.132, $p = 0.720$<br>Father-reported: estimate = 0.02, SE = 0.176, $p = 0.898$<br><u>Autism * 1-year follow-up FFS system maintenance dimension scores interaction:</u><br>Child-reported: estimate = 0.08, SE = 0.205, $p = 0.687$<br>Mother-reported: estimate = 0.04, SE = 0.178, $p = 0.839$<br>Father-reported: estimate = 0.39, SE = 0.238, $p = 0.105$<br><u>Autism * 2-year follow-up FFS system maintenance dimension scores interaction:</u><br>Child-reported: estimate = 0.26, SE = 0.225, $p = 0.256$<br>Mother-reported: estimate = 0.08, SE = 0.204, $p = 0.708$<br>Father-reported: estimate = -0.06, SE = 0.268, $p = 0.831$ |  |
| **Van Steensel 2015 (97)**<br><u>Study design:</u><br>Cohort study<br>(Same sample as in Hagen et al., 2025) **(72)** | **Sample and setting:** CYP with at least one anxiety disorder receiving CBT for anxiety from one of the seven mental health centres in this study.<br>**Comparison:**<br><u>Autistic group:</u> Autism diagnosis (including Asperger's Syndrome, autistic disorder, PDD-NOS) using the Autism-Diagnostic Interview-Revised.<br><u>Non-autistic group:</u> Those who did not have an autism diagnosis. It was not specified if this group were screened or assessed for autism. | The autistic group was significantly less likely to be free of <u>all</u> anxiety disorders post-intervention and at follow-up timepoints. However, there were no significant differences between groups in proportion free from the <u>primary</u> anxiety disorder or anxiety disorder severity at any timepoint. There was also no significant between-group difference in change in anxiety disorder severity. Findings for anxiety symptoms were mixed and varied depending on time point and whether they were reported by the child or parent.<br>The autistic group showed smaller improvements in health-related quality of life at all time points. There were mostly no significant between-group differences in changes in Child Behavior Checklist scores over time, but Child Behavior | <u>Overall CASP rating:</u> High quality<br><u>AIRA ratings:</u><br>1. Involvement of autistic people in study design: No<br>2. Involvement of autistic people in outcome selection: No<br>3. Outcomes adapted/validated for autistic people: Partially<br>4. Risk of misinterpreting autistic traits/behaviours |
|  | **Analysis methods:**<br>• Logistic regression analyses explored differences in treatment outcomes between autistic and non-autistic groups for dichotomous variables. The ‘last observation carried forward’ method was used for cases that did not complete the assessment.<br>• Multilevel analyses using maximum likelihood estimation procedures were used for continuous outcome measures.<br>• Repeated measures (pre-, post-, and 3-year follow-up assessments) were nested within respondents, and respondents (children, mothers, and fathers) were nested within families.<br>• If an interaction effect was significant, then additional multilevel analyses examined time/respondent/group effects for different groups/respondents. | Checklist scores were significantly less likely to move from the clinical to non-clinical range for autistic participants:<br>**Percentage free of (primary) anxiety disorder:**<br>• There were no significant differences in the percentage of children free from their <u>primary anxiety</u> disorder between the autistic and non-autistic groups at post-intervention, 3-month, 1-year, or 2-year follow-ups.<br>• Participants in the autistic group were significantly less likely to be free from <u>all anxiety disorders</u> compared to those in the non-autistic group (at post-intervention, 3-month follow-up, 1-year follow-up and 2-years follow-up).<br>**Severity of anxiety disorders (ADIS-C/P):**<br>• No significant difference in total anxiety severity scores between the autistic and non-autistic groups across assessments (Cohen’s $d = 0.34$, $t = 1.87$, $p = 0.063$).<br>• The decrease in the total severity of anxiety disorders (slope) did not significantly differ between the autistic and non-autistic groups.<br>**Anxiety symptoms (SCARED-71):**<br>• The autistic group had higher levels of anxiety symptoms than the non-autistic group across assessments according to parent reports (Cohen’s $d = 0.46$) but not child reports (Cohen’s $d = 0.13$).<br>• There was no significant difference in SCARED-71 scores between the autistic and non-autistic groups at post-intervention ($d = 0.00$, $SE = 0.09$, $p > 0.05$). However, there was significantly smaller improvement in SCARED-71 scores in the autistic group at 3-month (Cohen’s $d = 0.23$, $SE = 0.12$, $p < 0.05$), 1-year (Cohen’s $d = 0.37$, $SE = 0.13$, $p < 0.01$), and 2-year (Cohen’s $d = 0.28$, $SE = 0.14$, $p < 0.05$) follow-up time-points.<br>• There was a significant difference in SCARED-71 scores between the autistic and non-autistic group at post-intervention between respondents (Cohen's $d = 0.39$, $p < 0.01$) but not at other follow-up time points (Cohen's $d$ ranging from 0.03 to 0.09, all $p > 0.05$).<br>Children in the autistic group reported less improvement at post-intervention (Cohen's $d$ at Time 1 = -0.38) compared with the children in the non-autistic group (Cohen's $d$ for Time 1 = -0.79) and compared with their parents (Cohen's $d$ at Time 1 = -0.85).<br>**Health-related quality of life (EuroQol 5-Dimension questionnaire):**<br>There were significant interactions between group and all time points ($p < 0.05$ at post-intervention and 2-year follow-ups, $p < 0.01$ at 3-month and 1-year follow-ups). EuroQol 5-Dimension questionnaire scores improved less in the autistic group (Cohen's $d$ ranging between 0.39 and 0.56) compared to the non-autistic group (Cohen's $d$ ranging between 0.77 and 0.98).<br>**Child Behavior Checklist:**<br>The autistic group had higher internalising (Cohen's $d = 0.54$, $p < 0.001$), externalising (Cohen's $d = 0.70$, $p < 0.001$) and other problem behaviour (Cohen's $d = 0.86$, $p < 0.001$) scores compared with the non-autistic group across timepoints.<br>There were no significant differences in changes in Child Behavior Checklist internalising, externalising, or other problem behaviour scores at post-intervention or follow-up timepoints between the autistic and non-autistic groups (all $p > 0.05$), except for Child Behavior Checklist internalising scores at 1-year follow-up (Cohens's $d = 0.36$, $p < 0.01$), which were significantly more decreased in the non-autistic group (Cohen's $d = -0.92$) compared to the autistic group (Cohen's $d = -0.62$).<br>**Normative comparisons** | and mental health symptoms: Yes<br>5. Adaptations to data collection: Partially<br>6. Intervention encouraged masking: No<br>7. Neurodiversity-affirming language used throughout: Partially<br>8. Alignment with priorities and values of autistic communities: Yes |
|  |  | • Parent-reported anxiety scores (SCARED-71) (at 3-month, 1-year, and 2-year follow-ups) in the autistic group were significantly less likely to be changed from clinical to non-clinical scores compared to the non-autistic group, but no significant difference post-intervention (p values not stated).<br>• Child-reported anxiety symptoms (SCARED-71) in the autistic group were only significantly less likely to be changed from clinical to non-clinical scores compared to the non-autistic group at post-intervention, not at 3-month, 1-year or 2-year follow-ups (p values not reported).<br>• Parents reported that internalising problems (Child Behavior Checklist) were significantly less likely to be changed from clinical to non-clinical scores in the autistic group compared to the non-autistic group at all time points (post-intervention, 3-month, 1-year, and 2-year follow-ups) (p values not stated). |  |
| **Dialectical behaviour therapy (DBT)** |  |  |  |
| **CYP** |  |  |  |
| **Phillips 2024 (55)**<br><u>Study design:</u><br>Cohort study (pre-post) | **Sample and setting:** Adolescents with 'emerging borderline personality disorder' receiving a comprehensive DBT programme at a specialist CAMHS DBT service.<br>**Comparison:** Participants were not systematically screened or assessed for autism.<br><u>Autistic group:</u> Had an autism diagnosis on clinical record.<br><u>Non-autistic group:</u> No diagnosis of autism on the clinical record.<br>**Analysis methods:** Independent samples t-tests and Mann-Whitney U tests investigated autistic and non-autistic group differences in | There were no significant differences between the autistic and non-autistic groups in pre-post changes in BPD symptoms, emotional regulation, depression symptoms, anxiety symptoms, frequency of self-harming behaviours, number of inpatient bed days, number of A&E attendances, or changes in education or work status:<br>After applying Bonferroni corrections to control for multiple comparisons, there were <u>no significant differences</u> between the autistic and non-autistic groups in pre-post changes in:<br>• **BPD symptoms** (Maclean Screening Instrument for Borderline Personality Disorder (MSI-BPD)): $t = 1.33$, d.f. = 150, $p = 0.18$, Cohen's $d = 0.31$<br>• **Emotional regulation**<br>○ Difficulties in Emotion Regulation Scale (DERS) Non-Acceptance subscale: $t = 0.53$, d.f. = 150, $p = 0.61$, Cohen's $d = 0.12$ | <u>Overall CASP rating:</u> Medium quality<br><u>AIRA ratings:</u><br>1. Involvement of autistic people in study design: No<br>2. Involvement of autistic people in outcome selection: No<br>3. Outcomes adapted/validated for autistic people: Partially<br>4. Risk of misinterpreting autistic traits/behaviours and mental health symptoms: Yes |
|  | pre-post symptom changes for continuous outcomes. Associations with categorical treatment outcomes were assessed using Fisher's Exact tests. Bonferroni correction was applied to control for multiple comparisons. Confounders are not accounted for in the analyses. | ○ DERS Goals subscale: $t = 0.55$, d.f. = 150, $p = 0.59$, Cohen's $d = 0.13$<br>○ DERS Impulsivity subscale: $t = 0.61$, d.f. = 150, $p = 0.55$, Cohen's $d = 0.14$<br>○ DERS Awareness subscale: $t = 0.18$, d.f. = 150, $p = 0.86$, Cohen's $d = 0.04$<br>○ DERS Strategies subscale: $t = 0.18$, d.f. = 150, $p = 0.86$, Cohen's $d = 0.04$<br>○ DERS Clarity subscale: $t = 1.53$, d.f. = 150, $p = 0.13$, Cohen's $d = 0.36$<br>○ DERS Total: $t = 0.65$, d.f. = 150, $p = 0.52$, Cohen's $d = 0.15$<br>● **Depression symptoms** (Mood and Feeling Questionnaire-YP): $t = 2.01$, d.f. = 150, $p = 0.05$, Cohen's $d = 0.47$<br>● **Anxiety symptoms:** (Screen for Child Anxiety-Related Disorders for Young People (SCARED-YP)): $t = 2.61$, d.f. = 150, $p = 0.01$, Cohen's $d = 0.61$<br>● **Self-harming behaviours** ($U = 1542.50$, $z = 0.90$, $p = 0.37$, $r = 0.07$)<br>● **Number of inpatient bed days** between the matched periods before and during treatment ($U = 1480.00$, $z = 0.57$, $p = 0.58$, $r = 0.05$)<br>● **Number of A&E attendances** before and during treatment ($U = 1614.50$, $z = 1.31$, $p = 0.19$, $r = 0.02$).<br>● **Changes in education status and work status** ($p = 0.55$) | 5. Adaptations to data collection: No<br>6. Intervention encouraged masking: Yes<br>7. Neurodiversity-affirming language used throughout: Yes<br>8. Alignment with priorities and values of autistic communities: No |
| **Radically open dialectical behaviour therapy (RO-DBT)** |  |  |  |
| **Adults** |  |  |  |
| **Cornwall 2020 (66)**<br><u>Study design:</u><br>Cohort study (pre-post without controls) | **Sample and setting:** Adults presenting with "maladaptive overcontrol" receiving radically open dialectical behaviour therapy (RO-DBT) in an NHS Community Mental Health Team.<br>**Comparison:** Autism diagnoses were determined from patients' electronic records. | In the per-protocol sample, the autistic group showed significantly greater improvements in global distress, wellbeing, problems, functioning, risk, and perceived recovery. However, in the intention-to-treat sample, these differences were no longer significant, except for functioning. Autism predicted global distress post-RO-DBT in the per-protocol analysis, but not in the intention-to-treat analysis. There was no significant | <u>Overall CASP rating:</u> Medium quality<br><u>AIRA ratings:</u><br>1. Involvement of autistic people in study design: No |
|  | <u>Autistic group:</u> Autism diagnoses from a specialist autism service or “working diagnosis” of autism whilst on the waitlist for specialist diagnostic assessment.<br><u>Non-autistic group:</u> No confirmed or ‘working’ diagnosis of autism.<br>**Analysis methods:** Stepwise regression examined factors predicting outcome score at 30 weeks in participants completing the programme to 30 weeks (per-protocol, n = 21) and the intention-to-treat sample (n = 35). Entered variables included: age, sex, initial severity (CORE global distress score at baseline), autism diagnosis, cohort, and number of sessions attended. | between-group difference in rates of meeting the CORE clinical recovery threshold after treatment:<br>**In the per-protocol sample (autistic group n = 11, non-autistic group n = 10):** Participants with a confirmed or working diagnosis of autism showed clinically reliable improvement and significantly better outcomes than non-autistic participants with respect to:<br>• CORE global distress scores (t = 3.04, d.f. = 20, p < 0.01)<br>• CORE well-being scores (t = 1.93, d.f. = 20, p = 0.03)<br>• CORE problems scores (t = 3.28, d.f. = 20, p < 0.01)<br>• CORE functioning scores (t = 2.47, d.f. = 20, p = 0.01)<br>• CORE risk scores (t = 1.96, d.f. = 20, p = 0.03)<br>• Questionnaire about the Perception of Recovery (5 = 2.29, d.f. = 20, p = 0.02)<br>**In the intention-to-treat sample (autistic group n = 18, non-autistic group n = 17):**<br>- The change in CORE functioning scores was significantly larger (t = 2.00, d.f. = 34, p = 0.03) in the autistic group compared to the non-autistic group.<br>- There were no other significant differences between groups in changes in:<br>• CORE global distress scores (t = 1.57, d.f. = 34, p = 0.06)<br>• CORE well-being scores (t = 0.26, d.f. = 34, p = 0.40)<br>• CORE problems scores (t = 1.46, d.f. = 34, p = 0.08)<br>• CORE risk scores (t = 0.62, d.f. = 34, p = 0.27)<br>• Questionnaire about the Perception of Recovery (5 = 1.46, d.f. = 31, p = 0.08)<br>**Clinical recovery threshold (CORE global distress score < 1.0):** At the end of treatment, there was no significant difference in the proportion of autistic and non-autistic participants who met the clinical recovery threshold ($\chi^2 = 2.01$, d.f. = 1, p = 0.16). | 2. Involvement of autistic people in outcome selection: No<br>3. Outcomes adapted/validated for autistic people: No<br>4. Risk of misinterpreting autistic traits/behaviours and mental health symptoms: Yes<br>5. Adaptations to data collection: No<br>6. Intervention encouraged masking: Yes<br>7. Neurodiversity-affirming language used throughout: No<br>8. Alignment with priorities and values of autistic communities: No |
|  |  | **Reliable improvement in CORE global distress scores (reduction in score >0.5):** Autistic participants were significantly more likely to have a reliable improvement in CORE global distress scores in the per-protocol (73% vs 10%, $\chi^2 = 8.21$, d.f. = 3, $p = 0.04$) but not the intention-to-treat sample (56% vs 18%, $\chi^2 = 4.58$, d.f. = 3, $p = 0.21$).<br>**Predictors of final CORE global distress score:** In the stepwise regression, initial severity and autism diagnosis accounted for a highly significant proportion of the variation in final outcome score. Autism diagnosis significantly predicted outcome in the per-protocol sample ($\beta = 0.75$, standard error = 0.23, 95% CI: 0.34-1.13, $t = 3.27$, $p < 0.01$), but not in the intention-to-treat sample ($\beta = 0.37$, standard error = 0.21, 95% CI: 0.01-0.73, $F = 1.76$, $p = 0.09$; $p = 0.09$). |  |
| **Cognitive Remediation and Emotional Skills Training (CREST)** |  |  |  |
| **Adults** |  |  |  |
| **Adamson 2018 (56)**<br><u>Study design:</u><br>Cohort study (pre-post comparative case series) | **Sample and setting:** Adults diagnosed with anorexia nervosa receiving CREST (group or individual) in a specialist inpatient eating disorder unit.<br>**Comparison:**<br><u>High autistic traits (HAT) group:</u> AQ-10 score of 6+<br><u>Low autistic traits (LAT) group:</u> AQ-10 score of <6<br>**Analysis methods:** Linear mixed-effects models used to analyse overall response in both individual and group CREST interventions and assess interaction with autism symptoms. BMI was included as a covariate. | There were no significant differences between the HAT and LAT groups in pre-post changes in social anhedonia or alexithymia for either the CREST group or individual interventions.<br>**CREST group intervention**<br>In the CREST group intervention group, there were <u>no significant differences</u> between the HAT and LAT groups in the change from pre- to post-intervention in Social Anhedonia Scale ($F(1, 36) = 0.27$, $p = 0.61$) or Toronto Alexithymia Scale ($F(1, 34) = 0.85$, $p = 0.36$). BMI was not a significant covariate in any of the analyses.<br>**CREST individual intervention**<br>In the CREST individual intervention group, there were <u>no significant differences</u> between HAT and LAT groups in the change in Social Anhedonia Scale ($F(1, 33) = 2.99$, $p = 0.09$) or Toronto Alexithymia Scale ($F(1, 34) = 0.10$, $p = 0.75$) scores from pre- to post-intervention. BMI was not a significant covariate in any of the analyses. | <u>Overall CASP rating:</u> Low quality<br><u>AIRA ratings:</u><br>1. Involvement of autistic people in study design: No<br>2. Involvement of autistic people in outcome selection: No<br>3. Outcomes adapted/validated for autistic people: No<br>4. Risk of misinterpreting autistic traits/behaviours and mental health symptoms: Yes |
|  |  |  | 5. Adaptations to data collection: No<br>6. Intervention encouraged masking: No<br>7. Neurodiversity-affirming language used throughout: Yes<br>8. Alignment with priorities and values of autistic communities: Yes |
| **Family Therapy for Anorexia Nervosa + CBT for low mood, anxiety, or OCD when indicated** |  |  |  |
| **CYP** |  |  |  |
| **Stewart 2017 (91)**<br><u>Study design:</u><br>Cohort study (pre-post) | **Sample and setting:** CYP with high and low autistic traits receiving Family Therapy for Anorexia Nervosa (FT-AN) (and additional CBT for low mood, anxiety, or OCD where indicated) at a specialist outpatient eating disorder service.<br>**Comparison:**<br><u>High autistic trait (HAT) group:</u> Score of $\geq 30$ on the AQ-50 (completed by caregiver) or a score $<16$ on the Social Aptitude Scale.<br><u>Low autistic traits (LAT) group:</u> score of $<30$ on AQ-50 or $\geq 16$ on the Social Aptitude Scale.<br>**Analysis methods:** Group differences in physical recovery were assessed using chi-square tests based on Morgan Russell criteria. Pearson and Spearman correlations evaluated relationships between autistic traits (AQ-50, SAS, Development and Well-Being Assessment [Autism Spectrum Disorder module]) and change in eating- | There were no significant differences between the HAT and LAT groups in Morgan Russell physical health outcomes or paediatric medical admissions. However, the HAT group had significantly higher rates of psychiatric ward and intensive day patient programme admissions when defined by AQ scores, but not when defined by SAS scores.<br>**Physical outcomes by autistic traits**<br>Morgan Russell physical health outcomes did not significantly differ between young people with high versus low autistic traits as measured by the AQ or Social Aptitude Scale (all $\chi^2 < 3.02$, $p > .05$), indicating comparable physical recovery across groups.<br>**Treatment augmentation (acute service use):**<br>There were significantly higher rates of psychiatric ward admission ($\chi^2 = 8.68$, $df = 1$, $p < 0.005$) in the HAT group compared to the LAT group, defined by AQ-50 scores, but not when defined by Social Aptitude Scale scores ($\chi^2 < .2.6$, $df = 1$, all $p > 0.05$).<br>There were significantly higher rates of admission to an intensive day patient programme in the HAT group compared to the LAT group, defined by AQ-50 scores ($\chi^2 = 6.21$, $df = 1$, $p < 0.025$) but not when defined by Social Aptitude Scale scores ($\chi^2 < .2.6$, $df = 1$, all $p > 0.05$). | <u>Overall CASP rating:</u> Low quality<br><u>AIRA ratings:</u><br>Involvement of autistic people in study design: No<br>Involvement of autistic people in outcome selection: No<br>Outcomes adapted/validated for autistic people: Partially<br>Risk of misinterpreting autistic traits/behaviours and mental health symptoms: Yes<br>Adaptations to data collection: No<br>Intervention encouraged masking: Can't tell<br>Neurodiversity-affirming language used throughout: Yes |
|  | disorder, depression, anxiety, quality-of-life, and OCD-type symptoms. | There was no significant difference in paediatric medical admissions between the HAT and LAT groups when defined by AQ-50 scores ($\chi^2 = 0.27$, $df = 1$, $p > 0.05$) or Social Aptitude Scale scores ($\chi^2 < .2.6$, $df = 1$, all $p > 0.05$). | 8. Alignment with priorities and values of autistic communities: Yes |
| **NHS Talking Therapies services** |  |  |  |
| **Adults** |  |  |  |
| **El Baou 2023 (69)**<br><u>Study design:</u><br>Retrospective matched cohort study | **Sample and setting:** Adults who received at least two sessions of psychological therapy in any NHS Talking Therapies service (formerly known as Improving Access to Psychological Therapies services) in England between 2012-19 and had a scores of 10+ on the PHQ-9 or 8+ on the GAD-7.<br>**Comparison:**<br><u>Autistic group:</u> People with an autism diagnosis recorded in Hospital Episode Statistics or Mental Health Services Data Set (F84.0, F84.1, and F84.5). Note: this would not include people with a historic PDD-NOS code. Autism diagnosis codes were considered regardless of their temporal relationship to NHS Talking Therapies referral.<br><u>Non-autistic group:</u> People without an autism diagnosis code recorded in Hospital Episode Statistics or Mental Health Services Data Set.<br>**Analysis methods:**<br>Logistic regression models were used for primary outcomes and linear regression models for secondary outcomes, in this order: | **Primary outcomes**<br>In all models, autistic adults were less likely to meet improvement and recovery criteria (measured with PHQ-9 and GAD-7) and were more likely to show a deterioration in their symptoms than adults without identified autism (all $p < 0.0001$). Smaller improvements in anxiety, depression, and functioning (WSAS) were also observed in the autistic group in all models (all $p < 0.0001$).<br><u>Model 3: Full sample (adjusted)</u><br>Reliable improvement ($n = 1,927,265$), OR = 0.76 (95% CI: 0.72 to 0.79), $p < 0.0001$<br>Reliable recovery ($n = 1,927,265$), OR = 0.71 (95% CI: 0.68 to 0.75), $p < 0.0001$<br>Reliable deterioration ($n = 1,927,265$), OR = 1.34 (95% CI: 1.24 to 1.45), $p < 0.0001$<br>PHQ-9 change ($n = 1,927,265$), adjusted mean difference = -1.00 (95% CI: -1.12 to -0.89), $p < 0.0001$<br>GAD-7 change ($n = 1,927,265$), adjusted mean difference = -0.76 (95% CI: -0.86 to -0.67), $p < 0.0001$<br>WSAS change ($n = 1,265,086$), adjusted mean difference = -1.20 (95% CI: -1.43 to -0.95), $p < 0.0001$<br><u>Model 4: Propensity score matched (adjusted)</u><br>Reliable improvement ($n = 17186$), OR = 0.75 (95% CI: 0.70 to 0.80), $p < 0.0001$<br>Reliable recovery ($n = 17186$), OR = 0.68 (95% CI: 0.63 to 0.73), $p < 0.0001$ | <u>Overall CASP rating:</u> High quality<br><u>AIRA ratings:</u><br>Involvement of autistic people in study design: No<br>Involvement of autistic people in outcome selection: No<br>Outcomes adapted/validated for autistic people: Partially<br>Risk of misinterpreting autistic traits/behaviours and mental health symptoms: Yes<br>Adaptations to data collection: No<br>Intervention encouraged masking: Can't tell<br>Neurodiversity-affirming language used throughout: Yes<br>Alignment with priorities and values of autistic communities: Yes |
|  | 1) <u>Model 1</u>: Included autistic group or comparison group<br>2) <u>Model 2</u>: Additionally adjusted for clinical and sociodemographic covariates<br>3) <u>Model 3</u>: Additionally adjusted for NHA Talking Therapies treatment factors<br>4) <u>Model 4</u>: Re-run using a propensity score matched sample | • Reliable deterioration (n = 17186), OR = 1.34 (95% CI: 1.18 to 1.48), p < 0.0001<br>• PHQ-9 change (n = 17186), adjusted mean difference = -0.97 (95% CI: -1.15 to -0.80)<br>• GAD-7 change (n = 17186), adjusted mean difference = -0.80 (95% CI: -0.96 to -0.65), p < 0.0001<br>• WSAS change (n = 10750), adjusted mean difference = -1.01 (95% CI: -1.37 to -0.66), p < 0.0001<br>**Subgroup analyses**<br>**Socioeconomic deprivation:**<br>• Whilst low socioeconomic deprivation was associated with improved outcomes in the non-autistic group, adults in the autistic group showed similar (poorer) outcomes regardless of level of deprivation.<br>• In high levels of socioeconomic deprivation, autistic adults had lower odds of reliable improvement than the non-autistic group (OR = 0.89, 95% CI: 0.79 to 1.00) but also had slightly lower odds in the lowest socioeconomic deprivation category (OR = 0.68, 95% CI: 0.58 to 0.79).<br>**Employment:**<br>• If employed, autistic adults had significantly lower odds of reaching reliable improvement after therapy than employed non-autistic adults (OR = 0.65, 95% CI: 0.58 to 0.72).<br>• If unemployed and seeking work, there was no significant difference between autistic and non-autistic adults in odds of reaching reliable improvement (OR = 0.95, 95% CI: 0.83 to 1.07).<br>Subgroup analyses showed that the strength of the differences in outcomes between autistic and non-autistic groups did not appear to be affected by other factors (e.g. gender, taking psychotropic medications, diagnosis of depression vs anxiety, presence of a long-term condition). |  |
| Counselling psychotherapy at a university counselling service |  |  |  |

| **Adults** |  |  |  |
| --- | --- | --- | --- |
| **Anderberg 2017 (57)**<br><u>Study design:</u><br>Retrospective cohort study | **Sample and setting:** Adults receiving counselling psychotherapy at a University Counselling and Psychological Services Centre.<br>**Comparison:** Grouped based on case note review by two independent raters into the following categories:<br>• <u>Reported autism:</u> if an autism diagnosis is reported<br>• <u>Possible autism:</u> if strongly suspected to be autistic or was pursuing an autism assessment<br>• <u>Non-autistic:</u> no autism diagnosis, not strongly suspected to be autistic, and not pursuing an autism assessment<br>**Analysis methods:**<br>• Multiple regression compared change in Outcome Questionnaire-45 scores between the ‘reported autism’, ‘possible autism’ and ‘non-autistic’ groups, controlling for gender, marital status, country of residence, ethnicity, year of therapy, and age.<br>• Chi squared tests analysed differences in rates of reliable improvement and deterioration in OQ-45 scores between groups. Reliable improvement = 14-point decrease in OQ-45 scores from intake. Reliable deterioration = 14-point increase in OQ-45 scores from intake.<br>• Explored change trajectories using a repeated measures mixed model with total OQ-45 score as the response<br>variable and diagnosis, session number, and the interaction between these as explanatory variables. Controlled for covariates including gender, marital status, country of citizenship, ethnicity, and age. | There were no significant differences between groups in reliable improvement in psychological functioning at the end of treatment or at any point during treatment. There was also no significant difference in rates of reliable deterioration in psychological functioning at the end of treatment, though reported autism and possible autism groups were significantly more likely to deteriorate at any point during treatment, and took longer to reach their best scores:<br>**Differences in OQ-45 outcomes between groups**<br>• There were no significant differences between the ‘reported autism’, ‘possible autism’ and ‘non-autistic’ groups in pre-post changes in OQ-45 scores ($F(2,17884) = 0.423$, $p = 0.66$), OQ-45 reliable improvement rates at any point during treatment ($\chi^2 = 0.82$, $p = 0.66$) or at the end of treatment ($\chi^2 = 3.20$, $p = 0.20$), or reliable deterioration rates at the end of treatment ($\chi^2 = 0.20$, $p = 0.90$).<br>• However, there was a significant difference in reliable deterioration rates with deterioration at any time during treatment ($\chi^2 = 10.49$, $p = 0.005$), with higher rates of deterioration in the autistic groups compared to the non-autistic group.<br>**Change trajectories:** OQ-45 change trajectories over the course of treatment differed significantly between ‘reported autism’ ($p < 0.001$) and ‘possible autism’ ($p = 0.002$) groups compared to the ‘non-autistic’ group, with those in the autistic groups taking more sessions to reach their lowest OQ-45 scores. | <u>Overall CASP rating:</u> Medium quality<br><u>AIRA ratings:</u><br>1. Involvement of autistic people in study design: No<br>2. Involvement of autistic people in outcome selection: No<br>3. Outcomes adapted/validated for autistic people: No<br>4. Risk of misinterpreting autistic traits/behaviours and mental health symptoms: Yes<br>5. Adaptations to data collection: No<br>6. Intervention encouraged masking: Can’t tell<br>7. Neurodiversity-affirming language used throughout: Yes<br>8. Alignment with priorities and values of autistic communities: Partially |

##### Cognitive Behavioural Therapy for Obsessive Compulsive Disorder

Seven studies examined outcomes following CBT for OCD (53,71,74,81,83,84,96). Two were rated low quality (81,84) and five medium quality (53,71,74,83,96). All studies, with the exception of Jassi et al. (2023) (74), had small overall sample sizes (n < 200) and/or small autistic groups (n < 50). Although Jassi et al. (2023) included a larger sample, more than 50% of outcome data were missing. Findings from these studies should therefore be interpreted cautiously.

Five studies compared participants with and without an autism diagnosis (71,74,83,84,96). One compared high autistic trait (HAT) and low autistic trait (LAT) groups based on autism screening measure scores (81), and another excluded people with an autism diagnosis and compared HAT and LAT groups (53). Four studies recruited only CYP (53,71,74,83), two adults only (81,84), and one both CYP and adults (96). In four studies, the intervention consisted of only CBT for OCD (71,83,84,96), whereas in the other three studies, medication could be offered in combination with CBT for OCD (53,74,81).

##### Meta-analysis

Meta-analysis of three studies (81,84,96) found a significant moderate-to-large between-group difference in post-treatment OCD symptom severity measured using the Yale Brown Obsessive-Compulsive Scale (Y-BOCS) (Hedges’ g = −0.70, 95% CI −1.06 to −0.33, p < 0.001), with low heterogeneity (I^2^ = 8.88%). Pooling studies, autistic adults had higher post-treatment OCD symptom severity than non-autistic adults. Meta-analysis was limited to these three studies as they were sufficiently comparable in terms of intervention (CBT for OCD), population (adults with OCD), and reported outcomes (pre-post Y-BOCS total scores).

Other evidence suggested no significant differences between autistic and non-autistic people in change in OCD diagnostic severity (71) or depression symptoms (84) during CBT. However, there were mixed findings regarding differences between groups in change in OCD symptoms in CYP (71,74,83), post-intervention OCD symptoms in CYP (53), response rates (71,74,83), remission rates (71,74,83,96), and functioning (71,96); some studies reported no significant differences between groups, whereas others found smaller improvements in the autistic group.

##### Cognitive behavioural therapy (CBT) for anxiety

Three studies examined outcomes of CBT for anxiety disorders in CYP with and without autism diagnoses in outpatient settings. Two of the studies (72,97) analysed outcomes from the same cohort, while the third study (60) examined a distinct sample. Two were rated high quality (72,97), and one medium quality (60). All three studies included moderately sized overall samples (n ≥ 200), with autistic subgroups exceeding 75 participants. Hagen et al. (2025) (72) did not report rates of missing outcome data, however, as the study analysed the same cohort as van Steensel et al. (2015) (97), which reported substantial levels of participant dropout, results from these two studies should be interpreted cautiously.

Cervin et al. (2023) conducted a secondary analysis of pooled data from five RCTs and reported mixed findings regarding differences between autistic and non-autistic CYP in anxiety and functioning (60). Post-CBT, after controlling for age, sex, baseline anxiety severity, and trial, autistic CYP had significantly more physical symptoms of anxiety and significantly higher impact of anxiety symptoms on family life. However, there were no significant between-group differences in anxiety symptom frequency or intensity, avoidance, or impact of anxiety on school/social life.

Van Steensel et al. (2015) measured participants’ outcomes post-intervention, and at 3-month, 1-year, and 2-year follow-up timepoints (97). There were no significant differences between autistic and non-autistic CYP in the proportion free from their primary anxiety disorder, change in overall anxiety severity, or internalising, externalising, or other problem behaviours at most timepoints. There was no significant difference between autistic and non-autistic groups in changes in anxiety symptoms from pre-to post-intervention, but improvements in anxiety symptoms at longer follow-up time points were significantly smaller in the autistic group. A significantly smaller proportion of autistic CYP were free from all anxiety disorders at all time points, and there were significantly smaller improvements in quality of life in the autistic group. Findings for the likelihood of anxiety reducing from the clinical to non-clinical range at different time points were mixed, depending on informant.

Hagen et al. (2025), which analysed the same cohort as van Steensel et al. (2015) (97), found no significant differences between autistic and non-autistic CYP in changes in family functioning from pre-to post-intervention or from post-intervention to the 3-month, 1-year, and 2-year follow-ups (72). They also reported that anxiety levels were significantly higher in the autistic group than the non-autistic group across time points (72).

Taken together, these studies mostly reported no significant differences between autistic and non-autistic CYP in changes in anxiety immediately following CBT for anxiety, although some evidence suggested significantly smaller improvements among autistic CYP at longer-term follow-up. Findings for functioning were also mixed. There was some evidence of smaller improvements in quality of life among autistic CYP. However, these findings should be interpreted cautiously, as two of the studies had high attrition rates, which may have increased the risk of bias.

##### Dialectical behaviour therapy (DBT)

One medium-quality study (55) examined outcomes following comprehensive DBT for adolescents with “emerging borderline personality disorder” (BPD) – a contested term sometimes used to describe clinically significant features of BPD in CYP under 18 – with and without an autism diagnosis. On the AIRA, the intervention was rated as potentially encouraging masking of autistic traits (e.g., modifying communication style to confirm to neurotypical expectations), a limitation that was also acknowledged by the authors. The study reported no significant differences between the autistic and non-autistic group in pre-post changes in BPD symptoms, difficulties in emotional regulation, depressive symptoms, anxiety symptoms, self-harming behaviours, number of inpatient days or Accident and Emergency (ACE) attendances before or during treatment, or changes in education status and work status. Within-group analyses suggested significant pre-post improvement in BPD symptoms and some domains of emotional regulation for the autistic group, while other within-group changes were non-significant. However, the autistic group was small (n = 27) and the authors’ power calculation indicated that analyses were underpowered to detect small-to medium-sized effects; as such, these null findings should be interpreted with caution.

##### Radically open dialectical behaviour therapy (RO-DBT)

One medium-quality study (66) compared outcomes of RO-DBT delivered within a Community Mental Health Team to adults presenting with “maladaptive overcontrol” with a confirmed or “working” diagnosis of autism to those without. RO-DBT is an adaptation of DBT designed to reduce excessive self-control by using strategies to improve emotional expression and behavioural flexibility, with the aim of enhancing social signalling and, in turn, social connectedness (66). On the AIRA, RO-DBT was rated as potentially encouraging masking autistic traits (e.g. differences in communication style, preferences for predictability, or repetitive behaviours) through its emphasis on increasing behavioural flexibility and emotional expressiveness. The overall sample size in the intention-to-treat analysis was small (n = 48, including 23 autistic participants), and the results should therefore be interpreted with caution.

In the intention-to-treat sample, pre-post changes did not differ significantly between the autistic and non-autistic groups for most outcomes, including perceived recovery, Clinical Outcomes in Routine Evaluation (CORE) domain scores (global distress, wellbeing, problems, and risk), reliable improvement in CORE global distress scores (reduction of >0.5), and the proportion meeting the clinical recovery threshold (CORE global distress <1.0) at treatment completion. The exception was the CORE functioning domain, which showed significantly greater improvement in the autistic group. Autism diagnosis was not a significant predictor of final CORE global distress scores in the intention-to-treat multivariate model.

##### Cognitive Remediation and Emotional Skills Training (CREST)

One study (56), rated low quality, evaluated CREST for adults with anorexia nervosa in a specialist inpatient eating disorder unit, comparing outcomes for HAT and LAT groups based on the Autism Quotient-50 (AQ-50) autism screening measure scores. There were no significant differences between the HAT and LAT groups in either the CREST group or individual interventions in pre-post changes in social anhedonia or alexithymia (difficulty recognising, understanding, and describing emotions). However, given the small overall sample (n ≤ 66) and autistic group sizes in the group (n = 21) and individual (n = 21) CREST intervention conditions, these findings should be interpreted with caution.

##### Family Therapy for Anorexia Nervosa (FT-AN) with Cognitive Behavioural Therapy (CBT) for low mood, anxiety, or obsessive-compulsive disorder (OCD) when indicated

One study (91), rated low quality, examined outcomes from FT-AN with CBT for low mood, anxiety, or OCD where indicated for female CYP. HAT and LAT groups were defined using cut-offs on the AQ-50 or Social Aptitude Scale (SAS). The overall sample was relatively large (n = 409) but the HAT subgroups were relatively small (AQ-50: n = 20; SAS: n = 52), and there were substantial rates of missing data for some outcomes, so findings should be interpreted with caution.

There were no significant differences between the HAT and LAT groups in Morgan-Russell physical health outcomes or paediatric medical admissions. Rates of psychiatric ward and intensive day patient programme admissions were significantly higher in the HAT than the LAT group when defined by AQ-50 scores, but not when defined by SAS scores.

##### NHS Talking Therapies

Two studies examined outcomes of NHS Talking Therapies (a network of services delivering brief psychological therapies nationwide in England) for adults with and without autism diagnoses receiving treatment for depression and/or anxiety (69,89). One was rated high quality (69) and one low quality (89).

Proctor (2023) evaluated outcomes within a single NHS Talking Therapies service and reported descriptive comparisons between autistic and non-autistic service users but did not conduct statistical significance testing (89). El Baou et al. (2023) conducted a retrospective matched cohort study analysing a national English dataset (MODIFY) (69). The study included a very large overall sample size (n = 1927265), with large autistic (n = 8761) and non-autistic (n = 1918504) groups. The study was well powered to detect small-to-moderate between-group differences and used multivariable adjustment and propensity score matching to reduce confounding.

El Baou et al. (2023) reported that autistic adults were consistently significantly less likely than non-autistic adults to achieve ‘reliable improvement’ in anxiety or depression (defined as a reduction of ≥6 points on the Patient Health Questionnaire-9 [PHQ-9] or ≥4 points on the Generalized Anxiety Disorder-7 [GAD-7]) or ‘reliable recovery’ (reliable improvement plus scores below thresholds for caseness on the PHQ-9 and GAD-7 at the end of treatment). Autistic adults were also significantly more likely to experience ‘reliable deterioration’ (increase of ≥6 points on PHQ-9 or ≥4 points on GAD-7) (69). Autistic adults also showed significantly smaller reductions in depressive symptoms, anxiety symptoms, and functional impairment. These disparities persisted after adjustment for sociodemographic, clinical and treatment related factors, and in propensity score-matched analyses. Subgroup analyses showed that in the non-autistic group, improved outcomes were associated with employment and lower socioeconomic deprivation, whereas this was not the case for autistic adults.

##### Counselling psychotherapy at a University Counselling Service

One study, rated medium quality, evaluated outcomes for adults receiving counselling psychotherapy at a university counselling service (57). This study was conducted within the specific cultural and institutional context of a religiously affiliated university in Utah, USA, which may limit the generalisability of findings to other settings. Participants were grouped as ‘reported autism’ (reported diagnosis; n = 76), ‘possible autism’ (suspected to be autistic or pursuing assessment; n = 91), or non-autistic (n = 21546). There were no significant between-group differences in pre-post changes in overall psychological functioning (Outcome Questionnaire-45; OQ-45) or in rates of reliable improvement in psychological functioning.

However, ‘reliable deterioration’ at any point during treatment was significantly more common in both autistic groups compared to the non-autistic group. Autistic participants took significantly longer to reach their best psychological functioning scores and attended more courses of therapy and more sessions within their first course. Overall, the findings suggest broadly comparable short-term symptom change but greater treatment intensity and increased risk of deterioration among autistic participants.

#### 3.4.2. Multi-component or service-level approaches

Twenty-two studies examined compared outcomes of multi-component or service-level approaches for autistic and non-autistic people. The findings below are reported by type of intervention and tabulated in Table 6.

**Table 6:**
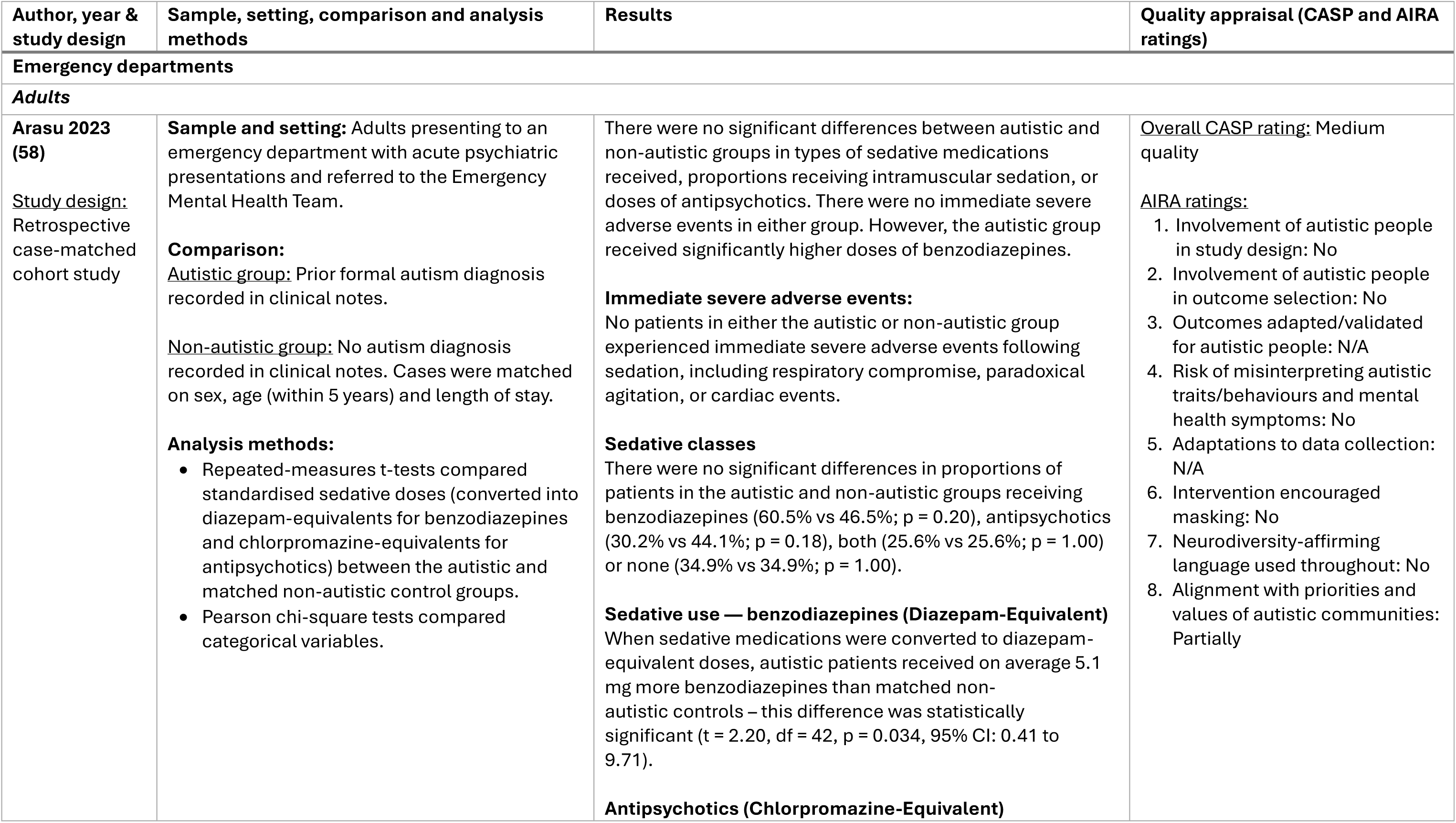
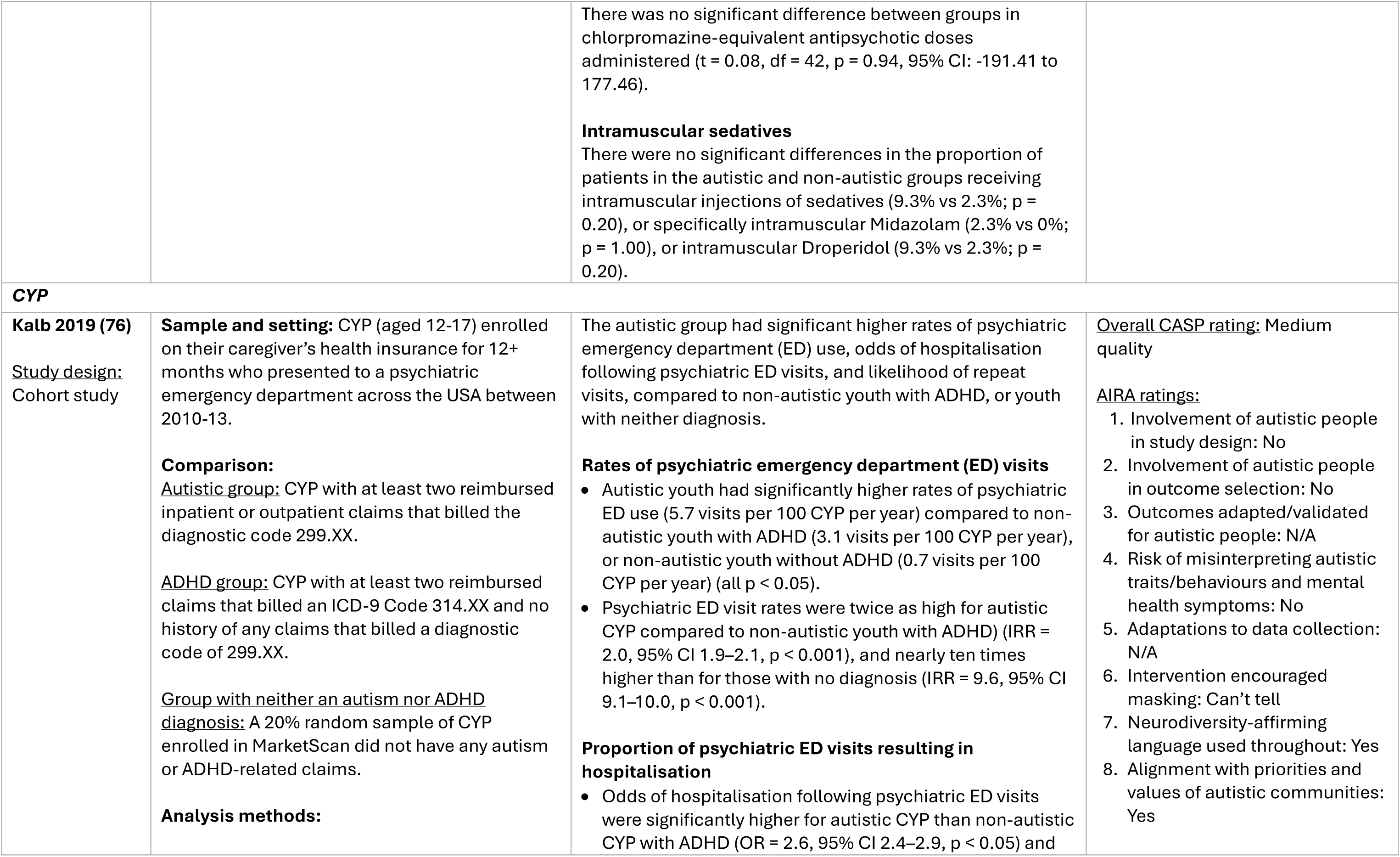
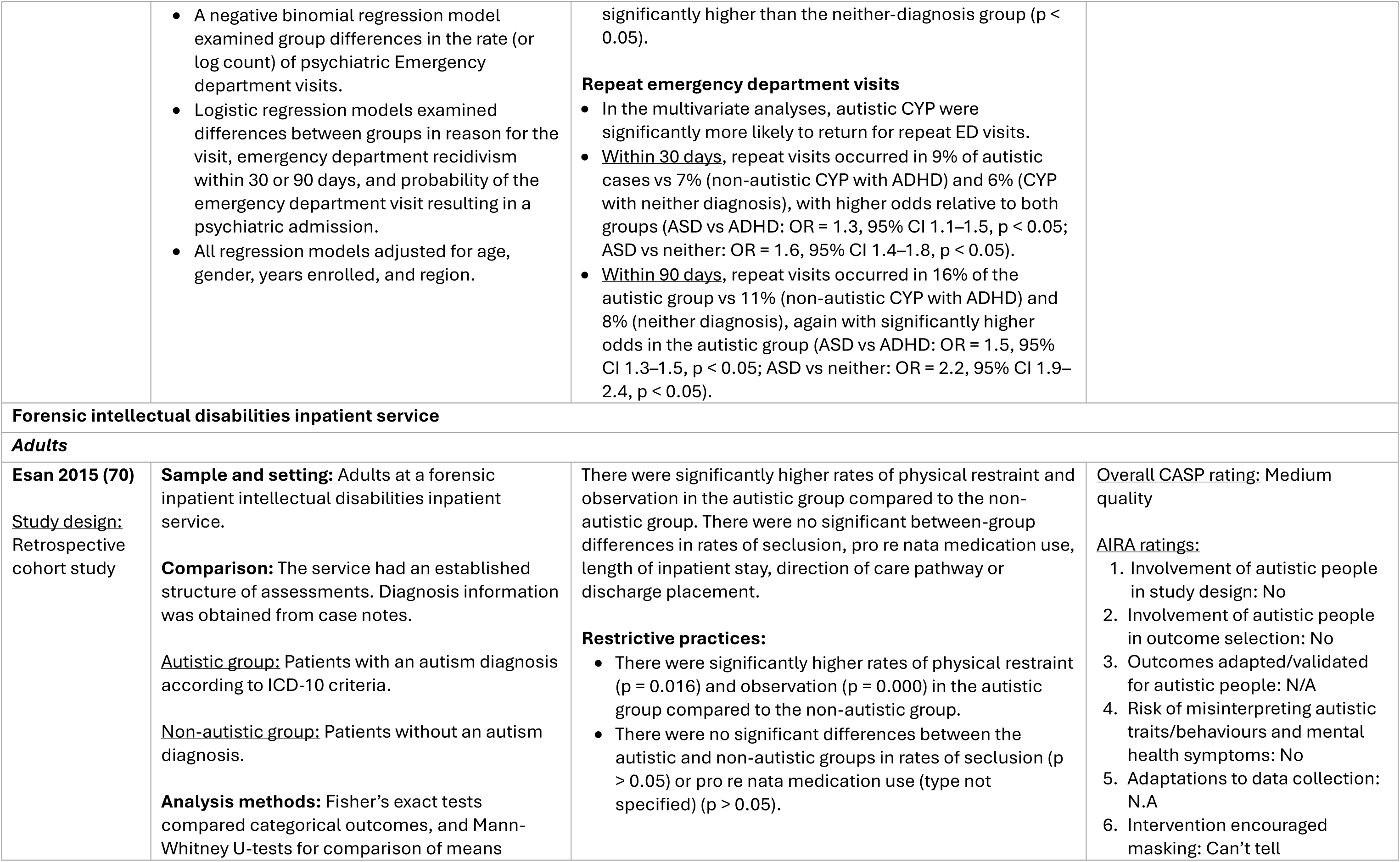
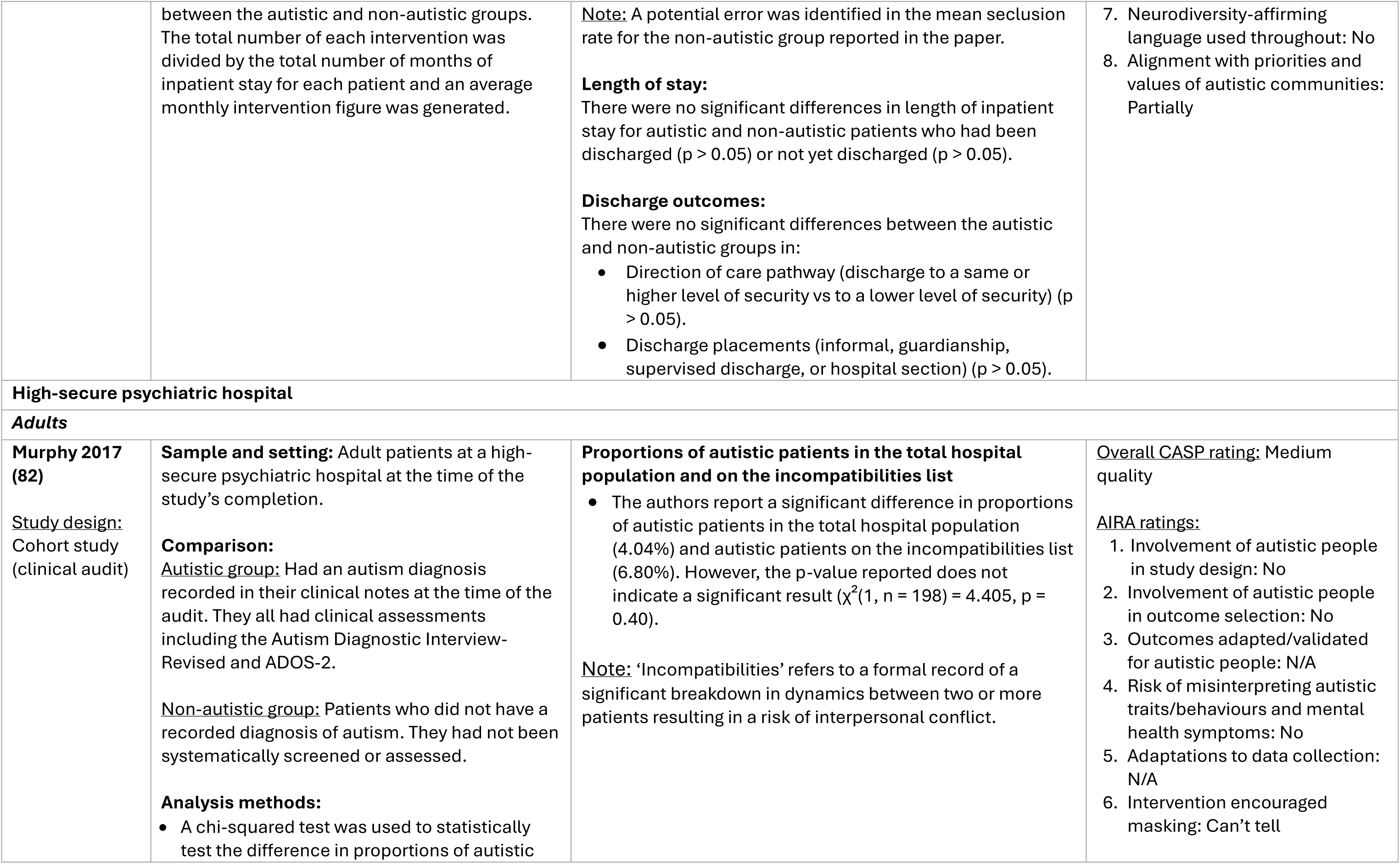
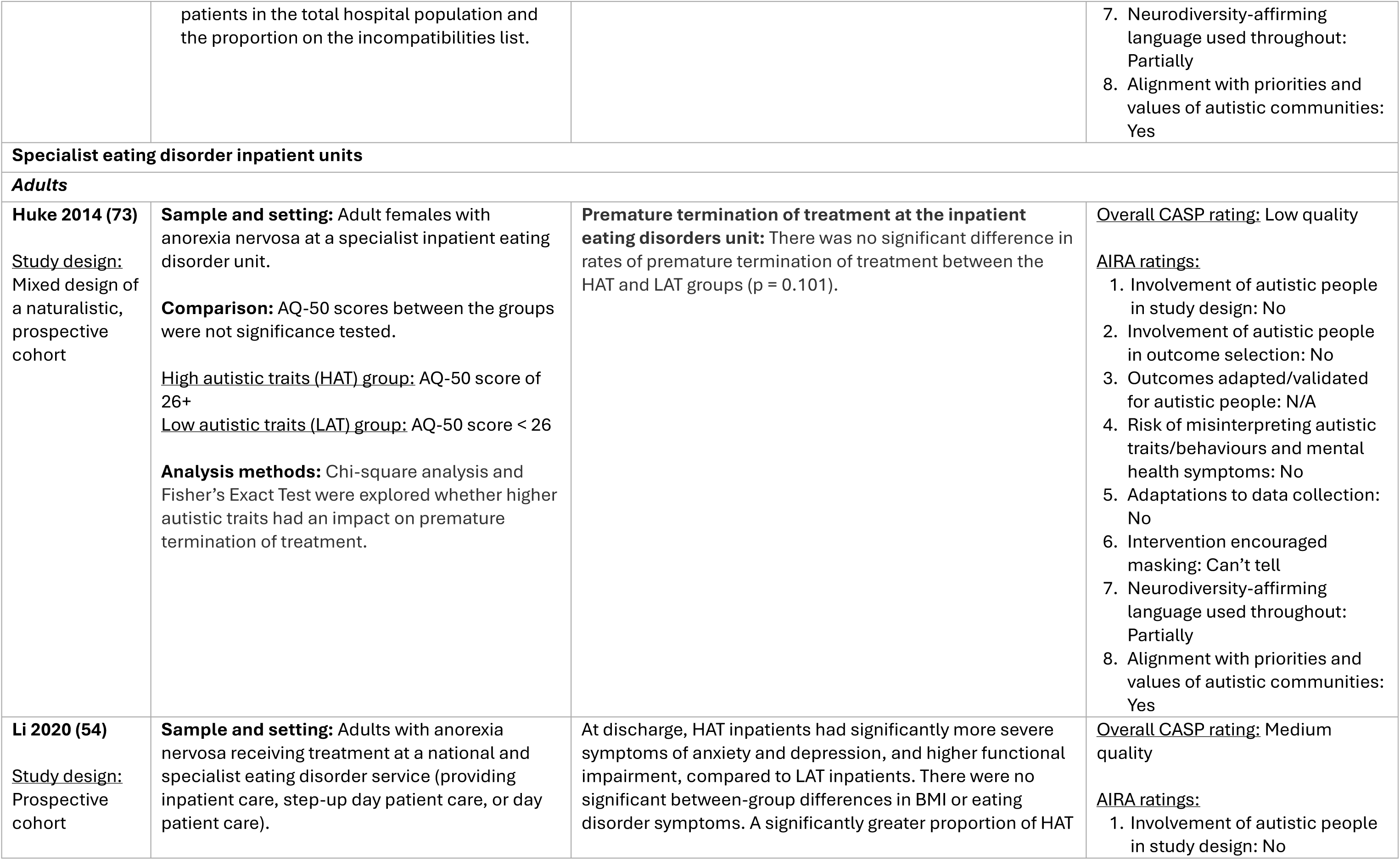
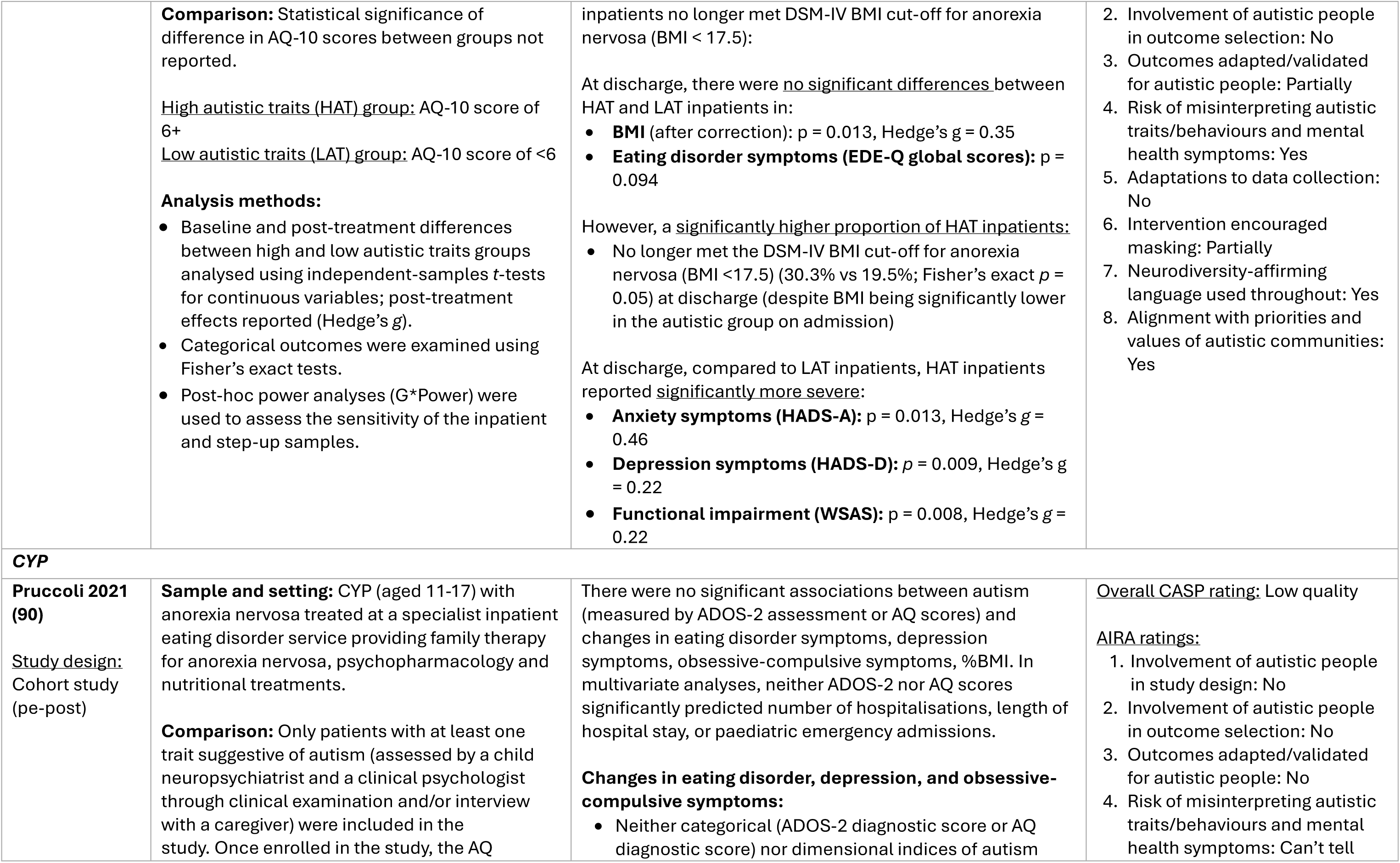
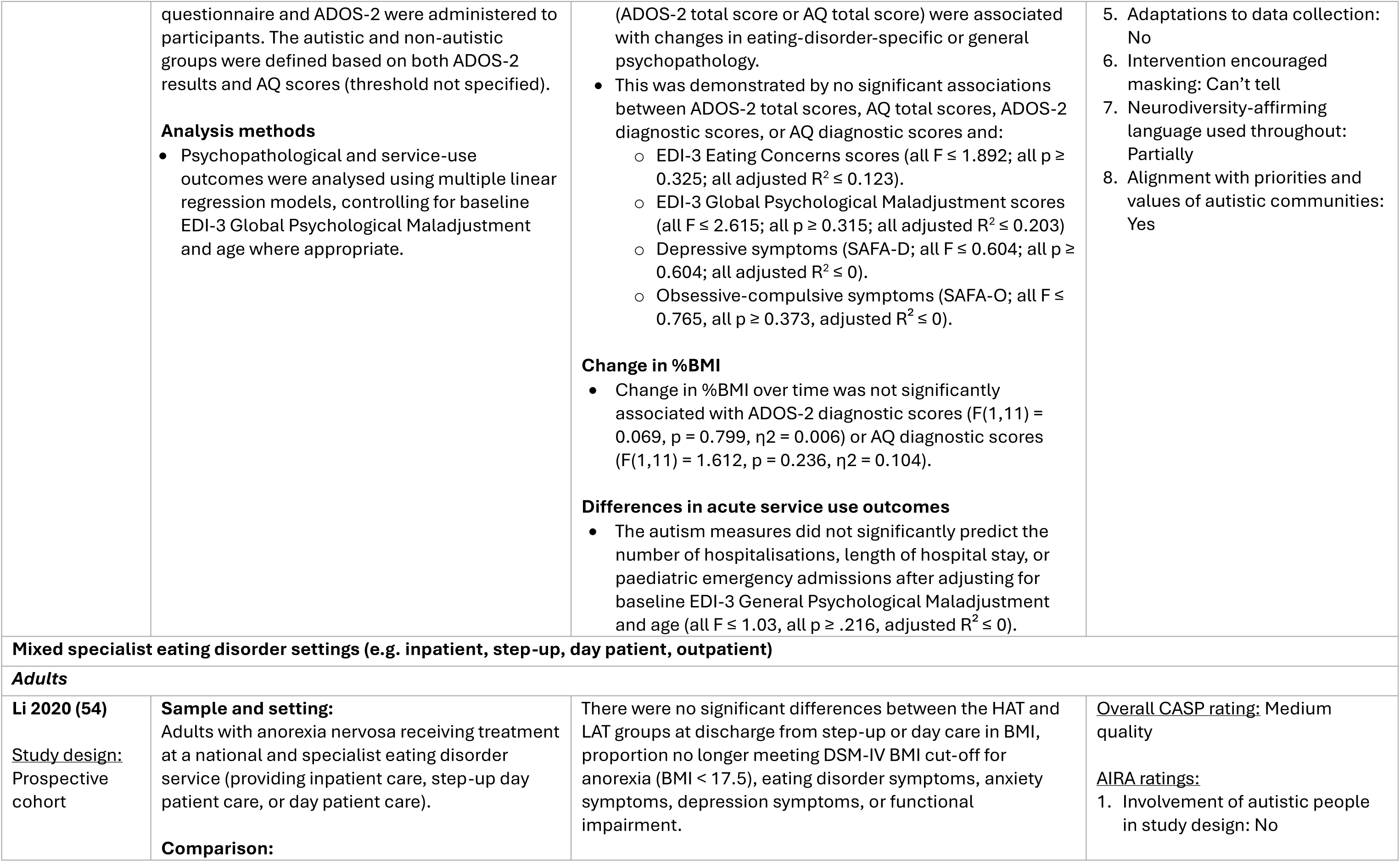
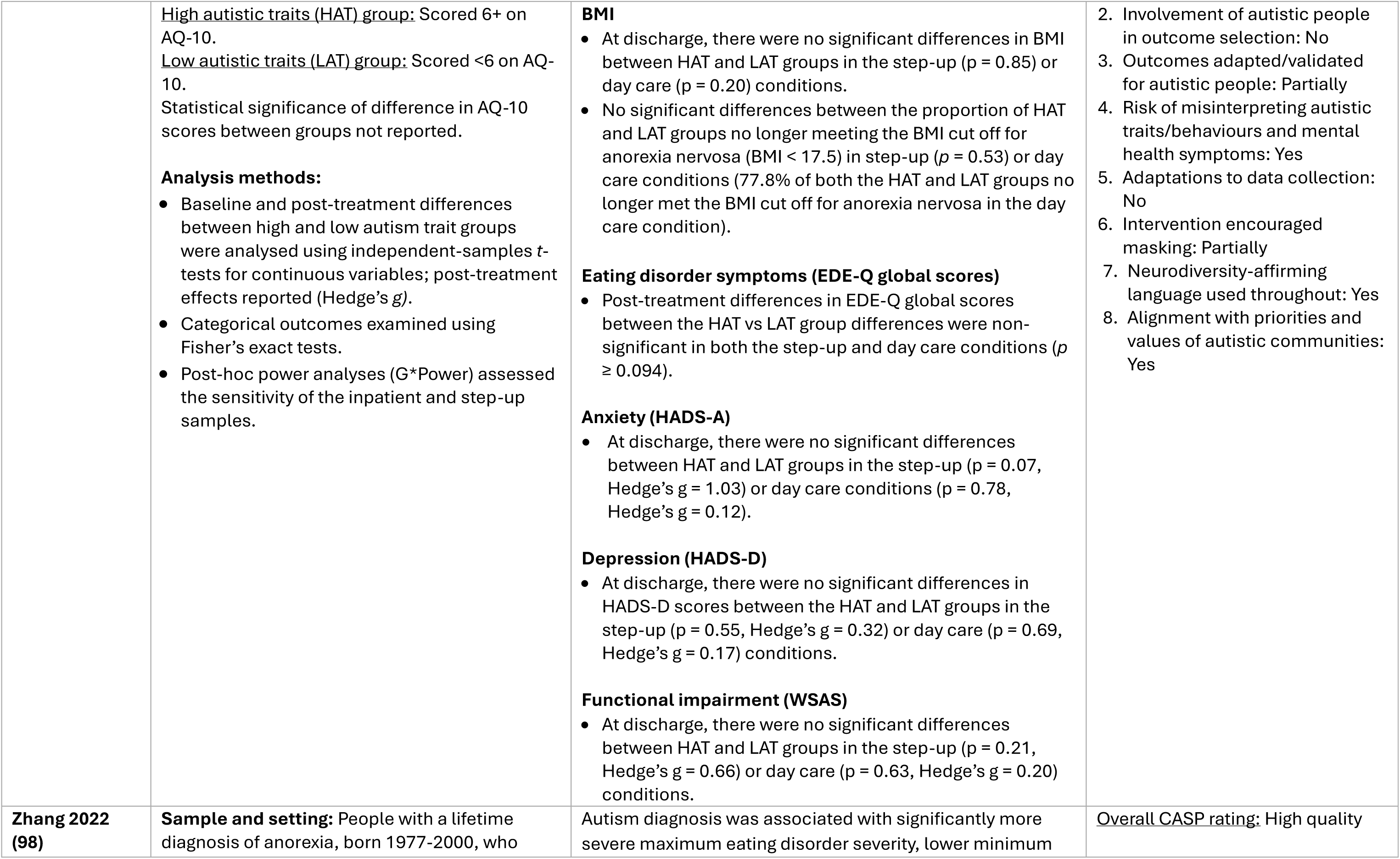
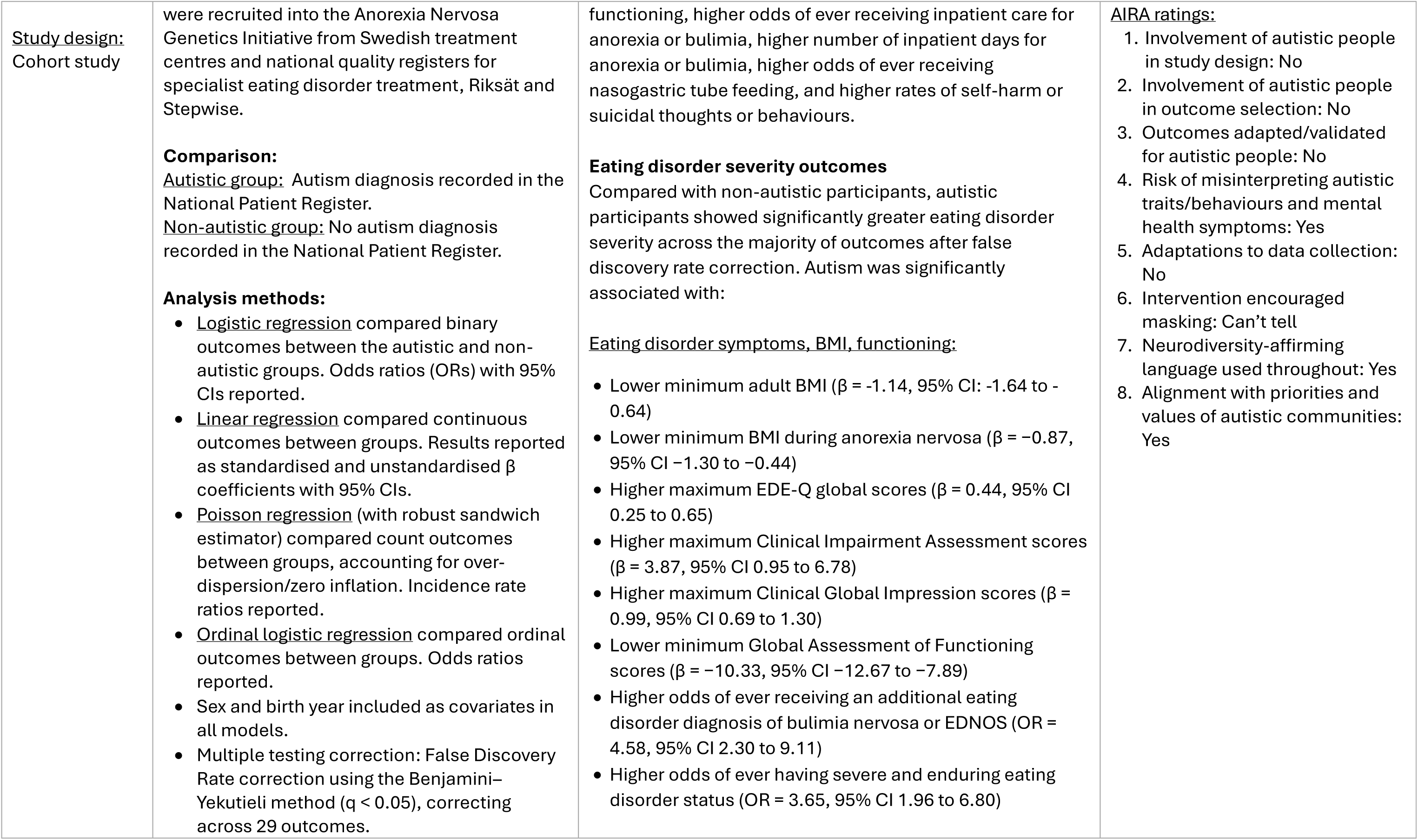
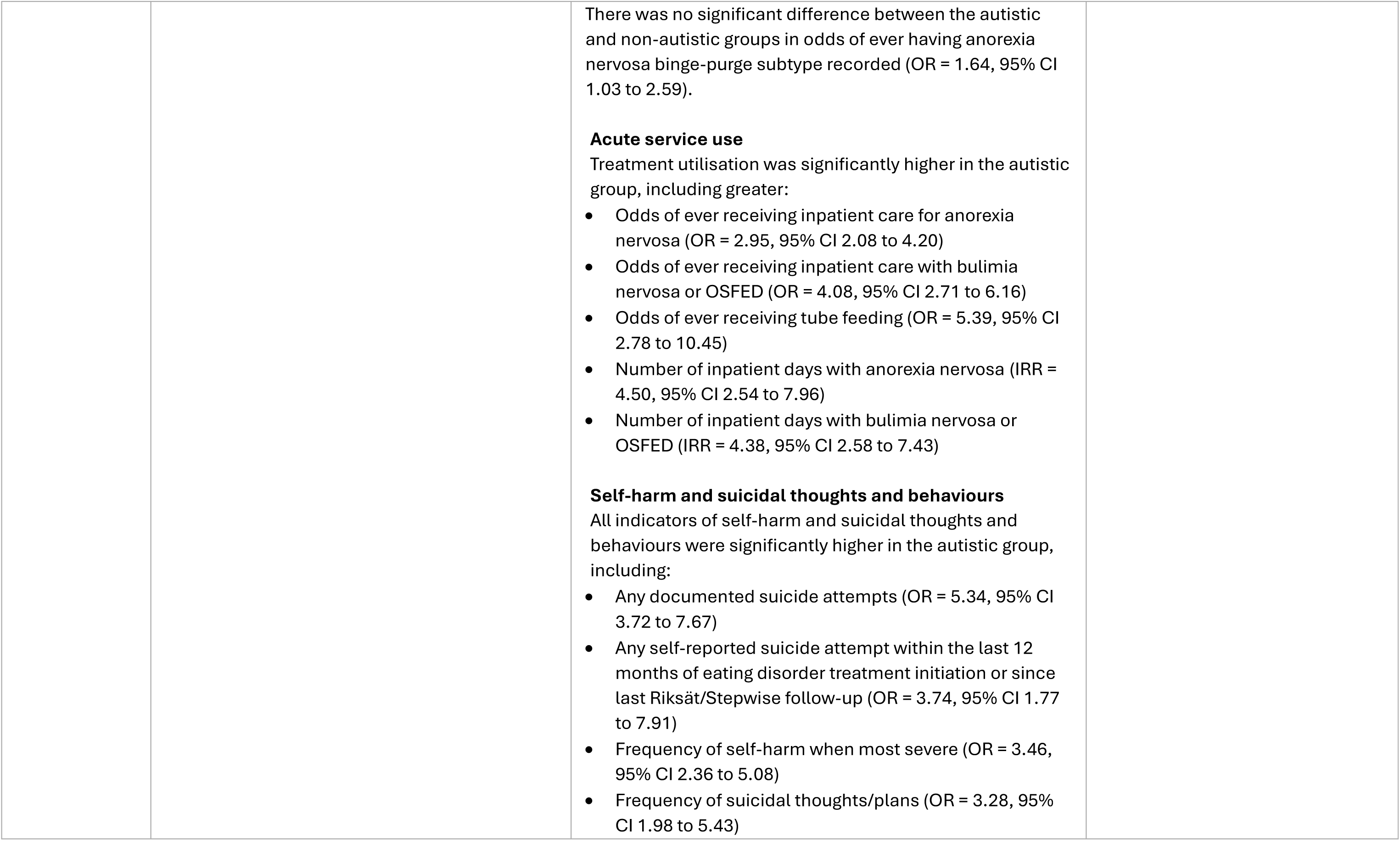
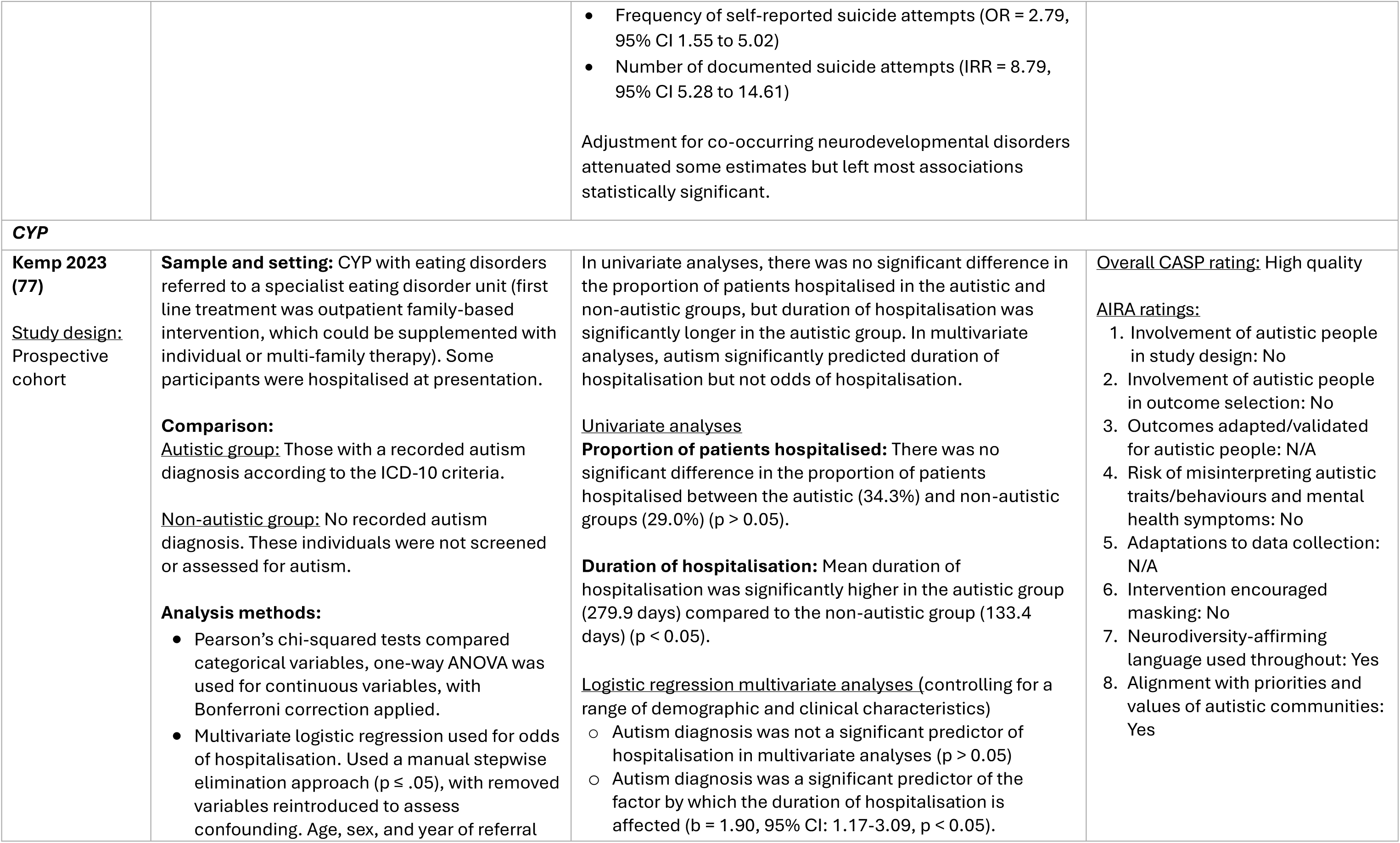
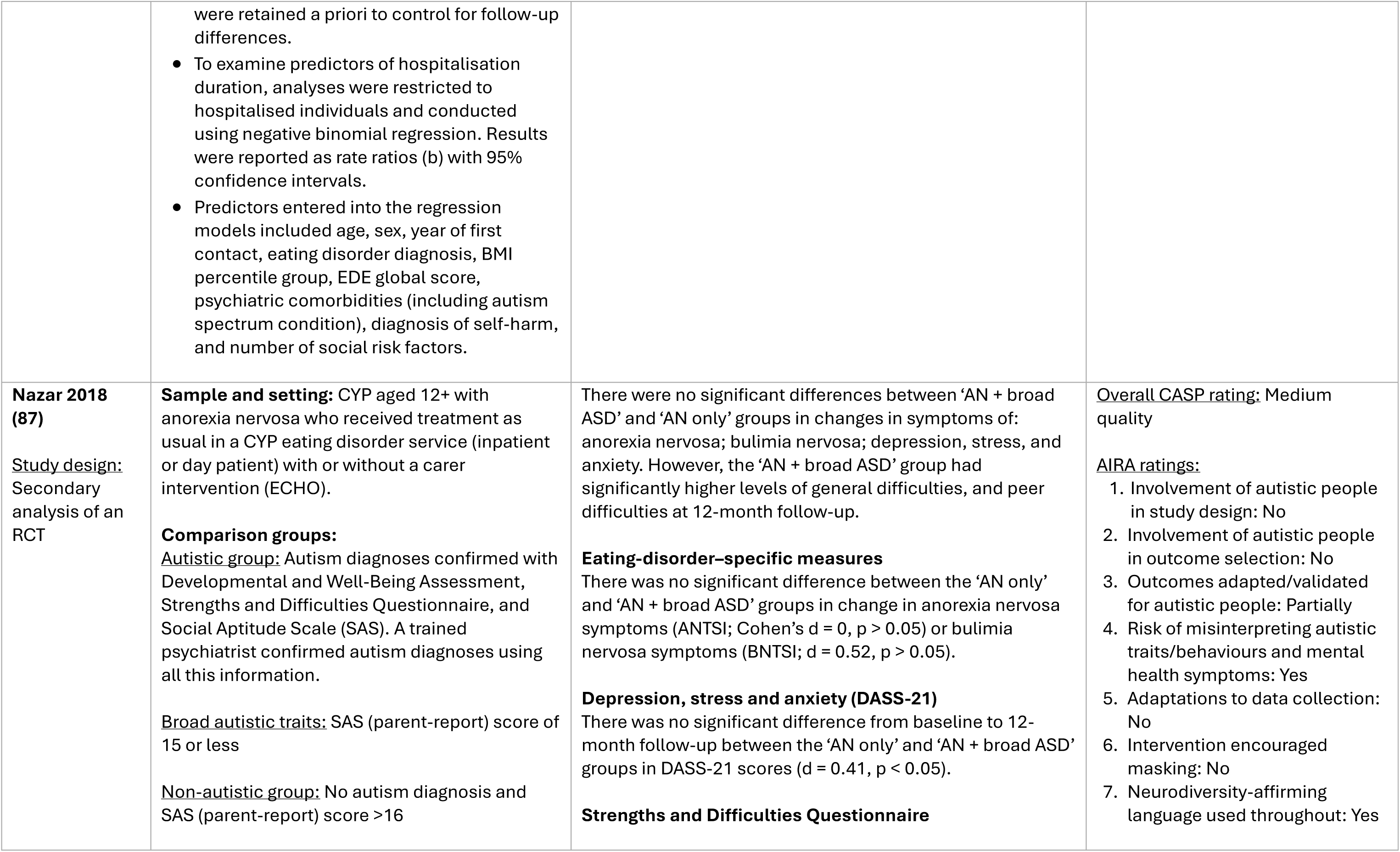
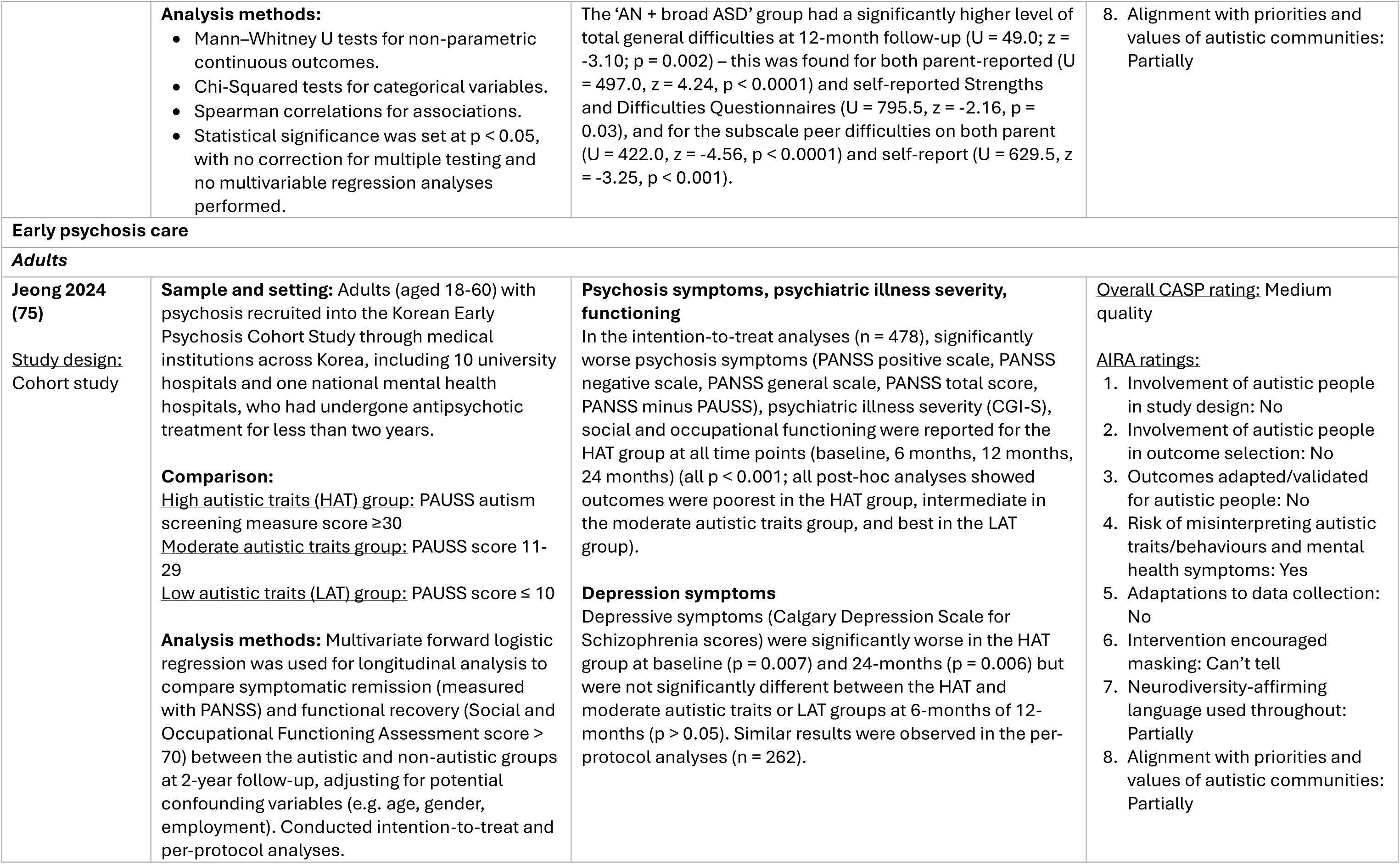
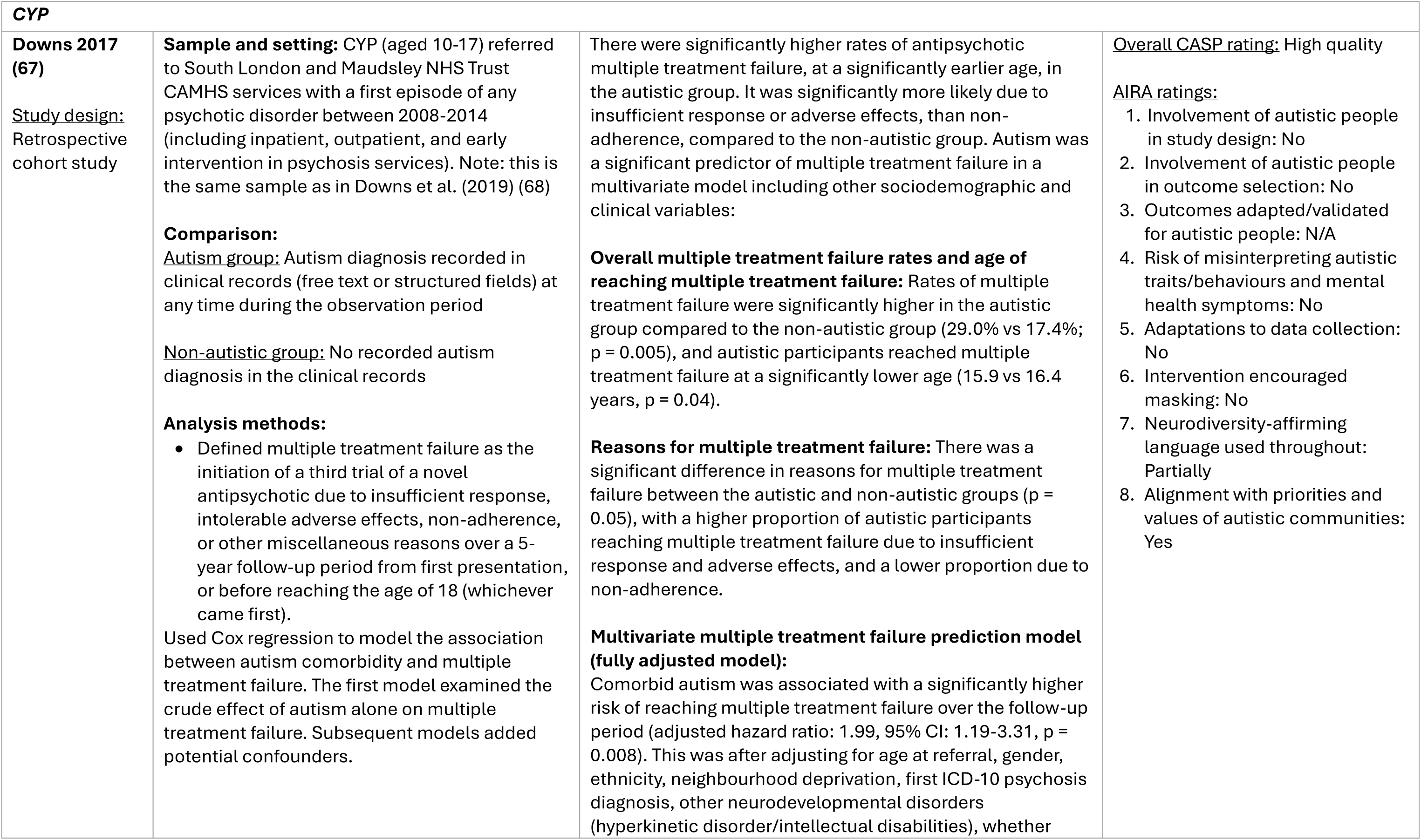
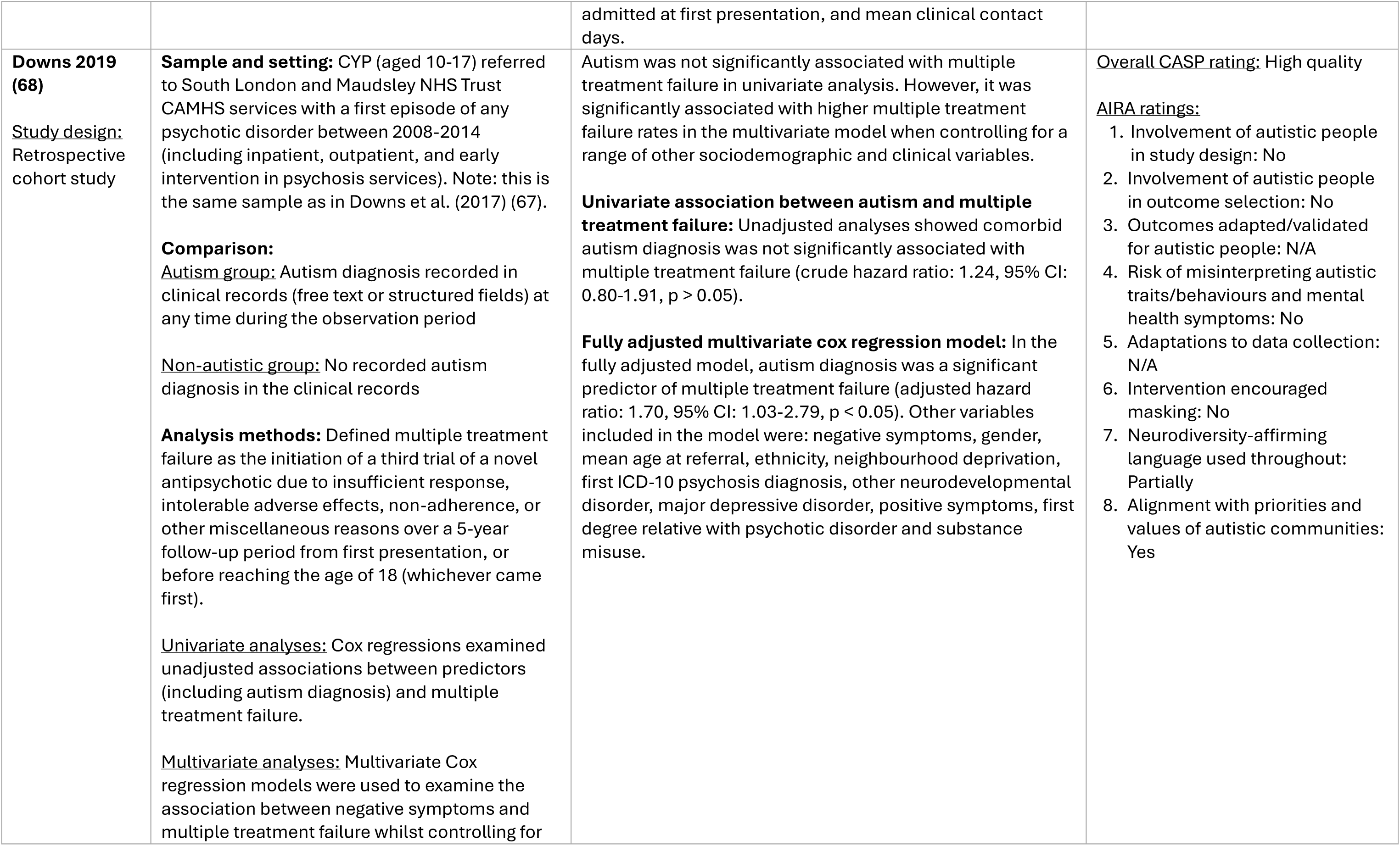
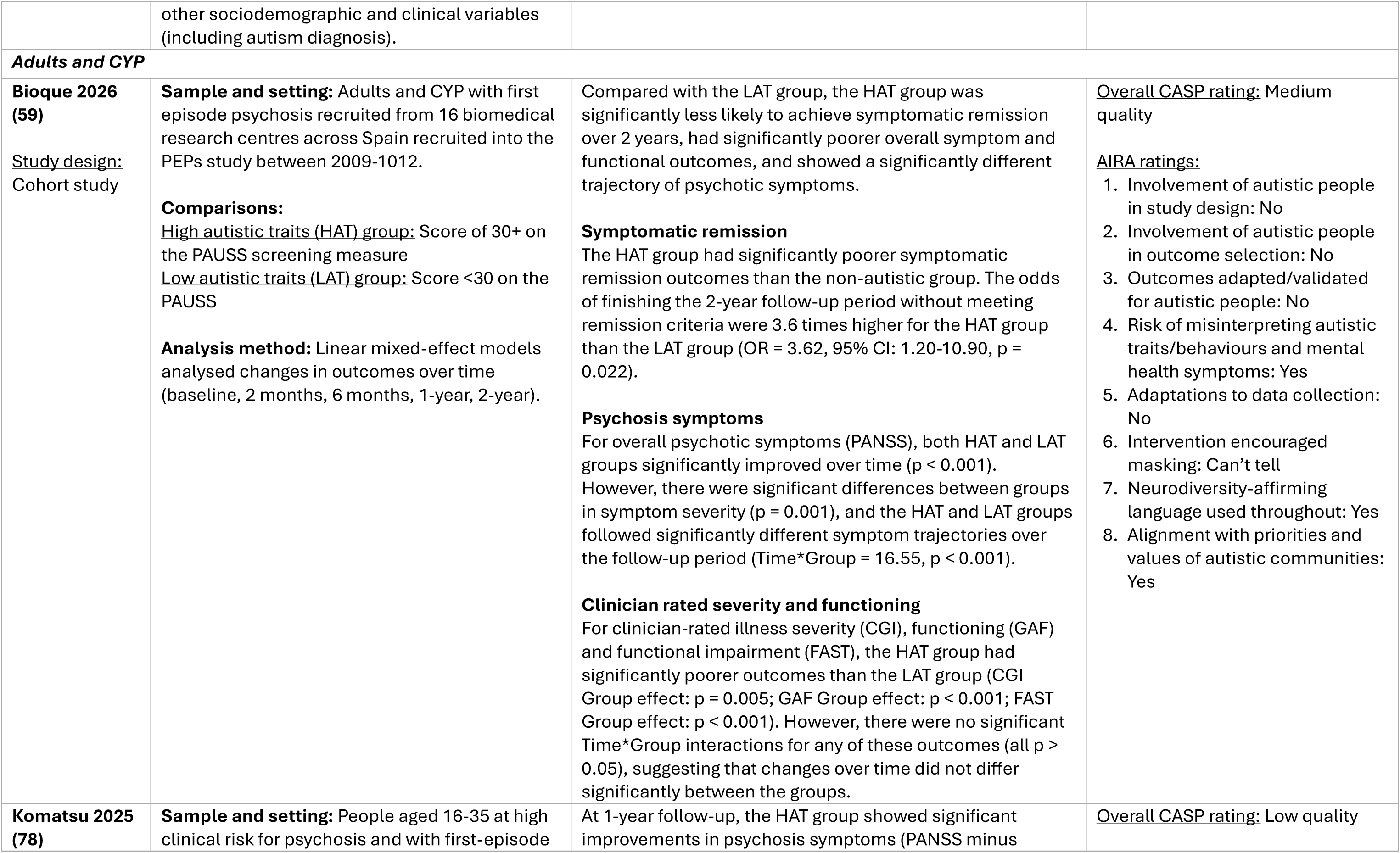
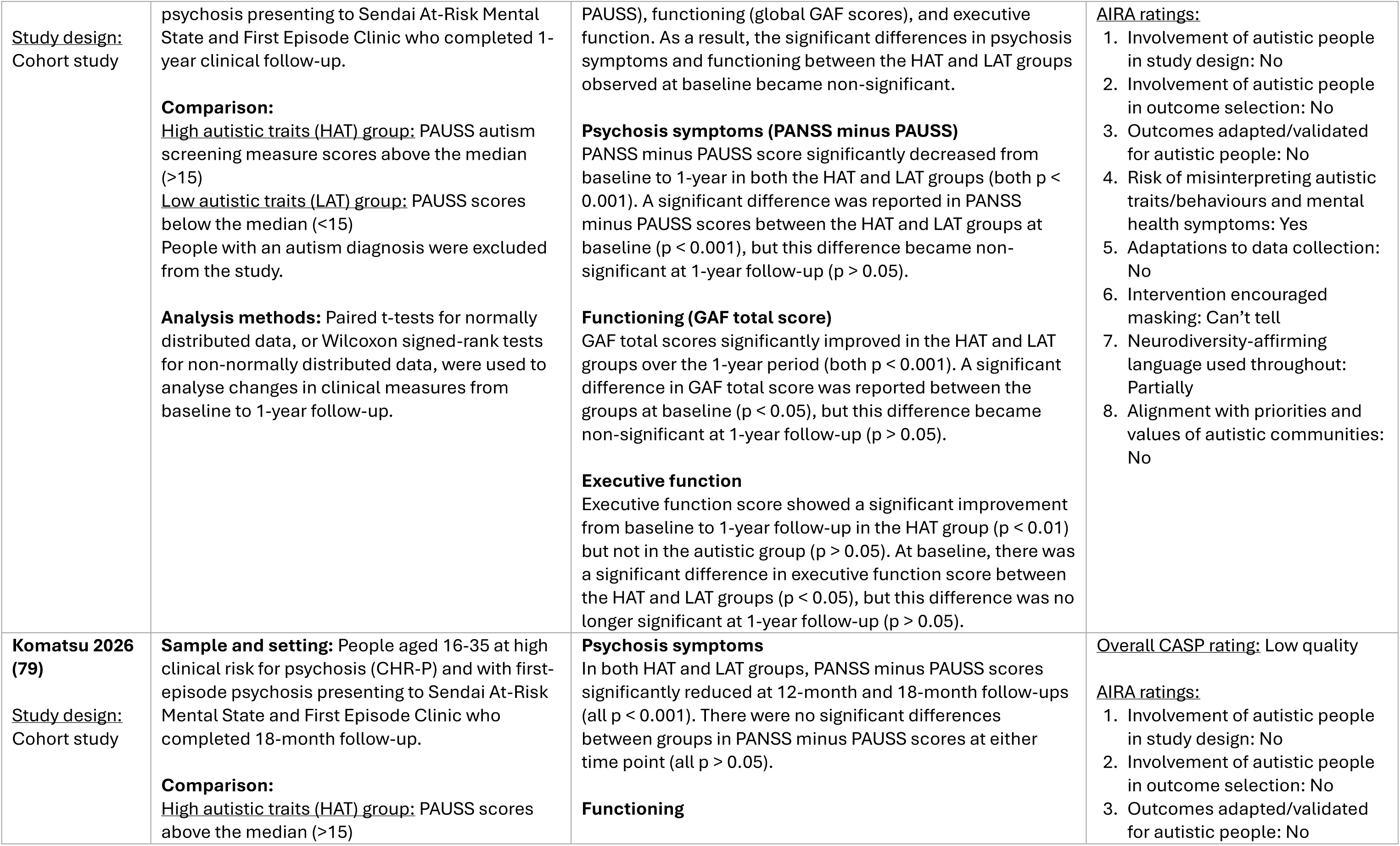
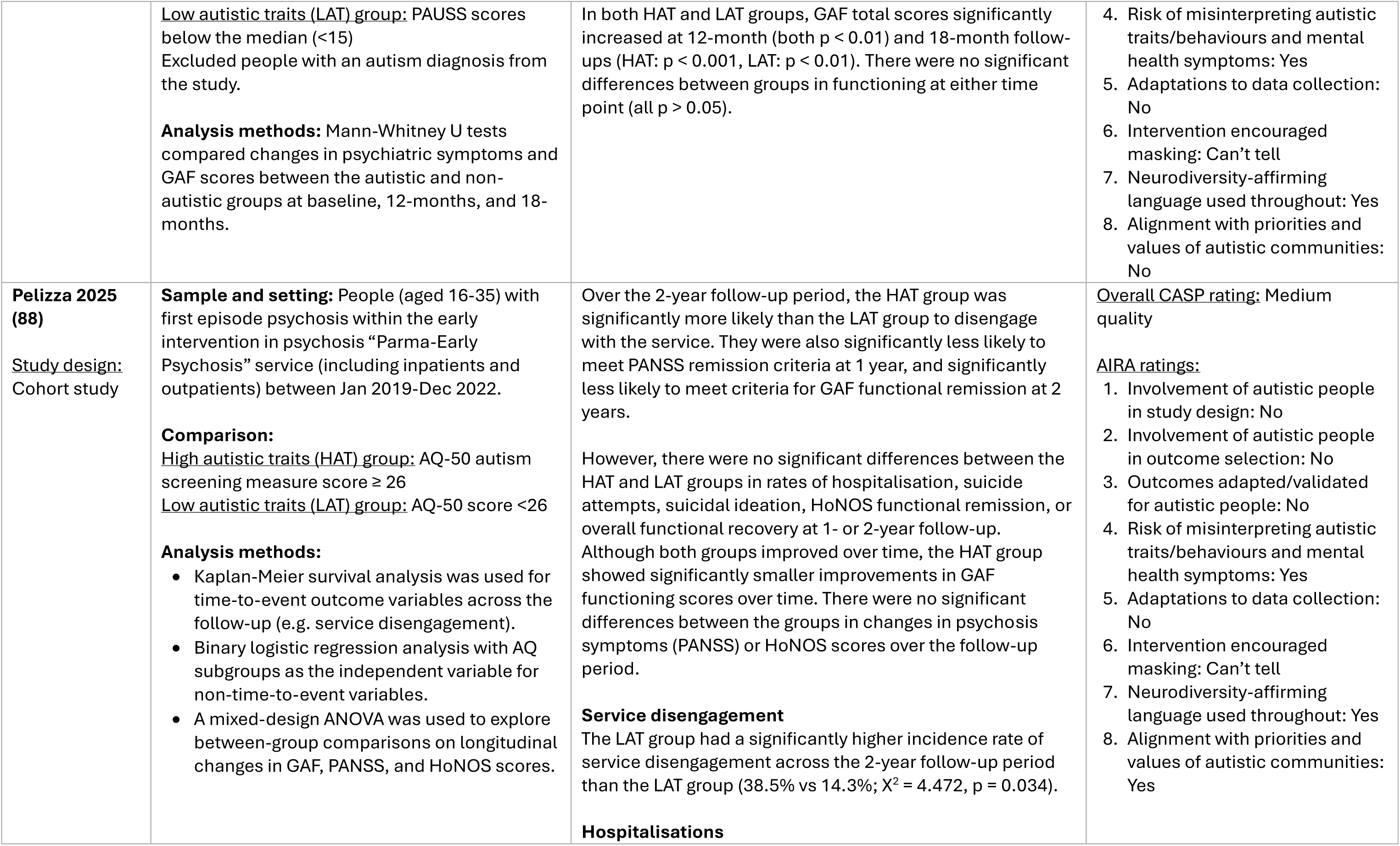
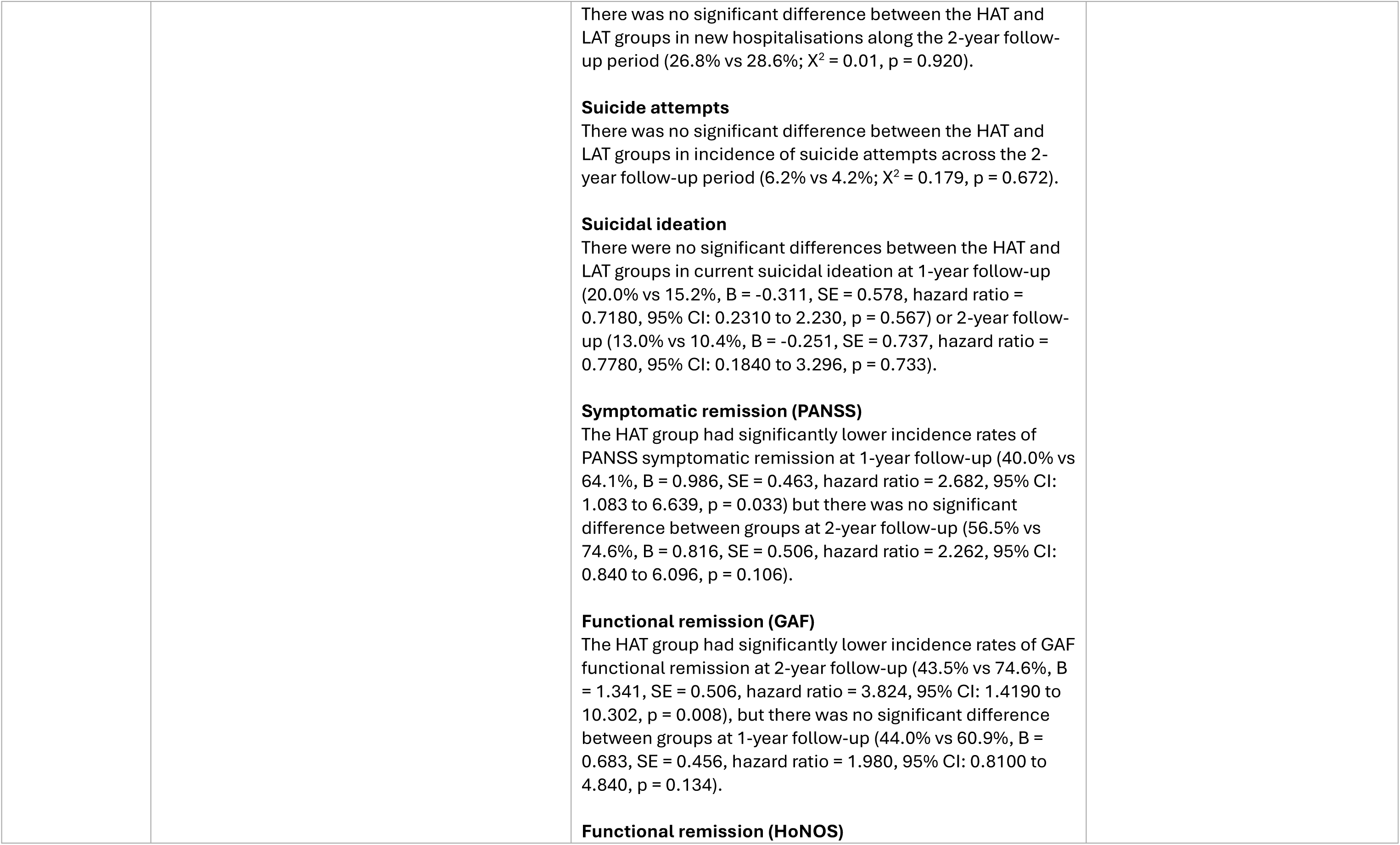
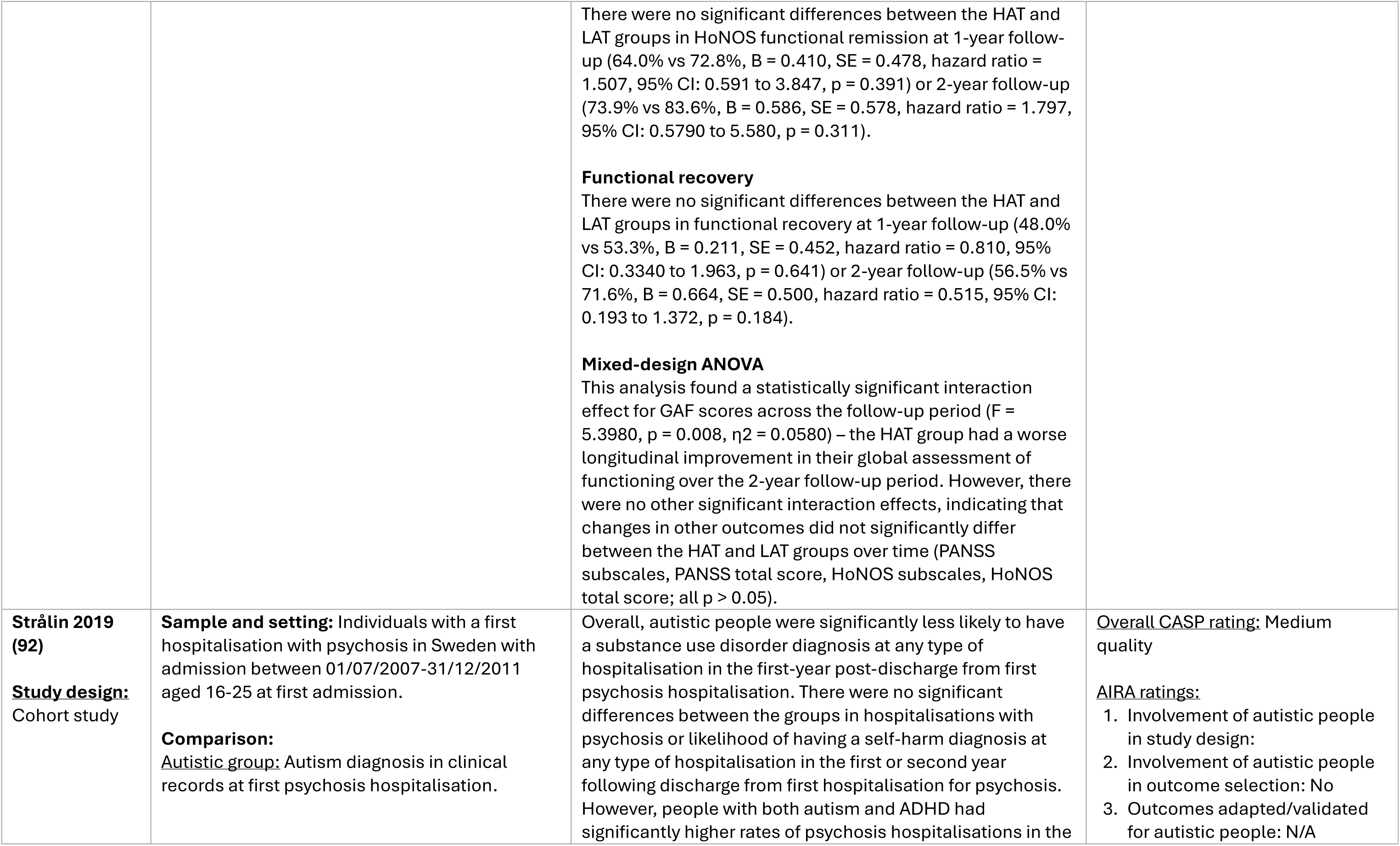
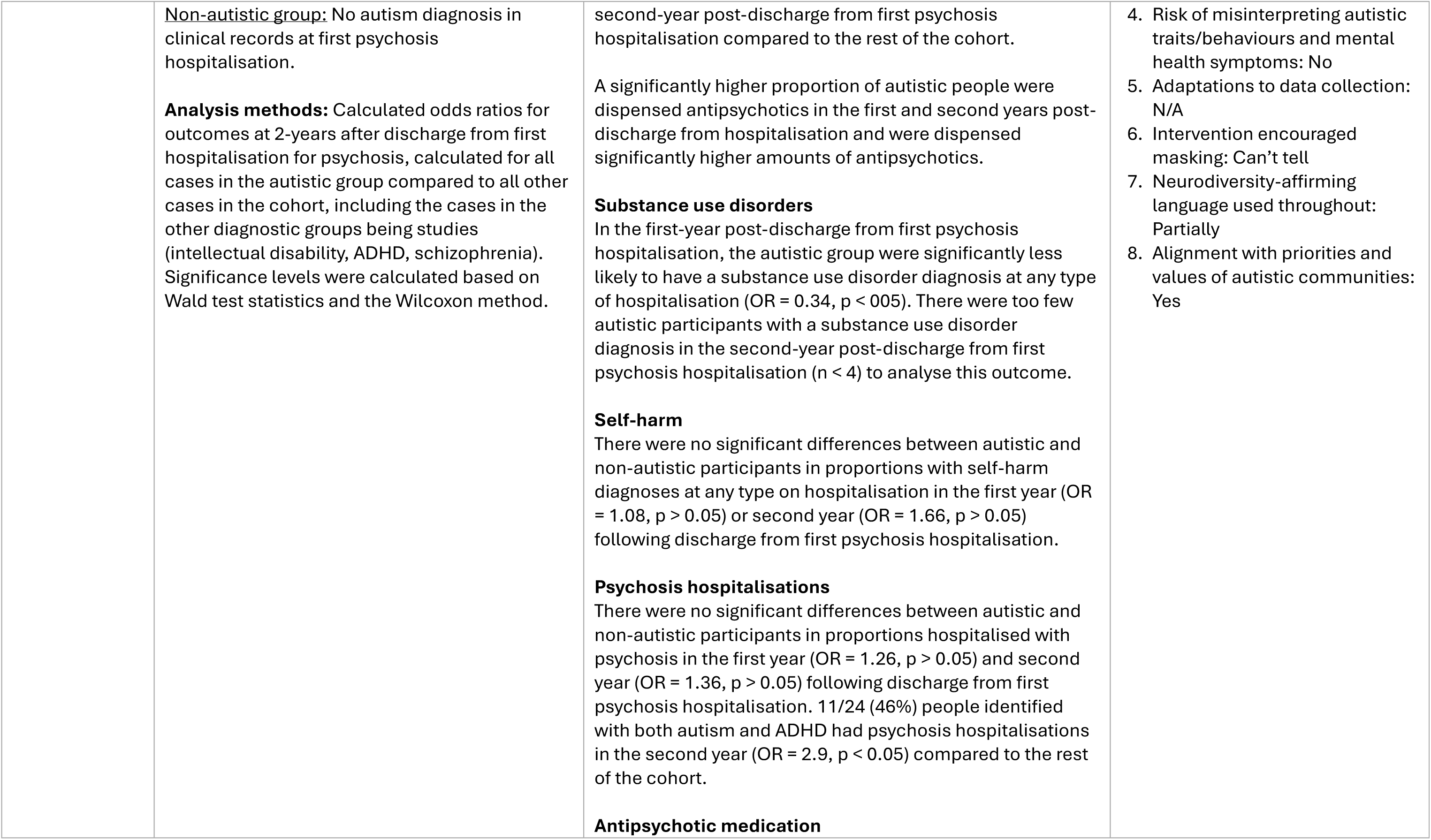
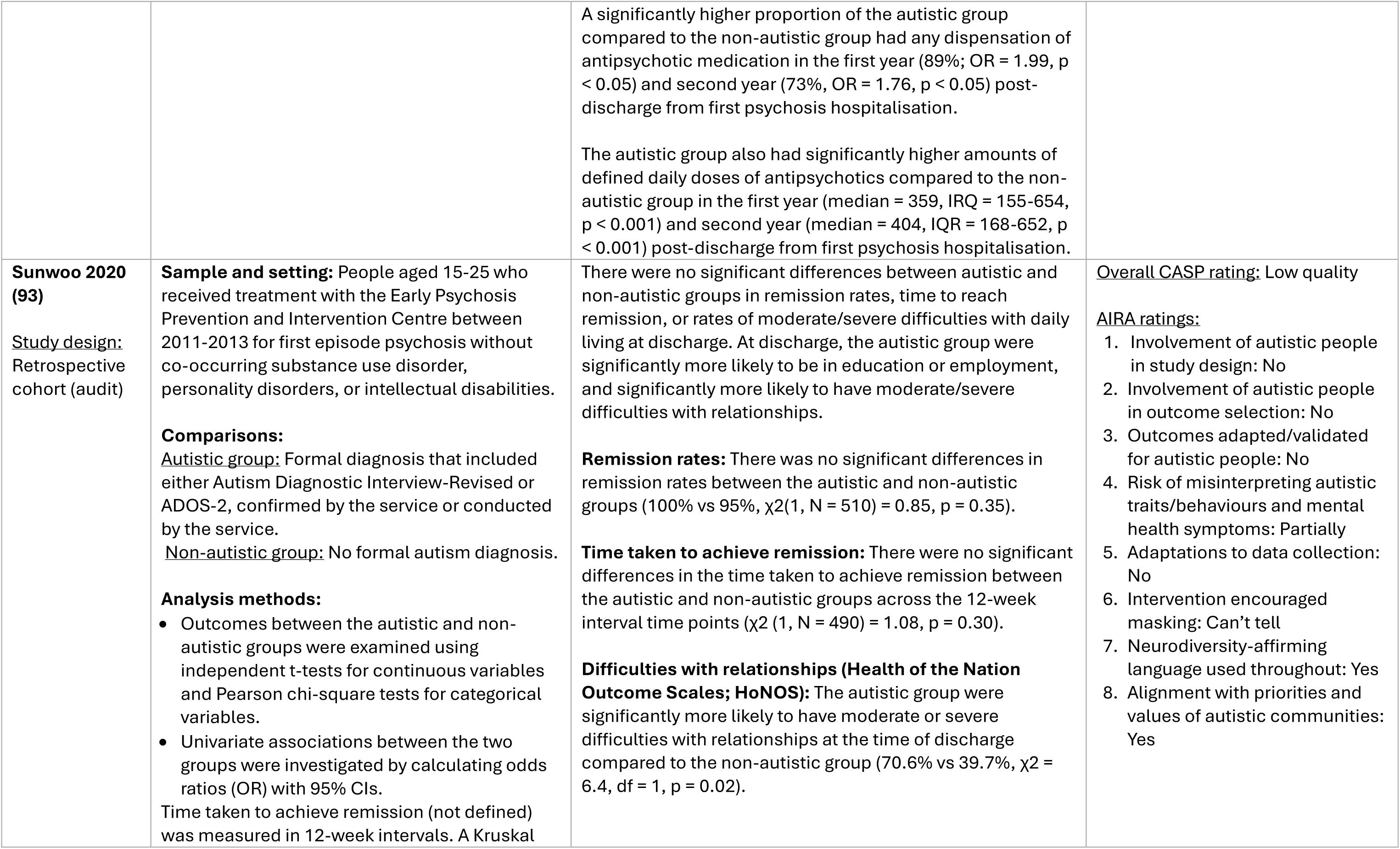
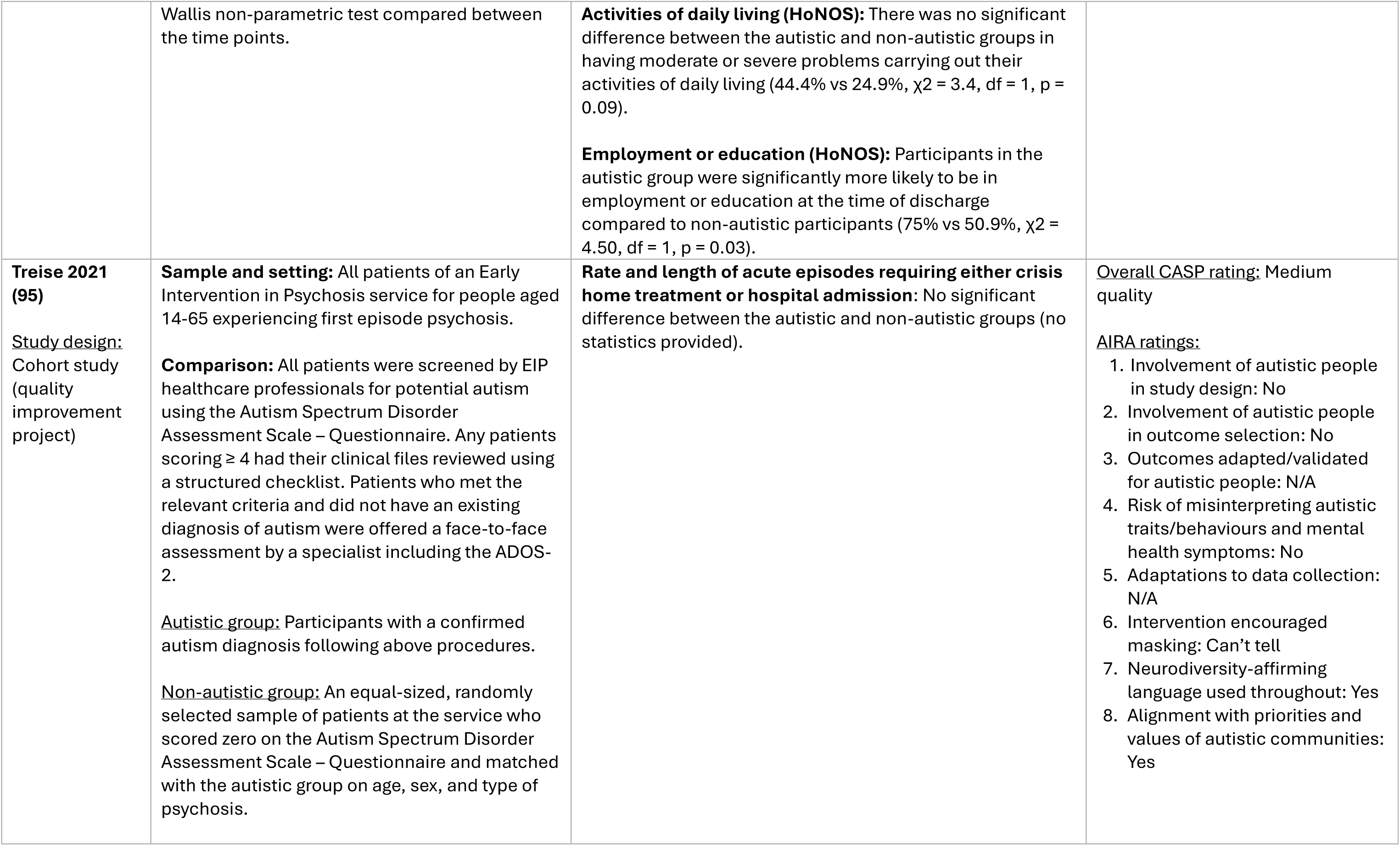
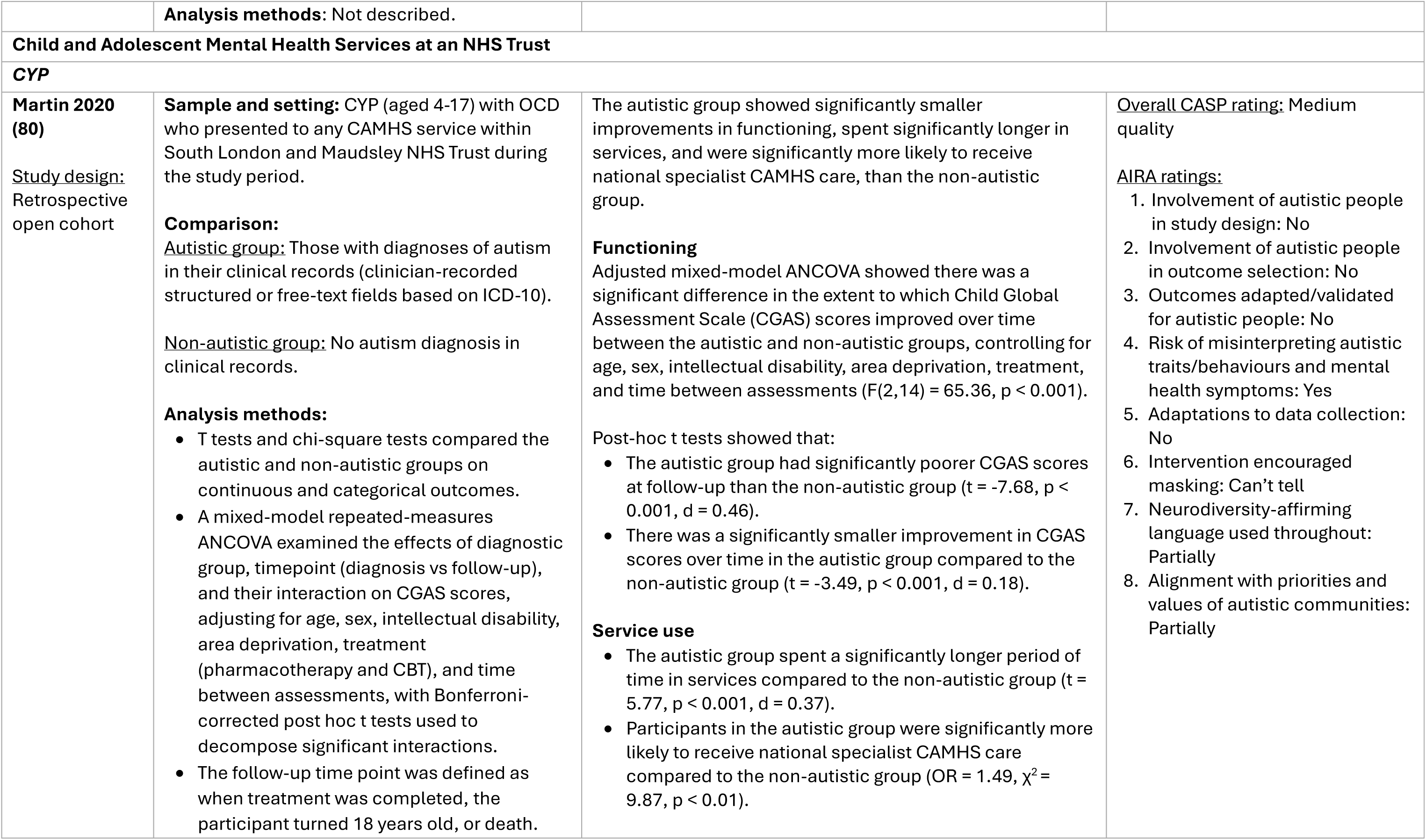
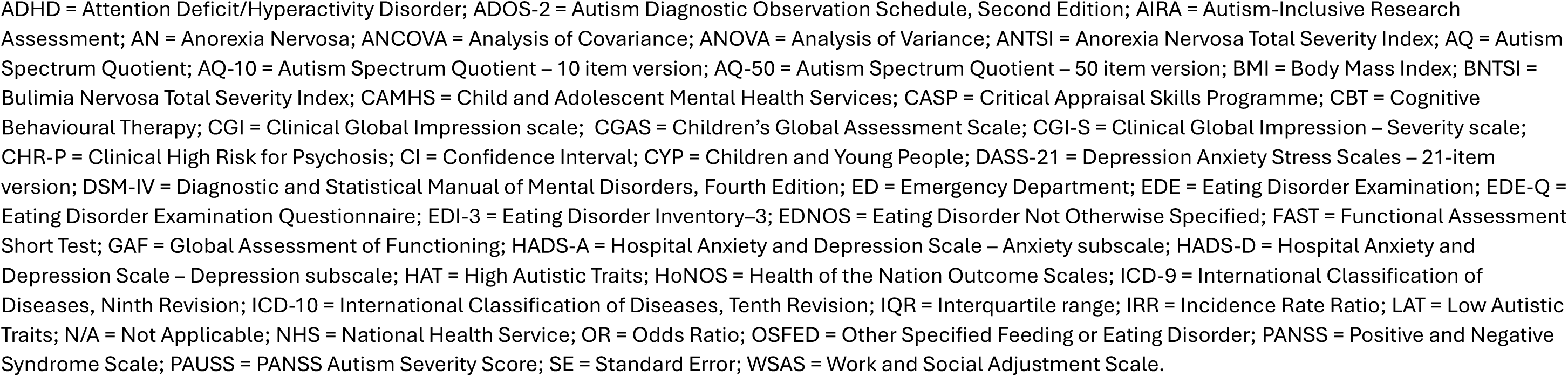
Quantitative evidence comparing mental health treatment outcomes for autistic and non-autistic people from multi-component/service-level approaches.

##### Emergency departments (EDs)

Two studies compared outcomes for people with and without autism diagnoses presenting to emergency departments for psychiatric reasons (58,76). Both were rated medium quality. One included CYP only (76) and one adults only (58).

Kalb et al.’s (2019) large study (autistic group n = 46343, overall n = 2784741) found autistic CYP reported significantly higher rates of ED visits for psychiatric crises compared with non-autistic people or those with other neurodevelopmental conditions, higher rates of hospitalisation, and increased likelihood of repeat presentations (76).

In a small sample (overall n = 86, autistic group n = 43), of adults with acute psychiatric presentations attending an emergency department and referred to an ‘Emergency Mental Health Team’, Arasu et al. (2023) reported no significant differences between autistic and non-autistic people in sedative medication use, except for significantly higher doses of benzodiazepines in the autistic group. There were no immediate severe adverse events reported following sedation in either group.

##### Forensic intellectual disabilities inpatient services

One medium-quality study (70), examined differences in restrictive practices and discharge outcomes among adults with and without an autism admitted to a forensic inpatient intellectual disabilities service. Autistic adults experienced significantly higher rates of physical restraint and observation than non-autistic adults, but there were no significant differences in rates of seclusion or pro re nata (PRN) medication use between groups. There was also no significant difference in length of stay, direction of the discharge pathway (discharge to the same or higher versus lower level of security) or in discharge placement locations across groups. Given the small sample size (overall n = 138, autistic group n = 42), these findings should be interpreted cautiously.

##### High-secure psychiatric hospital

One study (82), rated medium quality, compared outcomes for adults with and without an autism diagnosis in a high-secure psychiatric hospital. The autistic group size was small (n = 8), so results should be interpreted with caution. They reported that autistic adults were significantly over-represented on the hospital’s incompatibilities list (a formal record of a significant breakdown in dynamics between two or more patients resulting in risk of interpersonal conflict). The authors note that a formal incompatibility typically leads to patients being kept apart.

##### Specialist eating disorder inpatient units

Four studies evaluated specialist eating disorder inpatient units (54,73,90,94). Two were rated medium quality (54,94) and two low quality (73,90). Two studies compared HAT and LAT groups defined using autism screening measures (54,73). One study did not report how autistic and non-autistic people were categorised (94), and one used both diagnostic status and HAT/LAT groupings (90). Three studies recruited adults only (54,73,94), and one CYP only (90).

Li et al. (2020) included a moderately sized inpatient sample (HAT group n = 86; overall n = 306) (54). While post hoc power analysis indicated sufficient power to detect medium-to-large effects in BMI change, findings for other outcomes warrant cautious interpretation given substantial missing data. Baseline differences between groups were also not accounted for. They reported that at discharge, there were no significant differences between HAT and LAT adult inpatients in BMI and eating disorder symptoms, although significantly higher anxiety, depression, and functional impairment were reported by HAT inpatients.

Pruccoli et al. (2021) in a CYP sample reported no significant differences between autistic and non-autistic patients in changes in eating disorder symptoms, depressive or obsessive-compulsive symptoms, percentage Body Mass Index (BMI), length of inpatients stays, or rates of hospitalisations or paediatric emergency admissions (90). Huke et al. (2014) found no significant differences between HAT and LAT adults in rates of premature treatment termination (73). However, both studies had small sample sizes (overall n ≤ 32; HAT group n ≤ 8), so these results should be interpreted cautiously.

Tchanturia et al. (2021) compared inpatient length of stay before and after the implementation of the Pathway for Eating Disorders and Autism developed from Clinical Experience (PEACE pathway) between autistic and non-autistic people, but did not test the statistical significance of this difference (94).

Taken together, these findings suggest no significant differences between autistic and non-autistic CYP receiving specialist eating disorder inpatient care in changes in eating disorder symptoms, BMI, depression symptoms, obsessive-compulsive symptoms, or acute service use. However, despite these comparable changes in symptoms, adult HAT inpatients had significantly higher levels of anxiety, depression, and functional impairment at discharge, although there were no significant differences between HAT and LAT adult inpatients at discharge in eating disorder symptoms, BMI, or premature treatment termination rates. These findings should be interpreted cautiously due to small sample sizes.

##### Mixed specialist eating disorder settings (e.g. inpatient, step-up, day patient, outpatient)

Four studies examined treatment outcomes across mixed specialist eating disorder settings, including inpatient, day-patient, and outpatient services (54,77,87,98). Two studies were rated high quality (77,98), and two were rated medium quality (54,87). Two studies included adults only (54,98), and two included CYP only (77,87). Two studies compared people with and without autism diagnoses (77,98), whilst two compared HAT and LAT groups based on autism screening measure cut-offs (54,87).

Zhang et al. (2022) analysed data from a large national cohort of people with a lifetime diagnosis of anorexia nervosa, recruited between 2013 and 2016 through Swedish specialist eating disorder treatment centres and national quality registers for specialist eating disorder care (98). The sample included a substantial autistic subgroup (n = 134). However, analyses did not adjust for baseline differences in clinical severity or functioning so findings should be interpreted cautiously. Autistic adults demonstrated significantly greater eating disorder severity across most clinical outcomes, including lower minimum BMI, higher maximum eating disorder psychopathology, increased odds of severe and enduring eating disorder status, and poorer minimum functioning. Autistic participants also had significantly increased odds of inpatient admission, nasogastric tube feeding, and longer inpatient stays, as well as significantly elevated rates of self-harm and suicidal thoughts and behaviours. Most associations remained significant after adjustment for sex, birth year, and other comorbid neurodevelopmental disorders.

Kemp et al. (2023) examined outcomes for CYP referred to a specialist eating disorder unit providing inpatient and outpatient care (77). The sample was relatively large (autistic group n = 70; overall n = 552) (77). Autism diagnosis significantly predicted longer inpatient stays in both univariate and multivariate analyses controlling for demographic, diagnostic, and clinical factors, but was not significantly associated with likelihood of hospitalisation.

Nazar et al. (2018) (87) found no significant differences between HAT and LAT groups in changes in eating disorder symptoms, or depression, anxiety, and stress symptoms among CYP with anorexia nervosa accessing specialist eating disorder inpatient and day patient care. However, the HAT group had significantly greater general psychological difficulties, including peer relationship difficulties, at 12-month follow-up (not accounting for baseline differences). Li et al. (2020) found no significant differences between HAT and LAT groups in BMI, eating disorder symptoms, anxiety, depression, or functional impairment at discharge among adults with anorexia nervosa receiving step-up day patient or day patient care, but did not account for baseline differences (54). Findings from these studies should be interpreted cautiously due to their small sample sizes (autistic groups n ≤ 29).

Overall, evidence for greater illness severity after receiving mixed specialist eating disorder services and increased acute service utilisation among autistic individuals is strongest in large cohort studies such as Zhang et al. (2022) (98). In smaller clinical samples (54,77,87), findings suggest broadly similar mental health symptom improvement, longer average inpatient stays (but not significantly greater rates of admission) in the autistic group, and mixed findings regarding functional outcomes.

##### Early psychosis care

Ten studies examined outcomes from psychiatric care for early psychosis (59,67,68,75,78,79,88,92,93,95), including two studies analysing one cohort (67,68), and two studies both analysing another cohort (78,79). Two studies were rated high quality (67,68), five medium quality (59,75,88,92,95), and three low quality (78,79,93). One recruited adults only (75), two CYP only (67,68), and seven both adults and CYP (59,78,79,88,92,93,95). Five studies compared people with and without an autism diagnosis (67,68,92,93,95), four compared HAT and LAT groups based on autism screening measures (59,78,79,88), and one study compared high, medium, and low autistic traits groups based on autism screening measure scores (75).

Seven studies had small autistic group sample sizes (autistic group n ≤ 40), so their findings should be interpreted with caution (59,75,78,79,88,93,95). The remaining three studies had relatively large sample sizes, each with more than 100 autistic participants (67,68,92).

##### Antipsychotic treatment outcomes

Two studies of the same cohort, both rated high quality on the CASP, examined antipsychotic treatment outcomes among CYP referred to Child and Adolescent Mental Health Services (CAMHS) in a large urban NHS mental health trust with first episode psychosis (67,68). The sample sizes were relatively large (overall n = 638; autistic group n = 114). They found significantly higher rates of multiple treatment failure (defined as initiation of a third trial of a novel antipsychotic over a follow-up period of up to five years) for CYP with an autism diagnosis compared to those without, when controlling for a range of potential demographic and clinical confounders (67,68). Autistic participants reached multiple treatment failure at a significantly younger age (67). There were significant differences in reasons for multiple treatment failure, with autistic CYP more likely to require changes due to insufficient response and adverse effects, and a lower proportion due to non-adherence, compared to non-autistic CYP (67).

Strålin C Hetta (2019) found that following first psychosis hospitalisation, autistic people were significantly more likely to receive antipsychotic medication and had higher cumulative doses than non-autistic people during both the first and second years post-discharge.

##### Psychosis symptoms

Findings regarding psychosis symptom outcomes were mixed. Bioque et al. (2026) reported that, although psychosis symptoms improved significantly in both the HAT and LAT groups, the HAT group had significantly greater psychosis symptom severity throughout the two-year follow-up and followed a significantly different symptom trajectory (59). In contrast, Pelizza et al. (2025) found no significant difference between HAT and LAT groups in change in psychosis symptoms (88).

Jeong et al. (2024) found significantly greater psychosis symptom severity in the HAT group than the moderate and low autistic trait groups at all follow-up assessments over two years, but this did not account for baseline differences (75). In contrast, Komatsu et al. (2025, 2026) found that, although the HAT group had significantly higher psychosis symptom severity at baseline, there were no significant between-group differences at 12-or 18-month follow-up (78,79).

##### Symptomatic remission

Findings on symptomatic remission were also mixed. Bioque et al. (2026) found that the HAT group was significantly less likely to achieve symptomatic remission than the LAT group (59). Similarly, Pelizza et al. (2025) found significantly lower rates of symptomatic remission in the HAT group at one-year follow-up, although this difference was no longer significant at two years (88). In contrast, Sunwoo et al. (2020) observed no significant differences in psychosis remission rates, or time to remission, between autistic and non-autistic participants (93).

##### Other mental health outcomes

Findings regarding other psychiatric outcomes were mixed. Strålin & Hetta (2019) reported that autistic people were significantly less likely than non-autistic people to have a substance use disorder diagnosis during any hospitalisation in the first year following discharge from their first hospitalisation with psychosis (92). Jeong et al. (2024) reported greater depression symptom severity in the HAT group compared with the moderate and low autistic trait groups at some, but not all, time points over the two-year follow-up, and found no significant between-group differences in overall psychiatric illness severity at any time point, but did not account for baseline differences (75)

##### Suicide and self-harm

Strålin & Hetta’s (2019) national cohort study reported no significant differences between autistic and non-autistic people in rates of self-harm diagnoses at hospitalisations during the first or second year following discharge from their first psychosis hospitalisation (92). Similarly, Pelizza et al. (2025) found no significant differences between HAT and LAT groups in suicidal ideation or suicide attempts over a two-year follow-up (88).

##### Functional outcomes

Findings regarding functional outcomes were mixed. Among studies examining changes in functioning over time, Bioque et al. (2026) found no significant differences between HAT and LAT groups in changes in functioning or functional impairment over time (59). Pelizza et al. (2025) reported mixed findings: the HAT group showed smaller improvements and lower remission rates on the Global Assessment of Functioning (GAF), but no significant differences in Health of the Nation Outcome Scales (HoNOS) functioning scores or remission rates (88).

In studies comparing groups at individual assessment time points, Komatsu et al. (2025, 2026) found significantly poorer global functioning and executive functioning in the HAT group at baseline, but no significant between-group differences in executive functioning at 12-month follow-up or in global functioning at 12-or 18-month follow-up (78,79). Jeong et al. (2024) reported significantly poorer social and occupational functioning in the HAT group compared to moderate and low autistic traits groups at all follow-up assessments over two years (75). Sunwoo et al. (2020) found no significant differences between autistic and non-autistic participants in difficulties with daily activities at discharge from an Early Intervention in Psychosis (EIP) service, but autistic participants were significantly more likely to have relationship difficulties, and to be in employment or education at discharge (93). However, baseline differences were not taken into account.

##### Acute service use

Strålin & Hetta (2019) found no significant differences between autistic and non-autistic people in psychosis-related hospitalisation rates during the first or second year following discharge after a first hospitalisation for psychosis (92). However, people with co-occurring autism and Attention-Deficit/Hyperactivity Disorder (ADHD) had significantly higher rates of psychosis-related hospitalisation during the second-year post-discharge compared to the rest of the cohort. Similarly, Pelizza et al. (2025) found no significant differences in hospitalisation rates between HAT and LAT groups receiving care from an Early Intervention in Psychosis (EIP) service, although the HAT group had significantly higher rates of disengagement from the EIP service (88). Likewise, Treise et al. (2021) found no significant differences between HAT and LAT groups in the rate or duration of acute episodes requiring crisis home treatment or hospital admission among people attending an EIP service (95).

##### Summary

Overall, evidence regarding outcomes for early psychosis care for autistic compared with non-autistic people was mixed. There was evidence that autistic people were significantly more likely to receive antipsychotic medication, and higher cumulative doses of antipsychotics, following first psychosis hospitalisation, and that autistic CYP are significantly more likely to experience antipsychotic multiple treatment failure. Across studies, there were generally no significant between-group differences in self-harm, suicidal ideation, suicide attempts, crisis home treatment use or hospitalisations, although people with co-occurring autism and ADHD had significantly higher rates of psychosis-related hospitalisation following discharge from a first psychosis admission. This study also found that autistic people were significantly less likely to have a substance use disorder diagnosis during subsequent hospitalisations. Findings regarding psychosis symptoms, symptomatic remission, depression symptoms, and functional outcomes were mixed, with studies reporting either no significant differences or poorer outcomes among autistic groups. Given most studies’ small sample sizes, these findings should be interpreted with caution.

##### Child and Adolescent Mental Health Services at an NHS Trust

One study (80), rated medium quality, compared outcomes for CYP with OCD with and without an autism diagnosis presenting to CAMHS services within a large NHS Trust in England. The study included a relatively large autistic subgroup (n = 335). The study reported a significantly smaller improvement in a broad measure of functioning from baseline to follow-up in the autistic group compared to the non-autistic group, even when adjusting for a range of demographic and clinical factors that could influence outcomes. The autistic group also spent significantly longer in services and were significantly more likely to receive national specialist CAMHS care.

## 4. Discussion

### 4.1. Key findings

To our knowledge, this is the first systematic review to compare mental health treatment outcomes for autistic and non-autistic people across a broad range of mental health interventions and services, settings, populations, and outcomes. The review identified 40 relevant papers, nearly half (46.2%) of which reported on UK studies. Overall, the findings suggest that mental health treatment outcomes for autistic compared with non-autistic people vary across interventions and outcomes. Although many studies reported no significant between-group differences, where significant differences were observed they were usually in the direction of poorer outcomes for autistic people, with little evidence of better outcomes. Of the 36 studies conducting statistical comparisons, 28 reported at least one outcome with no significant difference between groups, 25 reported at least one significantly poorer outcome for autistic people, and five reported at least one significantly better outcome for autistic people (categories were not mutually exclusive). Importantly, poorer outcomes typically reflected smaller improvements in the autistic group rather than a complete lack of benefit. However, confidence in these findings is limited by variable study quality, small sample sizes, and limitations in the inclusiveness of research for autistic people.

Overall, evidence generally suggested either comparable or smaller improvements in mental health symptoms for autistic compared with non-autistic groups, although the pattern varied across interventions and services. Comparable outcomes were observed in some larger, medium-and high-quality studies, including for improvements in psychological functioning following university counselling psychotherapy (57) and hospitalisations with self-harm diagnoses following first psychosis hospitalisation (92). Evidence for CBT for anxiety also suggested generally comparable outcomes, although some differences emerged depending on the anxiety measure, follow-up time point, and informant (60,97). Most other studies reporting comparable symptom outcomes were small and did not report power calculations, limiting confidence in these findings. Other large, medium-and high-quality studies found poorer outcomes for autistic people, including smaller improvements in anxiety and depression for autistic adults in NHS Talking Therapies services (69), higher peak eating disorder symptom severity and rates of suicidal thoughts and behaviours for autistic people with anorexia nervosa followed-up from mixed specialist eating disorder services (98), and higher post-treatment OCD symptom severity following CBT for OCD for autistic adults in our meta-analysis (81,84,96).

Overall, this suggests that although symptom improvement occurs for many autistic people, it may be more limited in magnitude than for non-autistic people in some settings. Findings regarding functioning broadly reflected the pattern observed for mental health symptoms, with autistic compared with non-autistic people showing either comparable outcomes or poorer functional outcomes across interventions and services. A large, high-quality study of CBT for anxiety found no significant difference between autistic and non-autistic CYP in changes in family functioning (72), and a large, medium-quality study found no significant differences in functional impairment at discharge from specialist day care or step-up day care eating disorder services (Li et al., 2020). Most other studies reporting comparable functional outcomes were small and did not report power calculations, limiting confidence in these findings. In contrast, other large, medium-and high-quality studies reported poorer functional outcomes for autistic adults accessing NHS Talking Therapies services (69), CYP with OCD receiving CAMHS services (80), and people with anorexia nervosa accessing inpatient and mixed specialist eating disorder services (54,98). For some interventions, findings varied across studies or according to the functioning measure or follow-up time-point assessed (e.g., Sunwoo et al., 2020 (93); Pelizza et al., 2025 (88)). This highlights the importance of assessing multiple domains of functioning rather than assuming uniform effects.

These findings should be interpreted in the context that most interventions targeted mental health symptoms rather than broader functional needs. Although improvements in mental health symptoms may contribute to improved functioning, they may not address pre-existing or ongoing challenges that autistic people may experience more frequently due to unmet support needs, environmental barriers, or a lack of appropriate accommodations. In addition, functioning measures often assess domains such as social participation, education, employment, and independence, and may reflect normative expectations of functioning that do not necessarily capture what is meaningful to autistic people. This highlights the need for more holistic interventions that address wider challenges experienced by autistic people, guided by their individual goals, preferences, and desired ways of living.

In relation to psychosis, large high quality studies analysing the same cohort found that autistic CYP with first episode psychosis experienced higher rates of multiple antipsychotic treatment failure, occurring at a younger age, even after adjustment for demographic and clinical factors (67,68). Treatment failure was more often due to insufficient response or intolerable adverse effects rather than non-adherence, compared with non-autistic groups. Similarly, Strålin C Hetta’s (2019) medium quality Swedish national cohort study found that a significantly higher proportion of autistic people were dispensed antipsychotic medication and received significantly higher total antipsychotic doses than non-autistic people in the two years following discharge from first psychosis hospitalisation (92). Together, these findings suggest differences in antipsychotic treatment and response between autistic and non-autistic people with psychosis. The reasons for these differences remain unclear but may reflect differences in neurobiology, clinical complexity, co-occurring conditions, and difficulties distinguishing autistic traits, distress, and psychotic symptoms, which could impact assessment and treatment selection (1,67,103). In addition, prescribing may be influenced by clinician perceptions of autistic people’s clinical needs and risk, as well as the availability of treatment approaches perceived as suitable for autistic people with psychosis. Qualitative research exploring both autistic people’s experiences of antipsychotic treatment alongside clinicians’ decision-making processes could help disentangle these factors.

There was also some evidence from large, medium-and high-quality studies of higher acute service use among autistic individuals, including increased repeat emergency department presentations and higher hospitalisation rates following emergency department visits (76), greater likelihood of accessing national specialist CAMHS for autistic CYP with OCD (80), and longer inpatient stays for autistic people accessing mixed specialist eating disorder services (77,98). There was also evidence of higher rates of psychosis-related hospitalisations in the two years post-discharge from first psychosis hospitalisation for autistic people with co-occurring ADHD (92). These findings are consistent with broader evidence of high and increasing rates of mental health service use by autistic people (9,104). For example, national statistics indicate that admissions of autistic people to mental health hospitals in England have increased over time, and that in May 2026 over 40% of autistic inpatients had been in hospital for two years or more (16). Other studies in this review reported no significant differences in acute service use, or mixed findings, but had small samples, limiting confidence in these results.

Some studies additionally reported higher rates of restrictive practices, such as nasogastric tube feeding for autistic people with anorexia nervosa (98). This is consistent with national data in England indicating that autistic people with eating disorders in inpatient settings are more likely to be nasogastric tube fed (17). Other examples included higher rates of physical restraint and observation in a forensic intellectual disabilities inpatient service (70). These patterns align with broader evidence that autistic people experience high rates of restrictive practices (e.g., physical restraint, chemical restraint, seclusion) (15,16,105–107), which can undermine therapeutic relationships and cause physical and psychological harm (108,109), highlighting this as an area of concern in current service provision.

Observed differences in outcomes may reflect interacting systemic, clinical, and individual-level factors. One potential explanation is baseline differences in clinical presentation, as autistic people may present to services with greater support needs or difficulties in day-to-day functioning, higher rates of suicidal thoughts and behaviours, more unmet support needs, or more complex or long-standing mental health difficulties (1,4,69,110–112). These factors are associated with smaller and slower improvements and higher rates of deterioration (111,113,114). However, differences in outcomes persisted in some studies even after controlling or accounting for baseline demographic and clinical characteristics (e.g. baseline mental health symptoms, diagnoses, intellectual disability, psychotropic medication) (60,67–69,77,96), suggesting additional influences.

Differences in outcomes may also reflect previously described barriers to accessing timely, effective, and appropriately adapted care, including mental health services and interventions that have not always been designed with autistic people in mind and may not adequately accommodate their needs, alongside variation in autism-specific expertise and supports (19,31,115–118). Delayed access to effective support may contribute to more severe difficulties at first presentation, longer or more intensive treatment, and higher use of acute services and more restrictive practices.

Autistic people may also experience and communicate distress differently, for example, through changes in repetitive behaviours, heightened sensory sensitivities, or shutdowns. Their distress may also be shaped by different drivers, such as sensory overload, social exclusion, masking, and chronic minority stress arising from environments not designed for autistic people (119–125). Further research is needed to understand how these factors may influence assessment, treatment response, and outcomes.

Limitations in outcome measurement may compound these challenges. Most studies that included patient-reported outcomes used outcome measures not developed or validated for autistic populations, risked misinterpreting autistic traits/behaviours and conflating or mistaking these for mental health symptoms, and did not adapt data collection tools or procedures to facilitate wide participation (e.g. providing accessible information, flexibility in communication modes, or more detailed response options than standard Likert scales, which may be experienced as ambiguous by some autistic people). Even where validation evidence exists, it remains limited, and not necessarily widely generalisable. This raises concerns about whether standard outcome measures are accessible or capture meaningful change for autistic people, particularly for under-represented groups (e.g., people with intellectual disabilities, individuals with minimal verbal communication, ethnic minority groups) (126,127).

Most studies in this review reported on mental health interventions or services without reported adaptations for autistic people. Where adaptations were described, these ranged from service-level models (e.g., PEACE pathway (94)) to simpler adjustments to intervention delivery (e.g., use of clear language, visual aids, and increased caregiver involvement in CBT for OCD (84)). Reporting of adaptations was often insufficient to determine what was implemented or who received adapted care. A few interventions (RO-DBT; (66); DBT; (55); a step-up day care programme for eating disorders; (54)) were rated as potentially encouraging masking of autistic traits/behaviours (e.g., encouraging neurotypical communication styles or participation without planning), which can have a negative impact on mental health and wellbeing (128). Previous research with professionals shows limited adaptation of mental healthcare for autistic people may reflect barriers including constrained resources, inflexible systems, clinician anxiety or uncertainty, and a need for more guidance, training, and practical tools (129,130).

Few studies in this review examined outcomes for different subgroups of autistic people, and some groups were consistently under-represented or excluded. For example, only 9/39 (23.1%) studies included autistic people with intellectual disabilities, and only 15/39 (38.5%) reported the racial or ethnic composition of their samples. Identification of autistic participants also varied substantially across studies. Few studies used both autism screening and diagnostic assessment. Most relied on existing diagnoses, increasing the risk of misclassification, while studies using screening measures often did not statistically compare HAT and LAT group screening scores, limiting confidence that groups were meaningfully distinct. Together, these limitations restrict the generalisability of the findings and make it difficult to determine whether outcomes differ across autistic populations. Systematically examining differences in outcomes between subgroups, particularly for marginalised and underserved groups, could help better understand mechanisms underlying treatment outcomes and inform more equitable care.

### 4.2. Strengths and limitations

To our knowledge, this is the first systematic review comparing mental health treatment outcomes between autistic and non-autistic people, with a broad scope across all ages, populations, interventions, settings, countries, and outcomes. The working group included academics, clinicians, a policymaker, and people with lived experience of mental health difficulties and/or caring for someone with such difficulties, including several autistic individuals. Members of the working group contributed to all stages of the review, enhancing the relevance, interpretation, and applicability of the findings. Comprehensive searches of academic and grey literature were conducted, with full double independent screening, data extraction, and quality appraisal. Autism-informed quality appraisal was applied using the AIRA, with at least one rating for each study completed by an autistic researcher.

Where feasible, quantitative data were synthesised via meta-analysis, but it was only possible to conduct one meta-analysis due to substantial heterogeneity across studies (i.e. differences in populations, interventions, settings, outcomes, and methods). In this meta-analysis, only post-intervention OCD symptom scores could be compared between groups, rather than pre-post change scores. However, across the three studies, no baseline differences in OCD symptoms were reported between the autistic and non-autistic groups, suggesting the groups were broadly comparable at baseline, supporting interpretation of post-intervention differences.

Additional limitations include the absence of a formal certainty assessment of the evidence. Although GRADE was pre-specified in the protocol, the substantial heterogeneity across studies made this challenging to apply, and an alternative framework was not undertaken. Also, three studies excluded participants with formal autism diagnoses while comparing groups above and below cut-off thresholds on autism screening measures (53,78,79). Although this met our eligibility criteria, these findings may be less generalisable to clinically diagnosed autistic populations. The review was also constrained by limitations of the literature, including many studies with small sample sizes and limited reporting of statistical power, as well as limited adjustment for confounders, lack of robustness in methods used to categorise autistic and non-autistic people, inadequate reporting of intervention content and delivery, and infrequent use of outcome measures validated or adapted for autistic people.

### 4.3. Implications for research, policy, and practice

The variability in outcomes identified in this review suggests that disparities in mental health treatment outcomes between autistic and non-autistic people are likely in at least some treatment settings. Where differences were identified, the direction of effects generally indicated poorer treatment outcomes among autistic people compared with non-autistic people. Many of the studies that did report similar outcomes had small samples and did not include power calculations or were under-powered, very much limiting confidence in these findings. However, more consistent evidence of poorer outcomes for autistic people from larger, higher-quality studies was observed for NHS Talking Therapies services, CBT for OCD, emergency departments, antipsychotic treatment in early psychosis, mixed specialist eating disorder services (covering combinations of inpatient, step-up day patient, day patient care, and specialist treatment centres), and CAMHS services for CYP with OCD. These areas may therefore represent priority settings for considering improvements to care. However, limited or inconsistent evidence in other settings should not be interpreted as evidence of no disparities, as this may be due to a lack of high-quality studies in these areas. The evidence base may also underestimate disparities, as autistic people who face barriers to accessing or engaging with mental health interventions are likely to be under-represented in treatment outcome studies.

Large, high-quality studies are needed that compare outcomes for autistic and non-autistic people across the full range of treatments and settings, with sufficient statistical power, longer follow-ups, appropriate adjustment for confounders, and more robust classification of autistic and non-autistic groups. A wider range of outcomes should also be examined, including self-harm, suicidal thoughts and behaviours, quality of life, treatment satisfaction, restrictive practices, and iatrogenic harms. Greater attention is also needed to trajectories of response, predictors of outcomes, and how these vary across interventions, settings, and populations. Routine data collection on autism within healthcare datasets, alongside relevant demographic and clinical characteristics, could support larger, more representative evaluations of treatment outcomes, enabling subgroup analyses and more robust assessment of disparities.

Improving the inclusiveness of research is also essential. This includes involving autistic people in the design, implementation, and evaluation of interventions and studies; using outcome measures that are valid and meaningful for autistic populations; adapting data collection methods to facilitate participation; and ensuring diverse and representative samples. More research is needed exploring outcomes among marginalised and underserved autistic groups (e.g., people with co-occurring intellectual disabilities, individuals with minimal verbal communication, people from ethnic minority backgrounds, people identifying as lesbian, gay, bisexual, transgender, or queer (LGBTQ+), or people with limited proficiency in the language(s) used to deliver services), including the impact of intersectional factors on access, experiences, and outcomes (26,27,29,31,32,131). Qualitative and realist approaches may be particularly valuable in understanding reasons for differences in outcomes, and identifying what works, for whom, and in which contexts. This could help inform improvements to make mental health care more inclusive and effective for all autistic people.

The differences in mental health treatment outcomes reported in this review – including smaller improvements in mental health and functioning, higher rates of self-harm and suicidal thoughts and behaviours, and higher use of acute services and restrictive practices for autistic people in some contexts – suggest that current provision does not always adequately meet autistic people’s needs. While efforts to reduce inappropriate psychiatric admissions and restrictive practices for autistic people are important, higher rates of acute service use may also indicate substantial mental health needs and unmet needs within existing services. Therefore, reducing reliance on restrictive care should be accompanied by ensuring that autistic people can access appropriate, effective mental health care when clinically indicated.

This points to the need for more adapted and tailored approaches to care. More broadly, individualised, person-centred care is widely recognised as a core principle of good clinical practice (132–135), and tailoring care for autistic people aligns with this. UK legislation (136,137), national policy and strategy (9,35–37,138), and clinical guidelines (99,100) already emphasise that services should be accessible and responsive to autistic people, including through provision of reasonable adjustments to support equitable access. Given evidence that many relatively simple adaptations are feasible and acceptable (45,46), and that a range of autism-informed approaches have already been developed, there may be value in greater implementation and evaluation of autism-informed approaches within routine mental health services. Further evaluation of autism-informed approaches currently used in routine clinical practice should examine their effectiveness and explore experiences of autistic service users, families/carers, and clinicians, alongside barriers and facilitators to implementation.

At a system level, sustained implementation of autism-informed care is more likely where consideration of neurodivergence is embedded within service design and delivery, supported by leadership, clear frameworks, and adequate resourcing. There is also considerable overlap with trauma-informed approaches, including shared principles of flexibility in service delivery, consistency and continuity of care, collaboration, personalised care, and attention to the care environment (139). Staff training on autism across professional groups – both in pre-registration education and in-service training, and ideally co-developed and co-delivered with autistic people – may support more consistent implementation of autism-informed care (9,24,99,100,134). Collaboration between general mental health services and services and organisations with expertise in autism may also support the ongoing upskilling of staff, contributing to more sustainable, system-wide change beyond training initiatives.

Finally, future research and service developments should be co-produced with, and where possible led by, autistic people, including those from marginalised and underserved groups, to better reflect their needs and priorities. Approaches such as James Lind Alliance priority setting partnerships could help identify priorities for future research (140). Meaningful involvement of family members/carers, clinicians, policymakers, and researchers, will be important in supporting coordinated efforts to improve mental health outcomes and experiences for autistic people.

## 5. Conclusions

This systematic review shows that mental health treatment outcomes for autistic compared with non-autistic people vary across interventions and different types of outcomes, with evidence generally indicating comparable or smaller improvements in the autistic group rather than a lack of benefit. However, confidence in these findings is generally limited by variable study quality, small sample sizes, and widespread limitations in the inclusiveness of research for autistic people. Action is needed to improve the accessibility and appropriateness of mental health care for autistic people, supported by larger, more inclusive high-quality quantitative studies exploring differences in outcomes between autistic and non-autistic people, including variation across subgroups. This could be complemented by qualitative and realist approaches to help understand what works, for whom, in which contexts, and why, to inform improvements to care. Future research and developments in mental health care should be co-produced with autistic people to help ensure they are acceptable and responsive to their needs.

## 6. Lived experience commentary by Rachel Rowan Olive and Danielle Baptiste

The use of the Autism Inclusive Research Assessment (AIRA) in this review highlights the gap between frameworks for inclusive autism research and research practice: most studies scored poorly, suggesting the field does not meaningfully reflect autistic perspectives and priorities. It is also noteworthy that highest risks of encouraging autistic masking were identified only in studies examining Dialectical Behaviour Therapy (DBT). This is consistent with the lived experience of one of the commentators: DBT made it easier to identify and communicate emotions, but also fundamentally misunderstood key aspects of their autistic experiences and needs. They are still unpacking harm from this in more neuro-affirming therapy a decade later.

This review suggests that autistic people may make less progress in psychological treatment than non-autistic people, although important limitations in the quality of the evidence must be considered. One explanation may be that therapies are not sufficiently adapted for autistic people or designed with autistic people in mind. However, another possibility is minority stress: the stress associated with living in a society that is not designed around one’s experiences and needs (119). If a person leaves the therapy room and returns to a life characterised by greater social stress, exclusion, discrimination, or a lack of appropriate support, this may limit the extent to which they are able to benefit from therapy. Importantly, minority stress will not be experienced in the same way by all autistic people. Autistic people who are also racialised, living in socioeconomic disadvantage, LGBTQ+, or who experience other forms of marginalisation may encounter additional or different stressors that shape their mental health and experiences of therapy. Intersectionality (141) must therefore be considered when examining mental health outcomes and intervention effectiveness.

The limited reporting of ethnicity and socioeconomic data, alongside the exclusion of those with intellectual disabilities, raises further questions about whose experiences are represented when interventions are developed and evaluated. We want more inclusive therapy but also a more inclusive society, alongside research that makes space for those perspectives overlooked by purely quantitative, outcomes-focused approaches.

## List of abbreviations

ACE: Accident and Emergency
ADHD: Attention-Deficit/Hyperactivity Disorder
AIRA: Autism-Inclusive Research Assessment
AQ-50: Autism Quotient-50
BMI: Body Mass Index
BPD: Borderline Personality Disorder
CAMHS: Child and Adolescent Mental Health ServicesC
ASP: Critical Appraisal Skills Programme
CBT: Cognitive Behavioural Therapy
CORE: Clinical Outcomes in Routine Evaluation
CREST: Cognitive Remediation and Emotional Skills Training
CYP: Children and Young People
DBT: Dialectical Behaviour Therapy
DSM-IV: Diagnostic and Statistical Manual of Mental Disorders, Fourth Edition
DSM-5: Diagnostic and Statistical Manual of Mental Disorders, Fifth Edition
ED: Emergency Department
EIP: Early Intervention in Psychosis
FT-AN: Family Therapy for Anorexia Nervosa
HAT: High Autistic Traits;
GAD-7: Generalized Anxiety Disorder-7
GAF: Global Assessment of Functioning
HoNOS: Health of the Nation Outcome Scales
IAPT: Improving Access to Psychological Therapies
ICD-10: International Classification of Diseases, Tenth Revision
ICD-11: International Classification of Diseases, Eleventh Revision
LAT: Low Autistic Traits
LGBTQ+: Lesbian, Gay, Bisexual, Transgender, or Queer
MHPRU: Policy Research Unit in Mental Health
N/A: Not Applicable
NHS: National Health Service
NIHR: National Institute for Health and Care Research
OCD: Obsessive-Compulsive Disorder
OQ-45: Outcome Questionnaire-45
PEACE: Pathway for Eating Disorders and Autism developed from Clinical Experience
PHQ-9: Patient Health Questionnaire-9
PRISMA: Preferred Reporting Items for Systematic Review and Meta-Analyses
PRN: Pro Re Nata
RCT: Randomised Controlled Trial
RO-DBT: Radically Open Dialectical Behaviour Therapy
SAS: Social Aptitude Scale
SMD: Standardised Mean Differences
UK: United Kingdom
Y-BOCS: Yale Brown Obsessive Compulsive Scale

## 8. Declarations

### Ethics approval and consent to participate

Not applicable.

### Consent for publication

Not applicable.

### Availability of data and materials

All data generated or analysed during this study are included in this published article and its supplementary information files.

## Funding

This study was funded by the National Institute for Health and Care Research (NIHR) Policy Research Programme (grant no. NIHR206125). The current study represents independent research funded in part by the National Institute for Health and Care Research (NIHR) Maudsley Biomedical Research Centre at South London and Maudsley NHS Foundation Trust and King’s College London. For JO, this paper represents independent research funded by the Wellcome Trust [308556/Z/23/Z].

The funders had no involvement in study design, data collection, analysis, interpretation or the decision to submit for publication. The views expressed are those of the author(s) and not necessarily those of the funders or the Department of Health and Social Care.

### Authors’ contributions

All authors, except KH and MC, substantially contributed to the conception or design of this study (JG, FE, JT, SL, RRO, AB, WL, MH, MM, DB, RA, PB, LG, HKL, PN, JEC, WM, JO, VT, KT, BLE, AS, SJ). Literature searching was conducted by JG and PB. Title and abstract screening was conducted by JG, FE, JT, KT. Full text screening was conducted by JG, FE, JT. Data extraction and quality appraisal were conducted by JG, FE, JT, KH, MC, SL. Evidence synthesis was led by JG and FE and supported by JT, SL, RRO, AB, WL, MH, MM, DB, RA, PB, LG, HKL, PN, JEC, WM, JO, VT, KT, BLE, AS, SJ. JG and FE led on drafting the manuscript with input and/or editing by all other authors (JT, SL, RRO, AB, WL, MH, MM, DB, RA, PB, MC, LG, KH, HKL, PN, JEC, WM, JO, VT, KT, BLE, AS, SJ). RRO and DB wrote the lived experience commentary. All authors read and approved the final manuscript (JG, FE, JT, SL, RRO, AB, WL, MH, MM, DB, RA, PB, MC, LG, KH, HKL, PN, JEC, WM, JO, VT, KT, BLE, AS, SJ).

## Supporting information

Additional File 2 (data extraction form template)

Additional File 3 (CASP and AIRA ratings)

## Acknowledgements

Not applicable.

## 10. Additional files

### Additional File 1 (Word document; .docx)

Appendices. Contains appendices, including PRISMA checklists; protocol deviations and justifications; search strategy and search terms; excluded full texts and reasons for exclusion; and a list of outcome measures reported on in this review with accompanying references.

### Additional File 2 (Excel file; .xlsx)

Data extraction form template. Excel file showing the data extraction form template used to systematically extract data from included studies.

### Additional File 3 (Excel File; .xlsx)

Critical Appraisal Skills Programme (CASP) and Autism-Inclusive Research Assessment (AIRA) scores. Excel file containing the CASP and AIRA items, scoring criteria, and full details of ratings.

