## Additional File 1 (Appendices) for "Comparing mental health treatment outcomes for autistic and non-autistic people: a systematic review"

### **Appendix A.** PRISMA checklists.

| Section and Topic | Item # | Checklist item | Location where item is reported |
|---|---|---|---|
| **TITLE** | | | |
| Title | 1 | Identify the report as a systematic review. | Page 1 |
| **ABSTRACT** | | | |
| Abstract | 2 | See the PRISMA 2020 for Abstracts checklist. | See table below |
| **INTRODUCTION** | | | |
| Rationale | 3 | Describe the rationale for the review in the context of existing knowledge. | Pages 4-6 |
| Objectives | 4 | Provide an explicit statement of the objective(s) or question(s) the review addresses. | Page 6 |
| **METHODS** | | | |
| Eligibility criteria | 5 | Specify the inclusion and exclusion criteria for the review and how studies were grouped for the syntheses. | Pages 10-12, 15 |
| Information sources | 6 | Specify all databases, registers, websites, organisations, reference lists and other sources searched or consulted to identify studies. Specify the date when each source was last searched or consulted. | Pages 9-10 |
| Search strategy | 7 | Present the full search strategies for all databases, registers and websites, including any filters and limits used. | Appendix B, Additional File 1 |
| Selection process | 8 | Specify the methods used to decide whether a study met the inclusion criteria of the review, including how many reviewers screened each record and each report retrieved, whether they worked independently, and if applicable, details of automation tools used in the process. | Pages 10-11 |
| Data collection process | 9 | Specify the methods used to collect data from reports, including how many reviewers collected data from each report, whether they worked independently, any processes for obtaining or confirming data from study investigators, and if applicable, details of automation tools used in the process. | Page 12 |
| Data items | 10a | List and define all outcomes for which data were sought. Specify whether all results that were compatible with each outcome domain in each study were sought (e.g. for all measures, time points, analyses), and if not, the methods used to decide which results to collect. | Page 12 |
| | 10b | List and define all other variables for which data were sought (e.g. participant and intervention characteristics, funding sources). Describe any assumptions made about any missing or unclear information. | Additional File 2 |
| Study risk of bias assessment | 11 | Specify the methods used to assess risk of bias in the included studies, including details of the tool(s) used, how many reviewers assessed each study and whether they worked independently, and if applicable, details of automation tools used in the process. | Pages 13-15 |

| Section and Topic | Item # | Checklist item | Location where item is reported |
|---|---|---|---|
| Effect measures | 12 | Specify for each outcome the effect measure(s) (e.g. risk ratio, mean difference) used in the synthesis or presentation of results. | Page 15 |
| Synthesis methods | 13a | Describe the processes used to decide which studies were eligible for each synthesis (e.g. tabulating the study intervention characteristics and comparing against the planned groups for each synthesis (item #5)). | Page 15 |
| | 13b | Describe any methods required to prepare the data for presentation or synthesis, such as handling of missing summary statistics, or data conversions. | Page 15 |
| | 13c | Describe any methods used to tabulate or visually display results of individual studies and syntheses. | Page 15 |
| | 13d | Describe any methods used to synthesize results and provide a rationale for the choice(s). If meta-analysis was performed, describe the model(s), method(s) to identify the presence and extent of statistical heterogeneity, and software package(s) used. | Page 15 |
| | 13e | Describe any methods used to explore possible causes of heterogeneity among study results (e.g. subgroup analysis, meta-regression). | N/A |
| | 13f | Describe any sensitivity analyses conducted to assess robustness of the synthesized results. | N/A |
| Reporting bias assessment | 14 | Describe any methods used to assess risk of bias due to missing results in a synthesis (arising from reporting biases). | N/A |
| Certainty assessment | 15 | Describe any methods used to assess certainty (or confidence) in the body of evidence for an outcome. | N/A |
| **RESULTS** | | | |
| Study selection | 16a | Describe the results of the search and selection process, from the number of records identified in the search to the number of studies included in the review, ideally using a flow diagram. | Pages 16-17 (including Figure 1) |
| | 16b | Cite studies that might appear to meet the inclusion criteria, but which were excluded, and explain why they were excluded. | Appendix C, Additional File 1 |
| Study characteristics | 17 | Cite each included study and present its characteristics. | Table 1 and pages 16-18 |
| Risk of bias in studies | 18 | Present assessments of risk of bias for each included study. | Tables 2 & 3 and pages 18-21 |
| Results of individual studies | 19 | For all outcomes, present, for each study: (a) summary statistics for each group (where appropriate) and (b) an effect estimate and its precision (e.g. confidence/credible interval), ideally using structured tables or plots. | Tables 5 & 6 |
| Results of syntheses | 20a | For each synthesis, briefly summarise the characteristics and risk of bias among contributing studies. | Tables 5 & 6 and pages 22-40 |

| Section and Topic | Item # | Checklist item | Location where item is reported |
|---|---|---|---|
| | 20b | Present results of all statistical syntheses conducted. If meta-analysis was done, present for each the summary estimate and its precision (e.g. confidence/credible interval) and measures of statistical heterogeneity. If comparing groups, describe the direction of the effect. | Pages 23-24 |
| | 20c | Present results of all investigations of possible causes of heterogeneity among study results. | N/A |
| | 20d | Present results of all sensitivity analyses conducted to assess the robustness of the synthesized results. | N/A |
| Reporting biases | 21 | Present assessments of risk of bias due to missing results (arising from reporting biases) for each synthesis assessed. | N/A |
| Certainty of evidence | 22 | Present assessments of certainty (or confidence) in the body of evidence for each outcome assessed. | N/A |
| **DISCUSSION** | | | |
| Discussion | 23a | Provide a general interpretation of the results in the context of other evidence. | Pages 40-46 |
| | 23b | Discuss any limitations of the evidence included in the review. | Pages 45-47 |
| | 23c | Discuss any limitations of the review processes used. | Pages 46-47 |
| | 23d | Discuss implications of the results for practice, policy, and future research. | Pages 48-51 |
| **OTHER INFORMATION** | | | |
| Registration and protocol | 24a | Provide registration information for the review, including register name and registration number, or state that the review was not registered. | Pages 3, 7 |
| | 24b | Indicate where the review protocol can be accessed, or state that a protocol was not prepared. | Pages 3, 7 |
| | 24c | Describe and explain any amendments to information provided at registration or in the protocol. | |
| Support | 25 | Describe sources of financial or non-financial support for the review, and the role of the funders or sponsors in the review. | Page 56 |
| Competing interests | 26 | Declare any competing interests of review authors. | Page 56 |
| Availability of data, code and other materials | 27 | Report which of the following are publicly available and where they can be found: template data collection forms; data extracted from included studies; data used for all analyses; analytic code; any other materials used in the review. | Additional File 2, page 55 |

*From:* Page MJ, McKenzie JE, Bossuyt PM, Boutron I, Hoffmann TC, Mulrow CD, et al. The PRISMA 2020 statement: an updated guideline for reporting systematic reviews. BMJ 2021;372:n71. doi: 10.1136/bmj.n71. This work is licensed under CC BY 4.0. To view a copy of this license, visit https://creativecommons.org/licenses/by/4.0/

| Section and Topic | Item # | Checklist item | Reported (Yes/No) |
|---|---|---|---|
| **TITLE** | | | |
| Title | 1 | Identify the report as a systematic review. | Yes |
| **BACKGROUND** | | | |
| Objectives | 2 | Provide an explicit statement of the main objective(s) or question(s) the review addresses. | Yes |
| **METHODS** | | | |
| Eligibility criteria | 3 | Specify the inclusion and exclusion criteria for the review. | Yes |
| Information sources | 4 | Specify the information sources (e.g. databases, registers) used to identify studies and the date when each was last searched. | Yes |
| Risk of bias | 5 | Specify the methods used to assess risk of bias in the included studies. | Yes |
| Synthesis of results | 6 | Specify the methods used to present and synthesise results. | Yes |
| **RESULTS** | | | |
| Included studies | 7 | Give the total number of included studies and participants and summarise relevant characteristics of studies. | Yes |
| Synthesis of results | 8 | Present results for main outcomes, preferably indicating the number of included studies and participants for each. If meta-analysis was done, report the summary estimate and confidence/credible interval. If comparing groups, indicate the direction of the effect (i.e. which group is favoured). | Yes |
| **DISCUSSION** | | | |

| Section and Topic | Item # | Checklist item | Reported (Yes/No) |
|---|---|---|---|
| Limitations of evidence | 9 | Provide a brief summary of the limitations of the evidence included in the review (e.g. study risk of bias, inconsistency and imprecision). | Yes |
| Interpretation | 10 | Provide a general interpretation of the results and important implications. | Yes |
| **OTHER** | | | |
| Funding | 11 | Specify the primary source of funding for the review. | ‘Funding’ section of manuscript (page 123) |
| Registration | 12 | Provide the register name and registration number. | Yes |

*From:* Page MJ, McKenzie JE, Bossuyt PM, Boutron I, Hoffmann TC, Mulrow CD, et al. The PRISMA 2020 statement: an updated guideline for reporting systematic reviews. BMJ 2021;372:n71. doi: 10.1136/bmj.n71. This work is licensed under CC BY 4.0. To view a copy of this license, visit https://creativecommons.org/licenses/by/4.0/

### **Appendix B.** Protocol deviations and justification.

In our protocol, we had planned to assess the certainty of the evidence using GRADE ratings (Schünemann et al., 2013). However, applying certainty ratings to complex interventions can be challenging (Montgomery et al., 2019). The interventions/services captured in this review often involved multiple components delivered flexibly, and studies often lacked detail about what support participants received. This makes it difficult to judge deviations from intended interventions. We therefore focused instead on assessing the certainty of the reported effects based on study design and consistency in the direct of effects relative to the null.

### **Appendix C.** Search strategy and search terms.

#### Academic literature search results

All academic database searches were limited to articles published in 1992 onwards. No language or location restrictions were applied.

| Academic database | Records identified in initial search | Results identified in updated search (30/04/26) |
|---|---|---|
| MEDLINE via Ovid | 235 (7th April 2025) | 61 |
| Embase via Ovid | 406 (7th April 2025) | 178 |
| PsycINFO via Ovid | 242 (7th April 2025) | 53 |
| CINAHL | 104 (7th April 2025) | 10 |
| PubMed | 382 (11th April 2025) | 97 |
| **Total** | 1,369 | 399 |

#### Academic database search terms

**MEDLINE, Embase, PsycINFO (all via Ovid)**

1. (autis* or Asperger* or ASC or ASD or PDD or "pervasive development*" or Kanner* or "childhood disintegrative disorder" or "PPD-NOS").mp.
2. ("mental health treatment*" or "mental health service*" or "health service*" or hospital or inpatient* or "community mental health" or "home treatment" or "crisis resolution" or "child and adolescent mental health service*" or CAMHS or CBT or "cognitive behavio* therap*" or DBT or "dialectical behavio* therap*" or "family therapy" or "interpersonal therap*" or psychodynamic or treatment* or intervention* or "occupational therapy" or mindfulness or "dynamic support register*" or "care and treatment review*" or "section 17 aftercare" or "employment support" or IPS or "social prescribing" or "care coordinat*" or "care co-ordinat*" or "case manage*" or "debt advice" or "psychological intervention*" or "psychological therap*" or "behavio* therap*" or psychotherap* or "acceptance and commitment therap*" or ACT or psychotropic medication* or antidepressant* or antipsychotic* or counselling or "peer support").mp.
3. ("mental health" or "mental illness*" or "mental disorder*" or "mental condition*" or "mental distress" or "mental wellbeing" or "mental well-being" or "psychological disorder*" or "psychological wellbeing" or "psychological well-being" or "psychological condition*" or anxi* or affective or depress* or "behavio* problem*" or "eating disorder*" or psychosis or psychotic or "bipolar disorder" or "impulse control disorder*" or "conduct disorder*" or "dissociative disorder*" or "feeding disorder*" or "mood disorder*" or "personality disorder*" or schizophrenia or "somatoform disorder*" or "substance-related disorder"

or ptsd or "post-traumatic stress disorder" or "trauma disorder*" or anorexia or bulimia or arfid).mp.

4. ((compar* or "control group*" or "matched group*" or "matched control*" or "case-control" or cohort) ADJ3 ("without autism" or "neurotypical" or "general population" or "non-autistic" or "healthy control* or allistic")).mp.
5. 1 and 2 and 3 and 4
6. limit 5 to yr="1992-Current”

**CINAHL (searched title and abstract)**
(autis* or Asperger* or ASC or ASD or PDD or "pervasive development*” or Kanner* or “childhood disintegrative disorder” or “PPD-NOS") AND ("mental health treatment*" or "mental health service*" or "health service*" or hospital or inpatient* or "community mental health" or "home treatment" or "crisis resolution" or "child and adolescent mental health service*" or CAMHS or CBT or "cognitive behavio* therap*" or DBT or "dialectical behavio* therap*" or "family therapy" or "interpersonal therap*" or psychodynamic or treatment* or intervention* or "occupational therapy" or mindfulness or “dynamic support register*” or “care and treatment review*” or “section 17 aftercare” or “employment support” or “IPS” or “social prescribing” or “care coordinat*” or “care co-ordinat*” or “case manage*” or “debt advice” or "psychological intervention*" or "psychological therap*" or "behavio* therap*" or psychotherap* or "acceptance and commitment therap*" or ACT or psychotropic medication* or antidepressant* or antipsychotic* or counselling or "peer support") AND ("mental health" or "mental illness*" or "mental disorder*" or "mental condition*" or “mental distress” or “mental wellbeing” or “mental well-being” or “psychological disorder*” or “psychological wellbeing” or “psychological well-being” or “psychological condition*” or anxi* or affective or depress* or "behavio* problem*" or "eating disorder*" or psychosis or psychotic or “bipolar disorder” or “impulse control disorder*” or “conduct disorder*” or “dissociative disorder*” or “feeding disorder*” or “mood disorder*” or “personality disorder*” or schizophrenia or “somatoform disorder*” or “substance-related disorder” or ocd or “obsessive-compulsive disorder” or “obsessive compulsive disorder” or ptsd or “post-traumatic stress disorder” or “trauma disorder*” or anorexia or bulimia or arfid) AND (compar* or "control group*" or "matched group*" or "matched control*" or “matched with” or “case-control” or cohort) N5 ("without autism" or “without identified autism” or “without ASD” or “no ASD” or “typically developing” or "neurotypical" or "general population" or "non-autistic" or "healthy control* or allistic")

**PubMed**
(autis*[Title/Abstract] OR Asperger*[Title/Abstract] OR ASC[Title/Abstract] OR ASD[Title/Abstract] OR PDD[Title/Abstract] OR "pervasive development*"[Title/Abstract] OR Kanner*[Title/Abstract] OR "childhood disintegrative disorder"[Title/Abstract] OR "PPD-NOS") AND ("mental health treatment*"[Title/Abstract] OR "mental health

service*"[Title/Abstract] OR "health service*"[Title/Abstract] OR hospital[Title/Abstract] OR inpatient*[Title/Abstract] OR "community mental health"[Title/Abstract] OR "home treatment"[Title/Abstract] OR "crisis resolution"[Title/Abstract] OR "child and adolescent mental health service*"[Title/Abstract] OR CAMHS[Title/Abstract] OR CBT[Title/Abstract] OR "cognitive behavio* therap*"[Title/Abstract] OR DBT[Title/Abstract] OR "dialectical behavio* therap*"[Title/Abstract] OR "family therapy"[Title/Abstract] OR "interpersonal therap*"[Title/Abstract] OR psychodynamic[Title/Abstract] OR treatment*[Title/Abstract] OR intervention*[Title/Abstract] OR "occupational therapy"[Title/Abstract] OR mindfulness[Title/Abstract] OR "dynamic support register*"[Title/Abstract] OR "care and treatment review*"[Title/Abstract] OR "section 17 aftercare"[Title/Abstract] OR "employment support"[Title/Abstract] OR "IPS"[Title/Abstract] OR "social prescribing"[Title/Abstract] OR "care coordinat*"[Title/Abstract] OR "care co-ordinat*"[Title/Abstract] OR "case manage*"[Title/Abstract] OR "debt advice"[Title/Abstract] OR "psychological intervention*"[Title/Abstract] OR "psychological therap*"[Title/Abstract] OR "behavio* therap*"[Title/Abstract] OR psychotherap*[Title/Abstract] OR "acceptance and commitment therap*"[Title/Abstract] OR ACT[Title/Abstract] OR psychotropic medication*[Title/Abstract] OR antidepressant*[Title/Abstract] OR antipsychotic*[Title/Abstract] OR counselling[Title/Abstract] OR "peer support") AND ("mental health"[Title/Abstract] OR "mental illness*"[Title/Abstract] OR "mental disorder*"[Title/Abstract] OR "mental condition*"[Title/Abstract] OR "mental distress"[Title/Abstract] OR "mental wellbeing"[Title/Abstract] OR "mental well-being"[Title/Abstract] OR "psychological disorder*"[Title/Abstract] OR "psychological wellbeing"[Title/Abstract] OR "psychological well-being"[Title/Abstract] OR "psychological condition*"[Title/Abstract] OR anxi*[Title/Abstract] OR affective[Title/Abstract] OR depress*[Title/Abstract] OR "behavio* problem*"[Title/Abstract] OR "eating disorder*"[Title/Abstract] OR psychosis[Title/Abstract] OR psychotic[Title/Abstract] OR "bipolar disorder"[Title/Abstract] OR "impulse control disorder*"[Title/Abstract] OR "conduct disorder*"[Title/Abstract] OR "dissociative disorder*"[Title/Abstract] OR "feeding disorder*"[Title/Abstract] OR "mood disorder*"[Title/Abstract] OR "personality disorder*"[Title/Abstract] OR schizophrenia[Title/Abstract] OR "somatoform disorder*"[Title/Abstract] OR "substance-related disorder"[Title/Abstract] OR ocd[Title/Abstract] OR "obsessive-compulsive disorder"[Title/Abstract] OR "obsessive compulsive disorder"[Title/Abstract] OR ptsd[Title/Abstract] OR "post-traumatic stress disorder"[Title/Abstract] OR "trauma disorder*"[Title/Abstract] OR anorexia[Title/Abstract] OR bulimia[Title/Abstract] OR arfid[Title/Abstract]) AND (compar*[Title/Abstract] OR "control group*"[Title/Abstract] OR "matched group*"[Title/Abstract] OR "matched control*"[Title/Abstract] OR "matched with"[Title/Abstract] OR "case-control" or cohort) AND ("without autism"[Title/Abstract]

OR "without identified autism"[Title/Abstract] OR "without ASD"[Title/Abstract] OR "no ASD"[Title/Abstract] OR "typically developing"[Title/Abstract] OR "neurotypical"[Title/Abstract] OR "general population"[Title/Abstract] OR "non-autistic"[Title/Abstract] OR "healthy control* or allistic"[Title/Abstract])

### Grey literature search results

| Grey literature search | Records identified in initial search | Records identified in updated search (30/04/26) |
|---|---|---|
| HMIC | 3 (9th April 2025) | 2 |
| Pre print servers (Medrxiv & PsyArxiv) | 352 (9th April 2025) | 237 |
| ISRCTN[1] | 12 (9th April 2025) | 0 |
| Google[1,2] | 4 (9th April 2025) | 1 |
| Google scholar[1,2] | 8 (10th April 2025) | 1 |
| **Total** | 379 | 241 |

1: Only records identified as potentially relevant based on title were taken forward for formal screening at title and abstract level
2: Only the first 10 pages of results were viewed

### Grey literature search terms

**HMIC**

1. (autis* or Asperger* or ASC or ASD or PDD or "pervasive development*" or Kanner* or "childhood disintegrative disorder" or "PPD-NOS").mp.
2. ("mental health treatment*" or "mental health service*" or "health service*" or hospital or inpatient* or "community mental health" or "home treatment" or "crisis resolution" or "child and adolescent mental health service*" or CAMHS or CBT or "cognitive behavio* therap*" or DBT or "dialectical behavio* therap*" or "family therapy" or "interpersonal therap*" or psychodynamic or treatment* or intervention* or "occupational therapy" or mindfulness or "dynamic support register*" or "care and treatment review*" or "section 17 aftercare" or "employment support" or IPS or "social prescribing" or "care coordinat*" or "care co-ordinat*" or "case manage*" or "debt advice" or "psychological intervention*" or "psychological therap*" or "behavio* therap*" or psychotherap* or "acceptance and commitment therap*" or ACT or psychotropic medication* or antidepressant* or antipsychotic* or counselling or "peer support").mp.

3. ("mental health" or "mental illness*" or "mental disorder*" or "mental condition*" or "mental distress" or "mental wellbeing" or "mental well-being" or "psychological disorder*" or "psychological wellbeing" or "psychological well-being" or "psychological condition*" or anxi* or affective or depress* or "behavio* problem*" or "eating disorder*" or psychosis or psychotic or "bipolar disorder" or "impulse control disorder*" or "conduct disorder*" or "dissociative disorder*" or "feeding disorder*" or "mood disorder*" or "personality disorder*" or schizophrenia or "somatoform disorder*" or ocd or "obsessive compulsive disorder" or "obsessive-compulsive disorder" or "substance-related disorder" or ptsd or "post-traumatic stress disorder" or "trauma disorder*" or anorexia or bulimia or arfid).mp.
4. ((compar* or "control group*" or "matched group*" or "matched control*" or "case-control" or "matched with" or cohort) adj5 ("without autism" or "without identified autism" or "without ASD" or "no ASD" or "typically developing" or "neurotypical" or "general population" or "non-autistic" or "healthy control*” or allistic)).mp.
5. 1 and 2 and 3 and 4
6. limit 5 to yr="1992 -Current"

**Medrxiv and PsyArxiv (via Europe PMC)**

(autis* or Asperger* or ASC or ASD or PDD or "pervasive development*" or Kanner* or "childhood disintegrative disorder" or "PPD-NOS") AND ("mental health treatment*" or "mental health service*" or "health service*" or hospital or inpatient* or "community mental health" or "home treatment" or "crisis resolution" or "child and adolescent mental health service*" or CAMHS or CBT or "cognitive behavio* therap*" or DBT or "dialectical behavio* therap*" or "family therapy" or "interpersonal therap*" or psychodynamic or treatment* or intervention* or "occupational therapy" or mindfulness or "dynamic support register*" or "care and treatment review*" or "section 17 aftercare" or "employment support" or IPS or "social prescribing" or "care coordinat*" or "care co-ordinat*" or "case manage*" or "debt advice" or "psychological intervention*" or "psychological therap*" or "behavio* therap*" or psychotherap* or "acceptance and commitment therap*" or ACT or psychotropic medication* or antidepressant* or antipsychotic* or counselling or "peer support") AND ("mental health" or "mental illness*" or "mental disorder*" or "mental condition*" or "mental distress" or "mental wellbeing" or "mental well-being" or "psychological disorder*" or "psychological wellbeing" or "psychological well-being" or "psychological condition*" or anxi* or affective or depress* or "behavio* problem*" or "eating disorder*" or psychosis or psychotic or "bipolar disorder" or "impulse control disorder*" or "conduct disorder*" or "dissociative disorder*" or "feeding disorder*" or "mood disorder*" or "personality disorder*" or schizophrenia or "somatoform disorder*" or ocd or "obsessive compulsive disorder" or "obsessive-compulsive disorder" or "substance-related disorder" or ptsd or

"post-traumatic stress disorder" or "trauma disorder*" or anorexia or bulimia or arfid) AND ((compar* or "control group*" or "matched group*" or "matched control*" or "case-control" or "matched with" or cohort) AND ("without autism" or "without identified autism" or "without ASD" or "no ASD" or "typically developing" or "neurotypical" or "general population" or "non-autistic" or "healthy control* or allistic"))
Limit to preprints

**Google**
Autism AND "mental health" AND “treatment outcomes”: first 10 pages searched.
Search conducted 09/04/2025: One potentially relevant document taken forward.
Updated search limited to the past year (30/04/26): 0 potentially relevant documents taken forward.

Autism AND "mental health" AND "therapy outcomes": first 10 pages searched.
Search conducted 10/04/2025: 2 potentially relevant documents taken forward (additional document identified however flagged as a duplicate of a record identified through main database searching).
Updated search limited to the past year (30/04/26): 0 potentially relevant documents taken forward.

**Google Scholar**
Autism AND "mental health" AND “treatment outcomes”: first 10 pages searched.
Search conducted 10/04/2025: 8 potentially relevant document taken forward.
Updated search limited to 2025-2026 (30/04/26): 1 potentially relevant document taken forward.

Autism AND "mental health" AND "therapy outcomes": first 10 pages searched.
Search conducted 10/04/2025: 0 potentially relevant documents taken forward.
Updated search limited to 2025-2026 (30/04/26): 0 potentially relevant documents taken forward.

**ISRCTN** (searched 09/04/2025)
“Autism” in text, no limits. 260 results. 12 taken forward to screening
Updated search (30/04/26): 43 additional results. 0 taken forward to screening.

### **Appendix D.** Excluded full texts and reasons for exclusion.

| Number | Full text reference | Reason for exclusion |
|---|---|---|
| 1 | Anonymous. Pai.ACT: An Artificial Intelligence-Driven Chatbot-Assisted ACT Mobile Platform With Mental Health Triage - A Randomized Controlled Trial and Regional Service Evaluation. clinicaltrials.gov [Internet]. 2025. Located at: NCT06808555. Available from: https://clinicaltrials.gov/study/NCT06808555https://librarysearch.kcl.ac.uk/discovery/openurl?institution=44KCL_INST&vid=44KCL_INST:44KCL_INST&sid=OVID:embase&id=pmid:&id=doi:&genre=article&atitle=Pai.ACT%3A+An+Artificial+Intelligence-Driven+Chatbot-Assis | Trial not complete |
| 2 | Avrahami M., Ben-Dor D.H., Ratzon R., Weizman A., Perlman Danieli P. Characterizing the clinical and sociodemographic profiles of hospitalized adolescents with autism spectrum disorder. Global Mental Health. 2024;11((Avrahami, Ben-Dor, Ratzon, Perlman Danieli) Child and Adolescent Division, Geha Mental Health Center, Petah Tikva, Israel(Avrahami, Ben-Dor, Weizman) Faculty of Medicine, Tel Aviv University, Tel Aviv, Israel(Weizman) Laboratory of Molecular and Biologic):e63. Located at: 2032223603. doi:10.1017/gmh.2024.63 | Wrong patient population |
| 3 | Babb C., Brede J., Jones C.R.G., Serpell L., Mandy W., Fox J. A comparison of the eating disorder service experiences of autistic and non-autistic women in the UK. European Eating Disorders Review. 2022;30(5):616EP – 627. Located at: 2018105000. doi:10.1002/erv.2930 | Wrong outcomes |
| 4 | Barbuti M, Amadori S, De Rosa U, Perugi G. Clinical characterization of bipolar disorder in adults with autism without intellectual disability: a cross-sectional study. Journal of Affective Disorders. 2026;405(h3v, 7906073):121527. doi:10.1016/j.jad.2026.121527 | Wrong outcomes |
| 5 | Barrett B.F., Wehmeyer M., Kolb J., Ostermann T., Sappok T. Impact of the emotional development approach on psychotropic medication in adults with intellectual and developmental disabilities: a retrospective clinical analysis. Journal of Intellectual Disability Research. 2024;68(8):941EP – 953. Located at: 2029159856. doi:10.1111/jir.13136 | Wrong intervention |
| 6 | Başaran AS, Türkel NN, Koparal B, Kuruoğlu A. Exploring the role of autistic traits in treatment-resistant and clozapine-resistant schizophrenia: a comparative study. Frontiers in psychiatry. 2025;16(NA):1541469–1541469. doi:10.3389/fpsyt.2025.1541469 | Wrong comparator |

| | | |
|---|---|---|
| 7 | Beverly J, Giannouchos TV, Callaghan T. Examining frequent emergency department use among children and adolescents with autism spectrum disorder. Autism : the international journal of research and practice. 2021;25(5):1382–94. doi:10.1177/1362361321990925 | Wrong intervention |
| 8 | Calabrese M.E., Sideridis G., Weitzman C. Physical and Pharmacologic Restraint in Hospitalized Children With Autism Spectrum Disorder. Pediatrics. 2024;153(1):e2023062172. Located at: 2030480045. doi:10.1542/peds.2023-062172 | Wrong patient population |
| 9 | Cervantes PE, Brown DS, Horwitz SM. Suicidal ideation and intentional self-inflicted injury in autism spectrum disorder and intellectual disability: An examination of trends in youth emergency department visits in the United States from 2006 to 2014. Autism : the international journal of research and practice. 2022;27(1):226–43. doi:10.1177/13623613221091316 | Wrong intervention |
| 10 | Coventry, Warwickshire Partnership NHST. Behavioural interventions to treat anxiety in adults with autism and moderate to severe intellectual disabilities [Internet]. 2021. Available from: https://www.isrctn.com/ISRCTNISRCTN12637590 | Wrong comparator |
| 11 | Cox JC, Neeley-Tass SE. Psychotherapy Outcome for College Students with ASD and other Disabilities in a National Sample: Replication, Refinement, and Extension. Centre for Collegiate Mental Health Blog [Internet]. 2022;(09/04/2025). Available from: https://ccmh.psu.edu/index.php?option=com_dailyplanetblog&view=entry&year=2022&month=04&day=28&id=24:psychotherapy-outcome-for-college-students-with-asd-and-other-disabilities-in-a-national-sample-replication-refinement-and-extension | Wrong source type |
| 12 | Dandil Y, Smith K, Adamson J, Tchanturia K. Individual cognitive remediation therapy benefits for patients with anorexia nervosa and high autistic features. European Eating Disorders Review. 2020;28(1):87–91. doi:10.1002/erv.2707 | Wrong outcomes |
| 13 | Dickson KS, Kennedy SM, Safer J, Brookman-Frazee L, Roesch S, Anthony LG. Study protocol for a hybrid type 2 effectiveness-implementation trial of two interventions for autistic and non-autistic youth in children's mental health settings: one tailored for neurodivergence and one universal. Implement Sci Commun. 2025;6(1):131. Located at: ClinicalTrials.gov/NCT06651086. doi:10.1186/s43058-025-00799-6 | Trial not complete |
| 14 | Elizabethtown C. Exploring the impact of mindfulness on children with mental health diagnoses [Internet]. 2023. | Wrong outcomes |

| | | |
|---|---|---|
| | Available from: https://www.isrctn.com/ISRCTNISRCTN10476703 | |
| 15 | Foss-Feig JH, Velthorst E, Smith L, Reichenberg A, Addington J, Cadenhead KS, et al. Clinical Profiles and Conversion Rates Among Young Individuals With Autism Spectrum Disorder Who Present to Clinical High Risk for Psychosis Services. J Am Acad Child Adolesc Psychiatry. 2019;58(6):582–8. doi:10.1016/j.jaac.2018.09.446 | Wrong intervention |
| 16 | Giannouchos TV, Beverly J, Christodoulou I, Callaghan T. Suicide and non-fatal self-injury-related emergency department visits among individuals with autism spectrum disorder. Autism. 2023 Oct;27(7):1983-96. | Wrong intervention |
| 17 | Giombini L, Nesbitt S, Kusosa R, Fabian C, Sharia T, Easter A, Tchanturia K. Neuropsychological and clinical findings of Cognitive Remediation Therapy feasibility randomised controlled trial in young people with anorexia nervosa. European Eating Disorders Review. 2022 Jan;30(1):50-60. | Wrong outcomes |
| 18 | Gray K.M., Hu N., Tonge B.J., Sanders M., Sofronoff K., Einfeld S.L. A community-based parenting intervention for parents of children with a disability: Comparison of effectiveness for children with and without autism spectrum disorder. Journal of Intellectual Disability Research. 2018;62(8):667. Located at: 623203420. doi:10.1111/jir.12512 | Wrong source type |
| 19 | Guffanti A., Mazzoni F., Silva A., Calorio F., Politi P., Brondino N., et al. Depression and suicidal risk in autism spectrum disorder: efficacy of intranasal esketamine. Clinical experience from a public mental health service in Italy. European Psychiatry. 2025;68(Supplement 1):S590–1. Located at: 648908328. doi:10.1192/j.eurpsy.2025.1204 | Wrong source type |
| 20 | Hertfordshire Partnership University NHSFT. Investigating the feasibility of virtual reality cognitive behaviour therapy for anxiety in autistic young people [Internet]. 2023. Available from: https://www.isrctn.com/ISRCTNISRCTN10127813 | Wrong comparator |
| 21 | Hirvikoski T, Boman M, Chen Q, D'Onofrio BM, Mittendorfer-Rutz E, Lichtenstein P, et al. Individual risk and familial liability for suicide attempt and suicide in autism: a population-based study. Psychological medicine. 2019;50(9):1463–74. doi:10.1017/s0033291719001405 | Wrong intervention |
| 22 | Hull L, Stark I, Lundberg M, Ahlqvist VH, Nordström SI, Ohlis A, et al. Sex differences in self-harm and suicide in young autistic adults. Acta psychiatrica Scandinavica. 2024;150(4):223–33. doi:10.1111/acps.13736 | Wrong intervention |

| | | |
|---|---|---|
| 23 | Jaka S., Gautam A., Kaur S., Patel R. 3.26 Hospitalization-Related Factors and Outcomes in Pediatric Patients With ASD: Insights From a Cross-Sectional Study in the United States. Journal of the American Academy of Child and Adolescent Psychiatry. 2025;64(10 Supplement):S229. Located at: 2040738810. doi:10.1016/j.jaac.2025.08.236 | Wrong source type |
| 24 | Kalb LG, Vasa RA, Ballard ED, Woods S, Goldstein M, Wilcox HC. Epidemiology of Injury-Related Emergency Department Visits in the US Among Youth with Autism Spectrum Disorder. Journal of autism and developmental disorders. 2016;46(8):2756–63. doi:10.1007/s10803-016-2820-7 | Wrong outcomes |
| 25 | Kaltenegger HC, Philips B, Wennberg P. Autistic traits in mentalization-based treatment for concurrent borderline personality disorder and substance use disorder: Secondary analyses of a randomized controlled feasibility study. Scandinavian journal of psychology. 2019;61(3):416–22. doi:10.1111/sjop.12595 | Wrong patient population |
| 26 | Kings College London I of P. Cognitive behaviour therapy for co-morbid obsessive compulsive disorder in autism spectrum disorder [Internet]. 2007. Available from: https://www.isrctn.com/ISRCTNISRCTN87114880 | Wrong comparator |
| 27 | Klebanoff S.M., Rosenau K.A., Wood J.J. The therapeutic alliance in cognitive-behavioral therapy for school-aged children with autism and clinical anxiety. Autism. 2019;23(8):2031EP – 2042. Located at: 627144337. doi:10.1177/1362361319841197 | Wrong study design |
| 28 | Kokoski C, Lunsky Y. Brief report: exploring treatment outcomes of individuals with autism spectrum disorder in a specialized dual diagnosis inpatient program. Journal on Developmental Disabilities. 2009;15(2):110. | Wrong patient population |
| 29 | Komatsu H, Ono T, Onoguchi G, Tomita H, Kakuto Y. Mediating effects of self-stigma and depression on the association between autistic symptoms and recovery in patients with schizophrenia-spectrum disorders: a cross-sectional study. BMC psychiatry. 2021;21(1):464–464. doi:10.1186/s12888-021-03472-z | Wrong comparator |
| 30 | Komatsu H, Ono T, Onouchi Y, Onoguchi G, Maita Y, Ishida Y, Maki T, Oba A, Tomita H, Kakuto Y. Polydipsia and autistic traits in patients with schizophrenia spectrum disorders. Frontiers in Psychiatry. 2023 Jul 6;14:1205138. | Wrong outcomes |
| 31 | Lipinski S, Blanke ES, Suenkel U, Dziobek I. Outpatient Psychotherapy for Adults with High-Functioning Autism Spectrum Condition: Utilization, Treatment Satisfaction, and Preferred Modifications. J Autism Dev Disord. 2019;49(3):1154–68. doi:10.1007/s10803-018-3797-1 | Wrong outcomes |

| | | |
|---|---|---|
| 32 | Makin L, Allen K, Tchanturia K. Time to notice neurodiversity in eating disorder services: a three-year real-world analysis of autism, ADHD, and AuDHD. Front Psychiatry. 2026;17:1787957. doi:10.3389/fpsyt.2026.1787957 | Wrong study design |
| 33 | Mayes S.D., Waxmonsky J.G., Baweja R., Mattison R.E., Memon H., Klein M., et al. Symptom scores and medication treatment patterns in children with ADHD versus autism. Psychiatry Research. 2020;288((Mayes, Waxmonsky, Baweja, Mattison, Memon, Klein, Hameed, Waschbusch) Department of Psychiatry, Penn State College of Medicine, Hershey Medical Center, 500 University Dr., Hershey, PA, United States):112937. Located at: 2005596674. doi:10.1016/j.psychres.2020.112937 | Wrong patient population |
| 34 | Mohandas P., McGurgan M. Changes in Working Patterns in a Community Eating Disorder Service; the Impact of the COVID-19 Pandemic and Beyond. BJPsych Open. 2025;11(Supplement 1):S198–9. Located at: 648608064. doi:10.1192/bjo.2025.10508 | Wrong source type |
| 35 | Neuhaus E., Osuna A., Tagavi D.M., Shah-Hosseini S., Simmons S., Gerdts J., et al. Clinical Characteristics of Youth with Autism or Developmental Disability during Inpatient Psychiatric Admission. Journal of Clinical Medicine. 2022;11(21):6328. Located at: 2020032639. doi:10.3390/jcm11216328 | Wrong patient population |
| 36 | Nikolić N, Sculthorpe C, Stock J, Stevens D, Eccles J. Determining unmet need: clinical relevance of suspected neurodivergence in first-episode psychosis. BJPsych bulletin. 2024;NA(NA):1–6. doi:10.1192/bjb.2024.64 | Wrong patient population |
| 37 | Plourde E.R., Ali M.M., West K.D. Psychotropic Medication and Psychosocial Service Use Among Transition Age Youth With Autism Spectrum Disorder. American journal on intellectual and developmental disabilities. 2024;129(1):1EP – 9. Located at: 643117653. doi:10.1352/1944-7558-129.1.1 | Wrong outcomes |
| 38 | Puleo CM, Kendall PC. Anxiety Disorders in Typically Developing Youth: Autism Spectrum Symptoms as a Predictor of Cognitive-Behavioral Treatment. Journal of autism and developmental disorders. 2010;41(3):275–86. doi:10.1007/s10803-010-1047-2 | Wrong patient population |
| 39 | Roberts-Collins C, Mahoney-Davies G, Russell A, Booth A, Loades M. Emotion awareness and cognitive behavioural therapy in young people with autism spectrum disorder. Autism. 2018;22(7):837–44. doi:10.1177/1362361317710215 | Wrong outcomes |

| | | |
|---|---|---|
| 40 | Sanchez A.R., Sterris J., Shoemaker E.Z. Pediatric patients with behavioral emergencies: Who's coming in and what happens while they're here? Western Journal of Emergency Medicine. 2019;20(2):VII. Located at: 628726373 | Wrong source type |
| 41 | Scott M, Jones A, Guyatt H. Longitudinal Changes in Crisis and Broader Mental Health Service Use Following Access to Digital Support Tool Brain in Hand: An Observational Cohort Study. Research Square. 2026. Located at: PPR1144591. doi:10.21203/rs.3.rs-8613829/v1 | Wrong study design |
| 42 | Storch EA, Merlo LJ, Larson MJ, Geffken GR, Lehmkuhl HD, Jacob ML, et al. Impact of comorbidity on cognitive-behavioral therapy response in pediatric obsessive-compulsive disorder. Journal of the American Academy of Child and Adolescent Psychiatry. 2008;47(5):583–92. doi:10.1097/chi.0b013e31816774b1 | Wrong patient population |
| 43 | Tchanturia K, Larsson E, Adamson J. How anorexia nervosa patients with high and low autistic traits respond to group Cognitive Remediation Therapy. BMC psychiatry. 2016 Sep 29;16(1):334. | Wrong outcomes |
| 44 | Thorsteinsdottir S, Olafsdottir AS, Traustadottir OU, Njardvik U. Changes in Anxiety following Taste Education Intervention: Fussy Eating Children with and without Neurodevelopmental Disorders. Nutrients. 2023;15(22). doi:10.3390/nu15224783 | Wrong patient population |
| 45 | Tint A, Chung H, Lai MC, Balogh R, Lin E, Durbin A, et al. Health conditions and service use of autistic women and men: A retrospective population-based case-control study. Autism : the international journal of research and practice. 2023;27(6):1641–57. doi:10.1177/13623613221144353 | Wrong intervention |
| 46 | Tsuchiyagaito A., Hirano Y., Tazaki M., Nakagawa A. Neuro structural predictors of cognitive behavioral therapy (CBT) for obsessive-compulsive disorder: Implications for the integration of neuro feedback training and CBT. NeuroRegulation. 2017;4(3–4):144EP – 145. Located at: 626603456. doi:10.15540/nr.4.3-4.100 | Wrong comparator |
| 47 | Vergara GA, Batejan KL, Worden P, Antkiewicz N, Brown KJ, Shogren N, et al. Adolescent Autism Traits at a DBT Partial Hospital: Treatment Outcomes, Suicidality, and Therapeutic Alliance. Clin Child Psychol Psychiatry. 2026 Apr;31(2):785–96. doi:10.1177/13591045251408156 | Wrong comparator |
| 48 | Vohra R, Madhavan S, Sambamoorthi U. Emergency Department Use Among Adults with Autism Spectrum Disorders (ASD). Journal of autism and developmental disorders. 2016;46(4):1441–54. doi:10.1007/s10803-015-2692-2 | Wrong outcomes |

| | | |
|---|---|---|
| 49 | Wielemaker J., Verheij F., Verhulst F. Follow-up of long-term child psychiatric residential care. European Child and Adolescent Psychiatry. 2011;20(SUPPL. 1):S76EP – S77. Located at: 70489392. doi:10.1007/s00787-011-0181-5 | Wrong source type |
| 50 | Wikramanayake WNM, Mandy W, Shahper S, Kaur S, Kolli S, Osman S, et al. Autism spectrum disorders in adult outpatients with obsessive compulsive disorder in the UK. International journal of psychiatry in clinical practice. 2017;22(1):54–62. doi:10.1080/13651501.2017.1354029 | Wrong intervention |
| 51 | Wolters LH, de Haan E, Hogendoorn SM, Boer F, Prins PJ. Severe pediatric obsessive compulsive disorder and co-morbid autistic symptoms: Effectiveness of cognitive behavioral therapy. Journal of Obsessive-Compulsive and Related Disorders. 2016 Jul 1;10:69-77. | Relevant outcome data could not be obtained |
| 52 | Wyman J, Claro A. The UCLA PEERS school-based program: Treatment outcomes for improving social functioning in adolescents and young adults with autism spectrum disorder and those with cognitive deficits. Journal of Autism and Developmental Disorders. 2020;50(6):1907–20. | Wrong intervention |
| 53 | Zheng S, Chua YC, Tang C, Tan GMY, Abdin E, Lim VWQ, et al. Autistic traits in first-episode psychosis: Rates and association with 1-year recovery outcomes. Early intervention in psychiatry. 2020;15(4):849–55. doi:10.1111/eip.13021 | Wrong comparator |
| 54 | Zijlmans J, van Rijn L, Alrouh H, Broek E, Luijten M, Tieskens J, van der Mheen M, van Oers H, Cahn W, Schellekens A. Mental health changes during and after the COVID-19 pandemic in children and adolescents with mental disorders. European Child & Adolescent Psychiatry. 2025 Sep;34(9):2833-42. | Wrong comparator |
| 55 | Zlomke KR, Jeter K. Comparative effectiveness of Parent-Child Interaction Therapy for children with and without autism spectrum disorder. Journal of Autism and Developmental Disorders. 2020;50(6):2041–52. doi:10.1007/s10803-019-03960-y | Wrong intervention |
| 56 | Zukerman G, Ben-Itzchak E. Compulsion Profile Differences Indicate Distinct Functional Mechanisms in Autistic and Non-Autistic University Students. Autism Res. 2026;e70215. doi:10.1002/aur.70215 | Wrong study design |

### **Appendix E.** List of measures included in the review with references.

| **Measure** | **Reference** |
|---|---|
| Anorexia Nervosa Total Severity Index (ANTSI) of the Short Evaluation of Eating Disorders (SEED) | (Bauer et al., 2005) |
| Anxiety Disorders Interview Schedule for DSM-IV: Child and Parent versions (ADIS-C/P) | (Silverman & Albano, 1996) |
| Autism Diagnostic Observation Schedule (ADOS) | (Lord et al., 1989) |
| Autism Diagnostic Observation Schedule, Second Edition (ADOS-2) | (Lord et al., 2012) |
| Autism Diagnostic Interview-Revised (ADI-R) | (Lord et al., 1994) |
| Autism-Inclusive Research Assessment (AIRA) | (Loizou et al., 2023; Pemovska et al., 2024) |
| Autism Quotient-10 (AQ-10) | (Allison et al., 2012) |
| Autism Quotient-50 (AQ-50) | (Baron-Cohen et al., 2001) |
| Autism Quotient-Japanese version (AQ-J) | (Wakabayashi et al., 2006) |
| Autism Spectrum Disorder in Adults Screening Questionnaire (ASDASQ) | (Nylander & Gillberg, 2001) |
| Autism Spectrum Screening Questionnaire (ASSQ) | (Ehlers et al., 1999) |
| Beck Depression Inventory-II (BDI-II) | (Beck et al., 2011) |
| Brief Psychiatric Rating Scale (BPRS) | (Overall & Gorham, 1962) |
| Bulimia Nervosa Total Severity Index (BNTSI) of the Short Evaluation of Eating Disorders (SEED) | (Bauer et al., 2005) |
| Calgary Depression Scale for Schizophrenia (CDSS) | (Addington et al., 1990) |
| Child Behavior Checklist (CBCL) | (Achenbach, 1991) |
| Child OCD Impact Scale-Child/Adolescent Report and Parent Report (COIS-C/P) | (Piacentini et al., 2001) |
| Children’s Global Assessment Scale (CGAS) | (Shaffer, 1983) |
| Children's Yale-Brown Obsessive-Compulsive Scale (CY-BOCS) | (Scahill et al., 1997) |
| Clinical Global Impression (CGI) Scale | (Guy, 1976) |
| Clinical Global Improvement (CGI) scale of the National Institute of Mental Health Global Obsessive-Compulsive Scale (NIMH-GOCS) | (Insel et al., 1983) |
| Clinical Impairment Assessment (CIA) | (Bohn et al., 2008) |
| Clinical Outcomes in Routine Evaluation – Outcome Measure (CORE-OM) | (Evans et al., 2002) |
| Critical Appraisal Skills Programme (CASP) Cohort Study Checklist | (Critical Appraisal Skills Programme, 2024) |

| | |
|---|---|
| Depression, Anxiety and Stress Scale (short-form version) (DASS-21) | (Henry & Crawford, 2005) |
| Developmental and Well-Being Assessment (DAWBA) | (R. Goodman et al., 2000) |
| Diagnostic and statistical manual of mental disorders (4th edition) (DSM-IV) | (American Psychiatric Association, 1994) |
| Diagnostic and statistical manual of mental disorders (4th edition, text rev.) (DSM-IV-TR) | (American Psychiatric Association, 2000) |
| Diagnostic and statistical manual of mental disorders (5th edition) (DSM-5) | (American Psychiatric Association, 2013) |
| Difficulties in Emotion Regulation Scale (DERS) | (Gratz & Roemer, 2004) |
| Eating Disorder Examination Questionnaire (EDE-Q) | (Fairburn & Beglin, 1994) |
| Eating Disorder Inventory, Third Edition (EDI-3) | (Garner, 2004) |
| Eating Disorder Quality of Life Scale (EDQLS) | (Adair et al., 2007) |
| EuroQol 5-D (EQ-5D) | ('EuroQol - a New Facility for the Measurement of Health-Related Quality of Life', 1990) |
| Family Functioning Scale (FFS) | (Bloom, 1985) |
| Functioning Assessment Short Test | (Rosa et al., 2007) |
| Generalized Anxiety Disorder-7 | (Spitzer et al., 2006) |
| Global Assessment of Functioning (GAF) Scale | (Endicott, 1976) |
| Health of the Nation Outcome Scales (HoNOS) | (Wing et al., 1998) |
| Hospital Anxiety and Depression Scale (HADS) | (Zigmond & Snaith, 1983) |
| International Classification of Diseases, 9th Revision (ICD-9) | (World Health Organization, n.d.) |
| International Statistical Classification of Diseases and Related Health Problems, 10th Revision (ICD-10) | (World Health Organization, 1992) |
| Kiddie Schedule for Affective Disorders and Schizophrenia – Present and Lifetime Version (K-SADS-PL) | (Kaufman et al., 1997) |
| McLean Screening Instrument for Borderline Personality Disorder (MSI-BPD) | (Zanarini et al., 2003) |
| National Institute of Mental Health Global Obsessive-Compulsive Scale (NIMH-GOCS) | (Insel et al., 1983) |
| Moods and Feelings Questionnaire for Young People (MFQ-YP) | (Angold & Costello, 1987) |
| Morgan-Russell criteria | (Morgan & Russell, 1975) |

| | |
|---|---|
| Obsessive Compulsive Disorder (OCD) – Child Obsessional Compulsive Inventory, self and parent report (CHOCI) | (Shafran et al., 2003) |
| Outcome Questionnaire-45 (OQ-45) | (Lambert et al., 2004) |
| PANSS Autism Severity Score (PAUSS) | (Kästner et al., 2015) |
| Patient Health Questionnaire-9 (PHQ-9) | (Kroenke et al., 2001) |
| Pediatric Anxiety Rating Scale (PARS) | (The Research Units On Pediatric P, 2002) |
| Positive and Negative Syndrome Scale (PANSS) | (Kay et al., 1987) |
| Questionnaire about the Process of Recovery (QPR) | (Neil et al., 2009) |
| Screen for Child Anxiety Related Emotional Disorders-71 (SCARED-71) | (Bodden et al., 2009) |
| Screen for Child Anxiety Related Emotional Disorders – Young Person version (SCARED-YP) | (Birmaher et al., 1997) |
| Self-Administered Psychiatric Scales for Children and Adolescents | (Cianchetti & Fancello, 2001) |
| Sheehan Disability Scale (SDS) | (Sheehan et al., 1996) |
| Short Evaluation of Eating Disorders (SEED) | (Bauer et al., 2005) |
| Short-form Scale for the Assessment of Positive Symptoms (SAPS) | (Andreasen, 1984) |
| Social and Occupational Functioning Assessment Scale (SOFAS) | ('Revising Axis V for DSM-IV', 1992) |
| Social Anhedonia Scale (SAS) | (Chapman et al., 1976) |
| Social Aptitude Scale (SAS) | (Liddle et al., 2009) |
| Social Responsiveness Scale-2 (SRS-2) | (Volkmar, 2021) |
| Strengths and Difficulties Questionnaire (SDQ) | (R. Goodman, 1997) |
| Structured Clinical Interview for DSM-IV Axis I Disorders (SCID-IV) | (First & Gibbon, 2004) |
| Structured Clinical Interview for DSM-IV Axis II Personality Disorders (SCID-II) Borderline Personality Disorder (BPD) module | (First et al., 1997) |
| Toronto Alexithymia Scale (TRS) | (Bagby et al., 1994) |
| Tower of London test, Japanese version of the Brief Assessment of Cognition in Schizophrenia (BACS) | (Keefe, 2004) |
| Wechsler Adult Intelligence Scale-Third Edition (WAIS-III) | (Wechsler, 1997) |
| Work and Social Adjustment Scale (WSAS) | (Mundt et al., 2002) |
| Yale-Brown Obsessive-Compulsive Scale (Y-BOCS) | (W. K. Goodman, 1989) |
